# Protocol for Bayesian combined multi-genotype and concentration informed tacrolimus dosing in paediatric solid organ transplantation (BRUNO-PIC)

**DOI:** 10.1101/2025.05.06.25327120

**Authors:** Dhrita Khatri, Andreas Halman, Joshua Kausman, Jacob Mathew, Elizabeth Bannister, Tayla Stenta, Claire Moore, Elizabeth Williams, Roxanne Dyas, Julian Stolper, Cecilia Moore, Mark Pinese, Rishi S. Kotecha, Christopher Gyngell, Sebastian Lunke, John Christodoulou, Amanda Gwee, Rachel Conyers, David Metz

## Abstract

**Introduction:** Tacrolimus is an immunosuppressant used extensively in solid organ transplantation. Whilst highly effective in preventing organ rejection, it has a narrow range of safe and effective concentrations and wide pharmacokinetic variability, which can lead to and suboptimal outcomes or unacceptable toxicities. Importantly, much is known about the sources of pharmacokinetic variability. There is a clear link between body size, pharmacogenetic variants *of cytochrome P450 (CYP) 3A4* and *CYP3A5* and typical dose requirement. Increasing individualisation of initial dose is expected to increase the proportion of transplant recipients with tacrolimus concentrations within the acceptable range in the initial post-transplant week. Subsequently, maximum *a posteriori* Bayesian dosing can better maintain concentrations within the acceptable range over time. Together, this should lead to improved patient outcomes.

**Method and analysis:** BRUNO-PIC is an open-label trial with a prospective intervention arm and a retrospective standard of care comparator arm. The prospective arm evaluates covariate informed initial dosing combined with Bayesian dose adjustment in children undergoing kidney, liver or heart transplant. *CYP3A5* and *CYP3A4* genotyping will be combined with allometric size scaling to predict initial tacrolimus dose. Post-transplant dose adjustment will be guided by NextDose, a Bayesian dosing platform that incorporates genotype, clinical characteristics, measured tacrolimus concentrations and a population pharmacokinetic (popPK) model to inform initial dosing and guide dose adjustments following transplant. The primary objective of this study is to determine whether genotype-informed Bayesian dosing of tacrolimus leads to better achievement of tacrolimus concentrations within the acceptable range over the first 8 weeks post-transplant in paediatric solid organ transplant (SOT) recipients (kidney, heart and liver), when compared to a retrospective historical control group using standard of care dosing. The primary outcome is the proportion of each cohort with tacrolimus concentration (steady-state average concentration) within the acceptable range of 80-125% of target at post-transplant dosing day 4 (DD4), at week 3 and at week 8. Secondary outcomes include the proportion with trough concentration within the acceptable range on DD4, the median time to acceptable range, and the time within acceptable range over the first 8-weeks post-transplant.

**Ethics and dissemination:** The ethics approval of the trial has been obtained from the Sydney Children’s Ethics Committee (2023/ETH02699). Findings will be disseminated through peer-reviewed publications and professional conference presentations.

**Trial registration:** ClinicalTrials.gov NCT 06529536

**STRENGTHS AND LIMITATIONS OF THIS STUDY:** *Strengths:* This is a prospective study in paediatric solid organ transplant recipients combining (a) pre-transplant *CYP3A5*/*3A4* genotyping to improve initial tacrolimus dose, and (b) post-transplant Bayesian individualised tacrolimus dosing to increase time within the safe and effective range. Use of a target concentration approach based on average steady state concentrations is expected to increase exposure in individuals with low tacrolimus clearance and reduce overexposure in individuals with high tacrolimus clearance (e.g. CYP3A5 expressors).

*Limitations:* The lack of a randomised comparator arm in this prospective interventional trial precludes unconfounded determination of superiority over standard care. The intervention may not be generalisable to other transplant recipients (e.g. lung, intestinal, hematopoietic stem cell transplant).

**ADMINISTRATIVE INFORMATION:** *Protocol Version:* This trial is approved Sydney Children’s Ethics Committee (2023/ETH02699). The study is currently version 4, 26.05.2025 (Supplementary File 2)

*Funding:* BRUNO-PIC is funded by the 2023 Medical Research Future Fund – Genomics Health Future Mission (MARVEL-PIC (MRF/2024900). This funding source had no role in the design of this study and will not have any role during its execution, analyses, interpretation of the data, or decision to submit results.

*Sponsorship:* Murdoch Children’s Research Institute

## 1. INTRODUCTION

### 1.1 Background and rationale

Patient outcomes from solid organ transplantation (SOT) saw major advances in the last two decades of the 20th century, with introduction of highly effective immunosuppressant drugs including tacrolimus, mycophenolate and anti-lymphocyte antibody induction ^1^. However, despite appreciable short-term gains, long-term graft and patient outcomes remain suboptimal ^2^ ^3^. These suboptimal long-term outcomes are influenced both by immunosuppressant exposure – current and cumulative – in the maintenance phase ^2^, but also to alloimmune events and subclinical rejection in the early months post-transplant ^4^.

Tacrolimus, a calcineurin inhibitor (CNI) immunosuppressant produced by the bacterium *Streptomyces tsukubaensis* ^5^, is a core component of modern SOT immunosuppression. It is used in the majority of recipients, surpassing the CNI cyclosporine after superiority demonstrated in the Elite-Symphony trial in 2007 ^6^.

As a “narrow therapeutic index” drug, tacrolimus requires concentration-controlled dosing ^7^ for safety and effective use ^8^ ^9^. Yet current practice - empiric dose adjustment to attain trough (pre-dose) tacrolimus concentrations within a “therapeutic window” ^10^ - only partially compensates for tacrolimus dose-response variability. The consequences of suboptimal dosing remain, with treatment failure in some ^11–16^, unacceptable toxicities in others ^2^ ^3^ ^17^.

#### Initial tacrolimus dose

Despite low incidence of early acute rejection in contemporary cohorts, acute rejection associated with low tacrolimus concentrations is still seen ^18–22^. One predictable contributor to initial underexposure – if not accounted for – is the group of individuals with *CYP3A5* expression, typically requiring 1.5 to 2-fold higher dose than those with non-expressor genotype ^18–22^. This is compounded by slow empiric dose titration, with some recipients not reaching target for up to 3 weeks ^23^.

Greater “individualisation” of initial tacrolimus dosing increases the proportion of recipients within the acceptable concentration range in the first post-transplant week. Meta-analysis of randomised controlled trials (RCTs) demonstrates that *CYP3A5* genotype-guided initial dosing increases the proportion of recipients within the acceptable range in the first week post-transplant from 31.5% to 44.1%, along with a reduction in median time to target ^24^. Subsequent trials report an even higher proportion of recipients within the acceptable range, 54.8% and 58% respectively, using combined *CYP3A4* and *CYP3A5* genotypes and superior size scaling in adult kidney transplant recipients ^23^ ^25^. This has not been tested in children to date.

Finally, whilst tacrolimus dose is typically scaled to size by simple linear function (dose/kg × body weight), linear scaling is inconsistent with the change in drug clearance with body size, leading to underexposure in young children ^26^. This has been empirically shown for tacrolimus in paediatric transplantation, with recommendation for higher per-kilogram dose at younger age or lower body weight ^27^ ^28^. Alternatively, allometric scaling describes the relationship between size and drug clearance across all body sizes, as well as having robust theoretic underpinnings ^26^ ^29^. This allows accurate size scaling of dose from adults through to infancy or early childhood, down to the age at which the drug’s clearance mechanisms have matured (12 months of age for tacrolimus).

#### Tacrolimus pharmacokinetics and initial dose

Tacrolimus undergoes hepatic clearance, with metabolism by *CYP3A4*, and by *CYP3A5* in individuals who express this enzyme ^30^ ^31^. These drug-metabolising enzymes reside predominantly in the liver, though are also present in enterocytes, where metabolism and P-glycoprotein-mediated efflux impact bioavailability.

All individuals express *CYP3A4*, with minimal levels at birth, increasing throughout infancy and reaching full adult values by one year of age ^32^. On the other hand, *CYP3A5* expression is present in 10-15% of Caucasian populations, 40%-50% of African and 50-70% of Asian populations ^33^. Individuals expressing *CYP3A5* require higher tacrolimus dosing to achieve equivalent exposure early post-transplant ^31^. The Clinical Pharmacogenetics Implementation Consortium (CPIC) and the French National Network of Pharmacogenetics (RNPGx) recommending a 1.5 to 2-fold increase in dose for both heterozygous and homozygous expressors, whilst the Dutch Pharmacogenetics Working Group (DPWG) recommending 1.5-fold increase for heterozygous expressors and 2.5-fold increase for homozygous *CYP3A5* expressors ^34^.

Pharmacogenetic variations in *CYP3A4* are quantitatively less pronounced than in *CYP3A5*, but a clinically relevant association with the *CYP3A4*\*22 polymorphism and lower dose requirements is increasingly recognised ^35^. The *CYP3A4*\*22 polymorphism is more common in European populations, where the association has been more clearly demonstrated ^36^.

In liver transplantation, the picture is more complex. Following transplantation, the recipient’s own *CYP3A* genotype contributes to CYP expression in enterocytes (influencing tacrolimus bioavailability), whilst the new liver contains the donor’s *CYP3A* genotype (contributing to both first-pass metabolism and drug clearance) ^37^ ^38^. Thus, the influence of recipient *CYP3A* genotype on tacrolimus disposition is less given impact on enterocyte *CYP3A* expression only. In popPK analyses in liver transplantation where both donor and recipient genotype were available, with the proportional increase in tacrolimus apparent clearance (CL/F) in recipient *CYP3A5* expressers is around half of those with both donor and recipient CPY3A5 expresser status (average across studies, a 38% increase compared to a 78% increase, respectively) ^37–41^. Thus, whilst there is precedence for applying the same proportion increase in tacrolimus dose for CYP3A5 expressers across all SOT recipients (including liver transplantation) ^27^, we favour an empiric reduction (halving) in the influence of recipient genotype in liver transplant recipients, given available data and mechanistic basis.

In addition, CYP isoenzyme expression is initially poor due to allograft ischaemia-reperfusion injury ^42^, a pro-inflammatory state associated with cellular dysfunction, including downregulation of CYP isoenzyme expression ^43–45^. Whilst liver transaminases typically start to decline within days post-transplant ^46^ ^47^, this does not directly correlate with CYP isoenzyme expression, with tacrolimus CL/F taking up to 2-3 weeks to reach full maturity ^39^ ^41^ ^48^. Thus, with tacrolimus CL/F increasing over the initial weeks post liver transplantation (due to an increase in intrinsic hepatic clearance), dose requirement increases too, a dynamic ideally accounted for in dose adjustment predictions.

#### Bayesian dosing to increase time within acceptable range of tacrolimus concentrations

In addition to the consequences of low tacrolimus concentrations in the initial post-transplant week in SOT recipients, there is a robust association between the proportion of time with tacrolimus concentrations outside the acceptable range over the first 6-months, and negative clinical outcomes. This includes association with acute rejection ^11^ ^49–51^, donor-specific antibody formation ^12^ ^14–16^ ^52^ and post-transplant malignancy ^53^, as well as overall poorer long-term patient and graft outcomes ^50^ ^52^ ^54–58^.

Yet maintaining tacrolimus concentrations within the acceptable range can be challenging, particularly in the initial post-transplant months. Published experience reports less than 60% of measurements within a trough concentration based therapeutic window ^59^ using a therapeutic window approach for dose adjustment (TWA)^7^. This is in part due to substantial between-occasion variability in tacrolimus PK early post-transplant, with contribution from variability in body size, haematocrit (HCT), prednisolone dose, as well as time-post transplant, an empirical predictor of early increased oral bioavailability ^49^ ^59–61^. In addition, empiric titration of dose based on TCA is both highly user-dependant and less accurate than pharmacokinetic-driven techniques ^10^.

Increasing time in range has been achieved using maximum *a posteriori* Bayesian estimation (Bayesian dosing), with superiority to TWA reported in two RCTs in adult kidney transplant recipients ^62^ ^63^. Bayesian dosing is a pharmaco-statistical technique that predicts a drug’s pharmacokinetic parameters in the individual - hence dose requirement - by leveraging prior population information about a drugs pharmacokinetic properties (a popPK model) with their measured drug concentrations and clinical characteristics. Maximum likelihood estimation is used to determine the “best fit” concentration time course to observed concentrations, from which is derived individual estimates of a drug’s PK parameters. There are then used to calculate dose required to achieve desired target concentration ^10^.

Although Bayesian dosing is routine in some transplanting centres ^64^ ^65^, these remain in the minority ^66^ ^67^. Barriers to implementation include (a) an evolving regulatory framework ^68^; (b) clinician acceptance, including choice of Bayesian platform and (c) lack of clinical trial evidence showing improvement in long-term outcomes ^67^. On the final point, however, it is crucial to note the substantial challenge in proving clinical superiority by RCT in modern transplant cohorts, given low rates of events in the first 3-6 months post-transplant (rejection, graft failure) ^64^ ^65^ ^69^. Whilst clearly advantageous, the low prevalence of hard endpoints in the first post-transplant year has made it harder to prove value of new interventions difficult, with very large numbers required for power to detect superiority ^2^ ^3^. This has led to academic ^3^ and regulatory calls ^70^ for innovative approaches, including surrogate biomarkers of long-term outcome.

In this context, tacrolimus Bayesian dose adjustment has longstanding precedence for use in some parts of the world such as Europe and Asia ^62^ ^63^, along with RCT evidence in adult kidney transplant recipients showing increased time within the acceptable range in the immediate post-transplant week and over the subsequent 3 months ^62^ ^63^. This has not been evaluated in children to date.

#### Bayesian dosing and NextDose

There are a range of platforms for Bayesian dosing ^59^ ^67^. The use of NextDose, noting the developer (Prof Nick Holford) is a co-investigator on BRUNO-PIC ^71^, allows for iterative refinement of the underlying model as required (e.g. for non-renal SOT recipient). Furthermore, there is published evidence supporting superiority of the tacrolimus model within NextDose.

The Størset tacrolimus model used by NextDose was developed from a large population dataset, with 242 kidney transplant recipients and 3100 tacrolimus whole blood concentrations. This included 64 full PK profiles (> 8 concentrations per occasion), over 153 limited sampling profiles and 1546 trough concentrations ^61^. It was developed by combining two separate populations, the pooled model outperforming those from which it was developed in external validation ^61^. The model was developed using principles of mechanistic modelling for both structure and covariate introduction ^61^ ^72^, a principle with clear basis in pharmacometrics (and statistical modelling more generally) to reduce bias and enhance external validity ^73^. Finally, its superiority has been reported in a published systematic review and external validation, e.g. *“the model by Størset et al, which was comparatively the best of all 16 published models…*” ^72^.

Whilst the NextDose tacrolimus model was developed in an adult kidney transplant population, this is not a barrier to use in children who have reached an age when the clearance mechanisms are mature after which, from a PK perspective, children *are* “small adults” ^26^. Full maturation of clearance mechanisms occurs before 1-2 years of age for tacrolimus. Therefore, application of allometric size scaling can be used to predict dose requirement in children using models developed with adult popPK (popPK) data ^26^ ^74^. Further, research groups have developed separate popPK models in children and in adults (excluding infancy) demonstrating similar PK parameters and concentration-time course (scaled for size) ^64^.

For use of NextDose in non-kidney transplantation, a further validation step is underway. Importantly, whilst the tacrolimus popPK model for NextDose was developed in a kidney transplant population, there is published precedence for pooling of popPK models between kidney and non-kidney transplant recipients ^65^ ^75^ ^76^. These population “meta-models” show *liver transplant recipients* initially have 50% lower tacrolimus CL/F recovering to stable values after 2 weeks (mechanisms discussed above). Importantly, by adding a parameter to account for reduced clearance early post liver transplantation, models initially developed in kidney transplant recipients have been shown to be able to predict tacrolimus concentrations in liver transplant recipients ^75^ ^76^. Finally, both liver and heart transplant recipients have lower tacrolimus CL/F beyond the initial weeks ^75^ ^77^ when compared with kidney transplant recipients ^78^. This published data has been used to add additional parameters for reduced clearance in NextDose, to allow for first dose prediction in liver and heart transplant recipients following internal validation.

#### Concentration target for dose titration: trough or average concentration

The long-accepted paradigm for tacrolimus dose titration is to use a concentration measured just before a dose (Ctrough). However, whether this is sufficient to optimise safety and effectiveness of tacrolimus is being increasingly challenged ^79^.

Whilst Ctrough is a pragmatic target in clinical care ^80^, it is used as a surrogate for overall drug exposure (the concentration area under the curve, AUC), the latter more directly linked with clinical effect. This is well established for cyclosporine, the predominant CNI before prior to tacrolimus ^81^. For tacrolimus, whilst most of the exposure-response data is with Ctrough, the same principle is assumed to apply given same downstream pharmacodynamic mechanism ^81^. This is supported by early exposure-response data linking AUC with drug effect ^82^. Subsequently, real-world data has shown substantial between-subject variability in AUC at a given trough concentration ^83^, with more recent data showing AUC as more predictive of late rejection than Ctrough in kidney transplantation ^84^.

There is substantial precedence for AUC-guided dosing as standard of care, from over two decades of use in certain regions with local expertise (e.g. Europe) ^17^ ^83–88^, though without broader global uptake.

Those most likely to benefit from the more precise exposure-guided dosing are pharmacokinetic ‘outliers’, i.e. *CYP3A5* expressers or those with other cause for high tacrolimus clearance ^33^ ^79^ ^87^. These individuals typically require substantially higher dose to achieve the same tacrolimus Csstrough ^89^ ^90^, often translating to higher peak concentrations and higher overall tacrolimus systemic exposure ^33^ ^87^ ^91^. Higher tacrolimus exposure despite Csstrough within acceptable range explain why these individuals see an increase toxicity including CNI nephrotoxicity ^89^ ^92–96^, BK virus nephropathy ^97–99^ and other toxicities ^33^ ^90^ ^100^.

Thus, with current tacrolimus dosing practice, *CYP3A5* expressors are at risk of rejection if they fail to achieve acceptable exposure in the early post-transplant period, and toxicities over time due to chronic over-exposure, with overall inferior graft and patient survival ^33^ ^79^ ^90^ ^100^. Greater individualisation of initial dose, followed by Bayesian dosing to exposure (AUC), has potential to ameliorate both of these issues, in CYP3A5 expressors, and in PK outliers of other aetiology.

### 1.2 OBJECTIVES

#### Primary Objective

- To determine whether genotype-informed Bayesian dosing of tacrolimus in paediatric SOT recipients (kidney, heart and liver) increases time with tacrolimus concentrations within the acceptable range over the first 8 weeks post-transplant, compared with a retrospective historical control group using standard of care dosing.

#### Secondary objectives

- To determine the safety and effectiveness of using a genotype-informed Bayesian dosing of tacrolimus in SOT within the initial 8 weeks post-transplant (prospective arm only, descriptive)
- To compare achievement and maintenance of acceptable concentrations achieved with genotype-informed dosing of tacrolimus with a retrospective group using standard of care dosing.
- To evaluate the feasibility of using NextDose for concentration guided dosing in the context of paediatric SOT.

### 1.3 METHODS: TRIAL DESIGN

BRUNO-PIC is a prospective, open-label interventional trial, with retrospective cohort as standard of care comparator. The prospective arm will recruit 45 children undergoing kidney, liver or heart transplantation, who will have pre-transplant genotyping of *CYP3A4* and *CYP3A5*, and initial tacrolimus dose using genotype and allometric size scaling. Subsequent dosing over the first 8 weeks post-transplant will be by Bayesian estimation using NextDose.

The retrospective cohort will consist of approximately 120 eligible paediatric recipients of kidney, liver, or heart transplants at the RCH over a 5-year period (January 2019 to April 2024). This cohort represents current tacrolimus dosing practice at the Royal Children’s Hospital (RCH), which involves a standard mg/kg protocol followed by trough concentration measurement and empiric dose adjustment using the TWA.

The ability of genotype-informed Bayesian-dosing to optimise tacrolimus exposure (concentrations within the acceptable range initially and over time) will be compared to the retrospective cohort.

## 2. METHODS: PARTICIPANTS, INTERVENTIONS, AND OUTCOMES

### 2.1. Trial setting

BRUNO-PIC will be conducted at a single tertiary institute in Victoria, Australia, enrolling children undergoing SOT (either kidney, liver or heart) at the RCH.

### 2.2. Eligibility criteria

#### Inclusion criteria

▪ Age 1-18 years of age
▪ Kidney, liver or heart transplant recipients
▪ Participant and/or parent consent to the study (prospective arm only)

#### Exclusion criteria

▪ Previous liver transplant.
▪ Lung OR Intestinal transplant.
▪ Insufficient time before transplant for pharmacogenomic analysis (prospective arm only)
▪ Immunosuppressant regimen not containing tacrolimus immediate release product
▪ Known hypersensitivity to tacrolimus and/or its formulation.

#### Participant enrolment: phase 1 and 2

A total of 45 individuals will be recruited prospectively to the trial, with phased opening of cohorts based on SOT type:

**Phase 1**: Enrolling kidney transplant recipients, the population in whom the popPK model used by NextDose was developed.

**Phase 2:** Enrolling other SOT recipients (liver and heart). This second phase will commence following evaluation of the NextDose model with adjustment for organ type (liver or heart), with acceptability of predicted doses for non-kidney SOT recipients based on virtual performance using retrospective data.

#### Evaluation of NextDose for non-renal organ transplant recipients

Additional parameters have been added to the NextDose tacrolimus model to account for reduced tacrolimus CL/F in heart and liver recipients in the first post-transplant week, based on published population PK models (including models pooling and comparing organ types) ^65^ ^75^ ^76^. Prior to use of the adjusted model in prospective heart and liver transplant recipients, the model performance will be evaluated using retrospective data, by assessing iterative dose recommendations over the first post-transplant week. Performance will be assessed against achieved tacrolimus concentrations, and with reference to accepted standards for bias and precision: bias by mean prediction error (percentage) ±15-20% and precision by mean absolute percentage error ≤20%^101^.

### 2.3. Intervention

#### Initial dose

Initial dose in BRUNO-PIC will use allometric size scaling from adult dose, with adjustment based on genotype (*CYP3A4* & *CYP3A5*).

Dose is scaled for size from a standard adult dose administered every 12 h. Taking the example of kidney transplantation, this is 5 mg in a 70 kg adult. Size scaling is by allometry, with fat free mass (FFM) as size descriptor, using the equation:

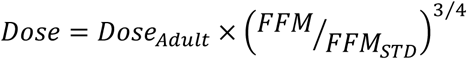

*where FFM_STD_ is 56.1 kg, based on a 70 kg 176 cm male adult*.

Genotyping of *CYP3A4* and *CYP3A5* will occur prior to transplantation in the prospective intervention arm. Genotypes of relevance to tacrolimus are:

- CYP3A4 Homozygous or heterozygous for *22 (e.g. *1/*22, *22/*22)
- CYP3A4 No *22 allele (e.g. *1/*1)
- CYP3A5 Normal or intermediate metaboliser (e.g. *1/*1, *1/*3)
- CYP3A5 Poor metaboliser (e.g. *3/*3, *6/*7)

Genotyping will be performed using Illumina’s genome-wide genotyping array (Infinium Global Screening Array). Pre-transplant genotyping will test for *CYP3A5* *3, *6, *7, *8 and *9 alleles, and will test for *CYP3A4*\*22 only (with *CYP3A4*\**1* reported if no variant corresponding to *22 was present). The results for *CYP3A4* and *CYP3A5* will be automatically extracted from the array data using an in-house program. The determined diplotype for *CYP3A5* will be matched with the predicted phenotype using the CPIC proposed genotype-to-phenotype translation table. The assignment of the phenotype is outlined in the CPIC guidelines^31^. In addition, influence of *CYP3A4* will be incorporated based on recent literature and interventional trials, see section 2.3 and Supplementary File 2 for detail.

Though both *CYP3A5*\*6 and *7 are rare in the population, they result in nonfunctional protein, and their impact on tacrolimus clearance and dose-adjusted trough concentrations is therefore presumed to be equivalent to *3 ^31^, thus treated here as functionally equivalent to *3 for dose adjustments ^61^.

Based on the literature for CYP3A5 ^61^ and CYP3A4 ^23^ ^102^ in ***kidney transplant recipients***, the group differences in tacrolimus CL/F and drug dose are as follows:

- CYP3A4 no *22 allele present & CYP3A5 non-expressor: Dose x 1 (No change)
- CYP3A4 homozygous or heterozygous for *22: Dose x 0.74
- CYP3A5 expressor (normal or intermediate metaboliser): Dose x 1.59 (fCL/F=1.3/0.82)
- CYP3A4 homozygous or heterozygous for *22 allele and CYP3A5 expressor: Dose x 1.18 (0.74 x 1.59)

It is assumed that the genotype effects on tacrolimus CL/F are the same for ***heart transplant recipients***, consistent with published data ^103^.

For ***liver transplant recipients***, the impact of recipient genotype will be reduced by 50%, as follows:

- CYP3A4 no *22 allele present & CYP3A5 non-expressor: Dose x 1 (No change)
- CYP3A4 homozygous or heterozygous for *22: Dose x 0.87
- CYP3A5 expressor (normal or intermediate metaboliser): Dose x 1.29
- CYP3A4 homozygous or heterozygous for *22 allele and CYP3A5 expressor: Dose x 1.09

The BRUNO-PIC first dose should commence following transplantation as soon as able and/or consistent with clinical unit guidelines (delayed until day 4 post-transplant in for liver transplant recipients on “renal-sparing” protocol). The dose immediately prior to transplant is 50% of maintenance dose, given the known concentration-dependent vasoconstrictive effect of tacrolimus on renal microvasculature ^104^.

#### Subsequent dosing

***Subsequent dosing*** will be guided by Bayesian estimation using NextDose. Estimation will occur with every dosing occasion over the first 8 weeks that contains a tacrolimus concentration, commencing on post-transplant dosing day 4 (DD4). In addition, on DD4, additional samples will be taken for tacrolimus concentration measurement: immediately pre-dose, then, 1 h, 2 h, and 3 h after the morning dose.

Tacrolimus concentrations are measured daily for the first 2-4 weeks post-transplant in all SOT types, then with reducing frequency (eventually twice or thrice weekly). Samples are taken at between 7-9 am each morning, prior to the morning tacrolimus dose, with exact time taken documented. For each dose estimation, tacrolimus concentrations from the day of dosing, and the prior 7 days, will be used within NextDose (excluding concentrations prior to DD4).

A unique patient identifier, date of birth (or current age), sex and genotype along with time associated dose and observations (such as weight, concomitant steroid use, transplant date, transplant organ type) are recorded in NextDose, along with tacrolimus concentrations as above. Following dose prediction on each dosing occasion with measured tacrolimus concentration, predicted dose, along with graphs of predicted and observed concentrations, will be communicated on the same day to the transplant treatment team via phone, secure chat with confirmation of receipt, or in-person. The predicted dose and the administered dose will be documented in the electronic medical record (EMR).

The 4-point profile performed on DD4 will also occur at week 3 and week 8, to provide additional information for dose estimation and as outcome measures. These additional samples on DD4, week 3 and week 8, are the only blood samples taken outside of standard of care, falling within bounds of acceptable sampling volume for clinical research ^105^.

#### Target concentration

Bayesian dosing uses a ‘target concentration intervention’ ^10^ approach to maximise time within the acceptable concentration range. The concentration measure used as target in BRUNO-PIC will be to the average steady state concentration *(Cssavg)*, which is the AUC divided by the dosing interval, thus applicable at any steady-state dose interval. The acceptable range is 80-125% of the target *Cssavg*.

Table 1 details the *Ctrough* therapeutic ranges for month 1, 2 and 3 after kidney transplantation, and the equivalent AUC and *Cssavg* targets. The process for deriving *Cssavg* from *Ctrough* is expanded upon in the subsequent heading.

**Table 1:** Conversion from protocol Ctrough target to Cssavg. Abbreviations: AUC: area under curve; C: concentration; M:month; Tx: transplant.

| Kidney Tx | HCT non-standardized |  |  |  |  | HCT45 |  |  |
| --- | --- | --- | --- | --- | --- | --- | --- | --- |
| Post month | Ctrough low | Ctrough target | Ctrough high | Equiv. AUC<sub>0-12h</sub><sup>a</sup> | Equiv. *Cssavg*<sup>b</sup> | Trough target<sup>c</sup> | AUC<sub>0-12h</sub> target | *Cssavg* target<sup>c</sup> |
| M1 | 8 | 10 | 12 | 175 | 14.6 | 13.6 | 240 | 20 |
| M2 | 6 | 8 | 10 | 140 | 11.7 | 10.9 | 180 | 15 |
| M3 | 6 | 7 | 8 | 122.5 | 10.2 | 9 | 158 | 13.125 |
| <sup>a</sup>See below for determination AUC from Ctrough |  |  |  |  |  |  |  |  |
| <sup>b</sup>Equal to AUC0-12h/12 |  |  |  |  |  |  |  |  |
| <sup>c</sup>HCTnonstd multiplied by 45/33 (month 1) or 45/35 (month 2) |  |  |  |  |  |  |  |  |

#### Justification for tacrolimus concentration target

The *Cssavg* target for organ and post-transplant period is derived from the transplant unit-specific therapeutic window, with subsequent conversion. Thus, for a therapeutic window of 8-12 mcg/L, the *Csstrough* target is 10 mcg/L. This is then converted to a haematocrit-standardized AUC, then to *Cssavg* (AUC/12 given twice daily dosing).

To determine the equivalent exposure (AUC or *Cssavg*) at a given target *Csstrough*, we reviewed tacrolimus pharmacokinetic studies with both *Csstrough* and full AUC data. This enabled aggregate comparison of the relationship between the central tendency of the two parameters, using a subject number weighted average to determine typical AUC concentrations. This represents the exposure at steady state over the dosing interval (AUCssDI) seen in the average individual when dose is adjusted to target Csstrough, i.e. the AUCss expected with the measured trough concentration when approaching steady state in current practice. We also compared results with published recommendations for AUC targets.

Results are summarised in Table 2 below. The table first shows weighted average values (not standardised to HCT-45) indicating that a “raw” AUC_0-12_ value of 175.6 mcg/L.h is the typical exposure associated with a “raw” Ctrough of 10 mcg/L. This aligns well with published expert opinion (175 mcg/L.h being the mid-point of proposed window of 150-200 mcg/L.h) ^17^) and published precedence, with Meziyerh et al ^84^ describing 10-year experience of AUC-guided dosing in 968 adult kidney transplant recipients, with a goal tacrolimus AUC_0-12h_ goal of 160-180 mcg/L.h (mid-point 170 mcg/L.h).

**Table 2:** Literature values and weighted average Cssavg target. Abbreviations: AUC: area under curve; C: concentration; HCT: haematocrit; ss: steady state.

| Source | N° of profiles | Average Csstrough | Average AUC<sub>0-12</sub> | AUC<sub>0-12</sub> at C<sub>0</sub> of 10 mcg/L | Cssavg (HCT45) |
| --- | --- | --- | --- | --- | --- |
| Saint-Marcoux 2013<sup>69</sup> | 113 | 7.5 | 152.0 | 202.7 | 23.0 |
| Saint-Marcoux 2013<sup>69</sup> | 95 | 12.5 | 218.0 | 174.4 | 19.8 |
| Scholten 2005<sup>85</sup> | 33 | 11.9 | 181.0 | 152.1 | 17.3 |
| Braun 2001<sup>106</sup> | 20 | 9.3 | 159.3 | 171.3 | 19.5 |
| Kuypers 2004<sup>107</sup> | 100 | 10.3 | 168.5 | 163.6 | 18.6 |
| Kuypers 2004<sup>107</sup> | 100 | 9.0 | 154.7 | 171.9 | 19.5 |
| Włodarczyk 2009<sup>108</sup> | 32 | 10.1 | 180.7 | 178.6 | 20.3 |
| Włodarczyk 2009<sup>108</sup> | 32 | 10.0 | 171.8 | 171.5 | 19.5 |
| Włodarczyk 2009<sup>108</sup> | 32 | 12.1 | 191.3 | 158.6 | 18.0 |
| Niokka 2012<sup>109</sup> | 47 | 9.3 | 164.8 | 177.2 | 20.1 |
| Sum or weighted average | 604 | 10.2 | 174.2 | **175.6** | **14.64** |
| Weighted average, HCT45 |  |  |  | **239.5** | **19.96** |

Subsequently, the calculated AUCss was standardized to a haematocrit of 45%. Tacrolimus target concentrations are typically derived from studies using whole blood tacrolimus concentrations unstandardised for HCT. However, standardizing whole blood tacrolimus concentrations to a haematocrit of 45% (HCT45) account for changes in whole blood concentration with HCT at the same pharmacological active unbound tacrolimus concentration. The use of haematocrit-standardized tacrolimus concentrations has been shown to improve target attainment ^61–63^. The typical HCT seen in the early post-kidney transplant period of 33% ^61^ may be used to standardise literature value to a HCT of 45%. Finally, these HCT45 AUC values were divided by 12 to estimate the equivalent HCT45 *Cssavg*.

These steps and analysis, outlined in table below, gives a concentration target of 19.96 mcg/L hence proposing for BRUNO-PIC a target HCT45 *Cssavg* of 20 mcg/L.

### 2.4 Comparator

The prospective intervention arm will be compared with a retrospective arm from the prior 5 years, using standard of care standard of care dosing. This involves linear weight-based dosing (mg/kg) followed by TWA empiric dose adjustment to achieve *Csstrough* concentrations within a ‘therapeutic window’ ^10^.

### 2.5 Outcomes

#### Primary outcome

- The proportion of participants with tacrolimus concentration within 80-125% of concentration target (*Cssavg*) on post-transplant dosing DD4, Week 3 & Week 8, where *Cssavg* is calculated by dividing the associated dose by the individual Bayesian estimate of clearance and multiplying by the individual Bayesian estimate of bioavailability

#### Secondary outcomes

- Median time to acceptable range (80-125% *Cssavg*) in the immediate post-transplant period.
- Time within acceptable range (80-125% of *Cssavg*) in the first 8 weeks post-transplant
- Proportion of tacrolimus concentrations within 80-125% of Csstrough target on DD4
- Number of dose adjustments of tacrolimus based on TWA and/or Bayesian.
- Safety of genotype-informed Bayesian dosing, including description of number of clinical outcomes: rejection, donor-specific antibody formation; tacrolimus toxicities of new onset diabetes after transplantation.
- Feasibility of genotype-informed Bayesian dosing and barriers to implementation

#### Justification for outcome measures

As outlined in background, the excellent short-term outcomes from modern immunosuppression has challenge detecting superiority against hard outcomes with new interventions without very large numbers or longitudinal follow-up ^2^ ^3^, with academic and regulatory calls for innovative approaches including use of biomarkers as surrogates ^3^ ^70^. Robust data links tacrolimus underexposure and acute rejection, and reduced time within the acceptable range in the first 6 months post-transplant with negative short and long-term outcomes (rejection, *de novo* DSA formation, graft loss). In addition, these robust associations have a clear and causal mechanistic explanation. Thus, evidence for superior tacrolimus concentration control, with increased time in acceptable range, can reasonably be extrapolated to superior long-term outcomes.

### 2.6 Participant timeline

Eligible participants having undergone consent for BRUNO-PIC will have a blood sample sent for genotyping via a targeted gene panel looking at *CYP3A4* and *CYP3A5* as described in 3.1. A report is generated and made available at least 24 hours before transplant to be eligible to participate in the study. The assessments and data collection will be taken at set time points over an 8-week period post-transplantation (Table 3). The flow diagram for BRUNO-PIC study is shown in Figure 1.

**Figure 1:**
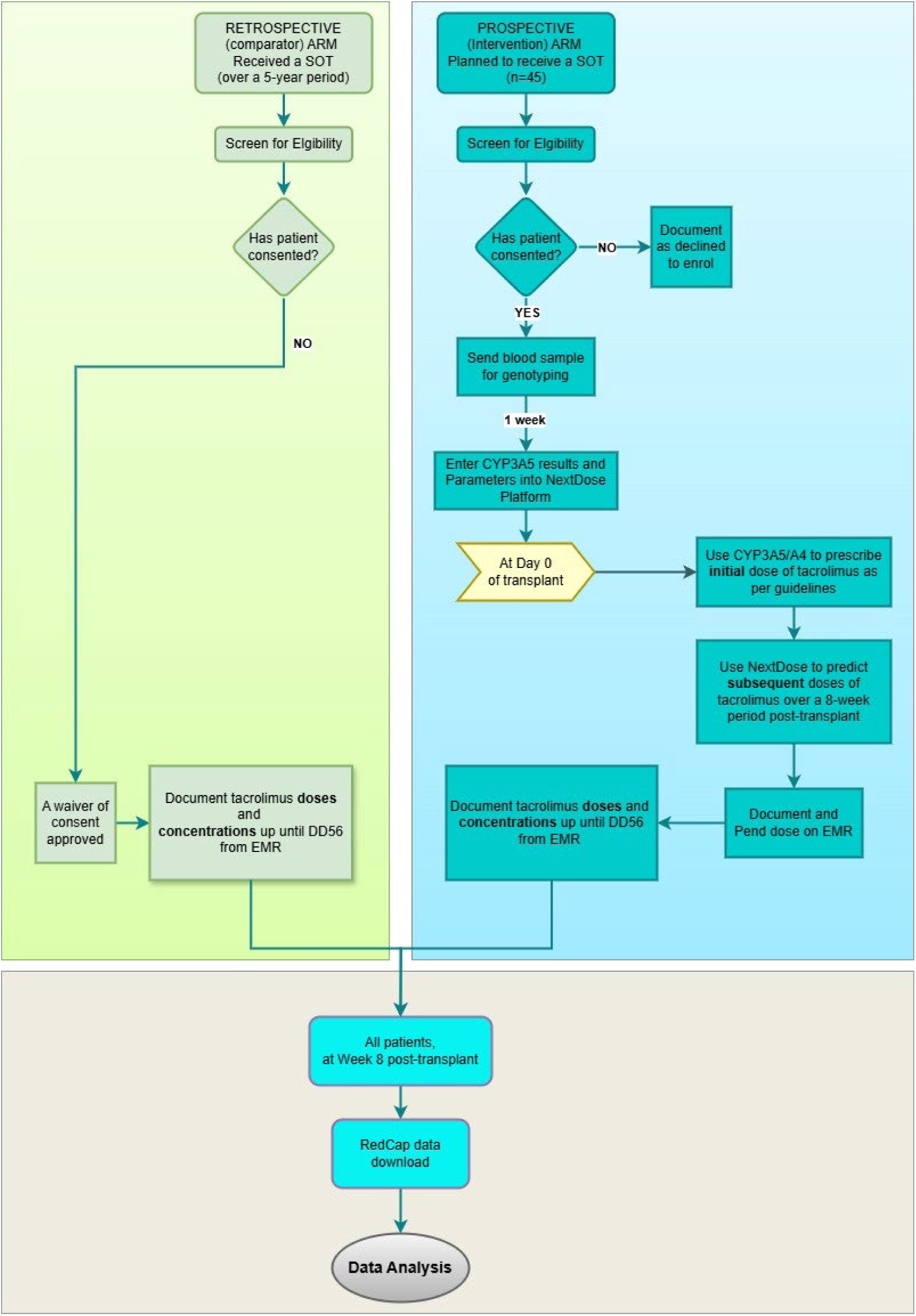
BRUNO-PIC study flow diagram. Abbreviations: CYP: cytochrome P450; DD0: Day of first tacrolimus dose where day 0 is the day of the first post-transplant dose; EMR: electronic medical record; SOT: solid-organ transplant. * For Retrospective arm: Tacrolimus doses, adjustments and concentrations will be taken from the EMR. If data is missing on D4, D21 or D56 timepoints, +2day (on D4) or ±7days (from D4 to D56) will be used. Any time points that are missing will be deemed as “lost to follow-up”. # For Prospective arm: Tacrolimus doses, adjustments and concentrations will be recorded from the EMR at set time points and as per transplant team policy.

**Table 3:**
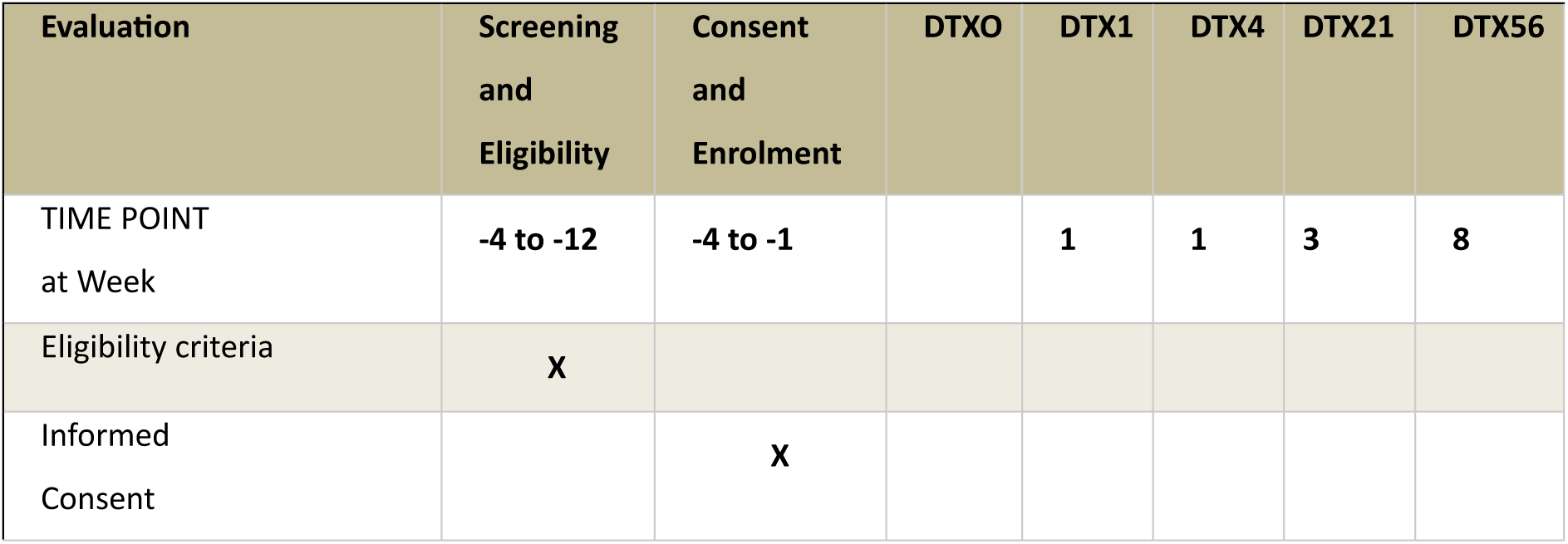
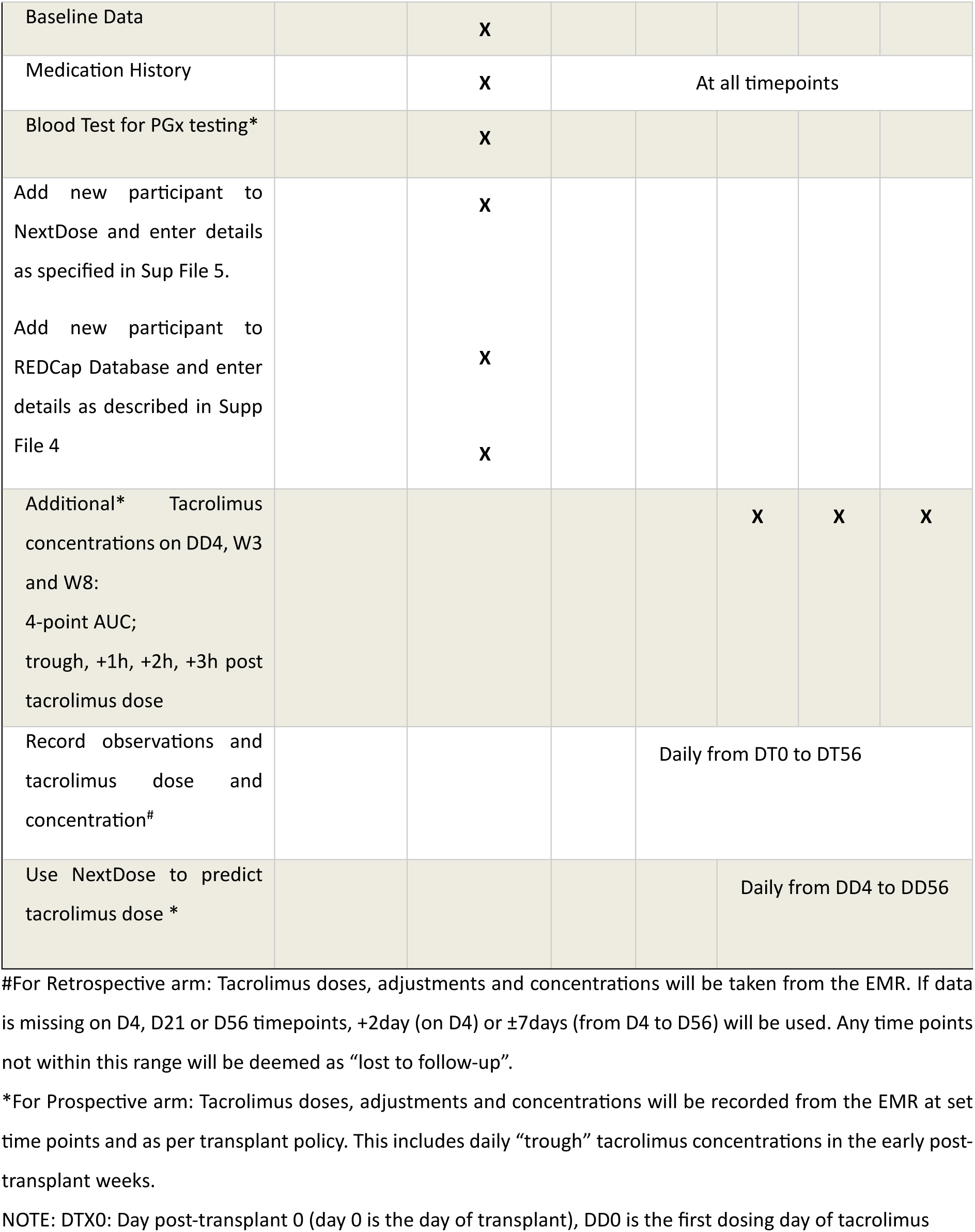
Overview of BRUNO-PIC study visits and timepoints.

| Evaluation | Screening and Eligibility | Consent and Enrolment | DTX0 | DTX1 | DTX4 | DTX21 | DTX56 |
| --- | --- | --- | --- | --- | --- | --- | --- |
| TIME POINT at Week | -4 to -12 | -4 to -1 |  | 1 | 1 | 3 | 8 |
| Eligibility criteria | X |  |  |  |  |  |  |
| Informed Consent |  | X |  |  |  |  |  |
| Baseline Data |  | X |  |  |  |  |  |
| Medication History |  | X | At all timepoints |  |  |  |  |
| Blood Test for PGx testing* |  | X |  |  |  |  |  |
| Add new participant to NextDose and enter details as specified in Sup File 5. |  | X |  |  |  |  |  |
| Add new participant to REDCap Database and enter details as described in Supp File 4 |  | X |  |  |  |  |  |
|  |  | X |  |  |  |  |  |
| Additional* Tacrolimus concentrations on DD4, W3 and W8:<br>4-point AUC; trough, +1h, +2h, +3h post tacrolimus dose |  |  |  |  | X | X | X |
| Record observations and tacrolimus dose and concentration<sup>#</sup> |  |  |  | Daily from DT0 to DT56 |  |  |  |
| Use NextDose to predict tacrolimus dose * |  |  |  |  | Daily from DD4 to DD56 |  |  |

#For Retrospective arm: Tacrolimus doses, adjustments and concentrations will be taken from the EMR. If data is missing on D4, D21 or D56 timepoints, +2day (on D4) or $\pm 7$ days (from D4 to D56) will be used. Any time points not within this range will be deemed as “lost to follow-up”.
\*For Prospective arm: Tacrolimus doses, adjustments and concentrations will be recorded from the EMR at set time points and as per transplant policy. This includes daily “trough” tacrolimus concentrations in the early post-transplant weeks.
NOTE: DTX0: Day post-transplant 0 (day 0 is the day of transplant), DD0 is the first dosing day of tacrolimus

### 2.4. Sample size

The sample size was calculated based on comparing proportion of participants within each group with a Ctrough concentration within acceptable range on day 4 post transplantation. Prior studies in adult kidney transplant recipients have shown 54% vs 24%^110^, 54.8% vs 20.8% ^63^ and 58% vs 37.4% ^23^, of patients with Ctrough within target range on Day 3-5 when treated with the standard, bodyweight-based dosing vs a dosing algorithm. We assumed 27% of the retrospective control cohort would have a tacrolimus Csstrough within acceptable range on DD4. 165 participants (120 control and 45 intervention participants) would provide 80% power to detect a risk difference of 24% assuming a two-sided alpha of 0.05.

### 2.5. Recruitment

Eligible participants will be screened in the outpatient or inpatient setting or through transplant planning meetings, held by the respective clinical departments, to identify those approaching need for kidney, liver or heart transplantation. Central transplant planning meetings help minimizing eligible participants being missed.

For the retrospective cohort, eligible individuals who underwent a SOT – liver, kidney, or heart – over a 5-year period will be extracted from the EMR.

## 3. METHODS: DATA COLLECTION, MANAGEMENT, AND ANALYSIS

### 3.1. Data collection

Study data from participants and electronic medical records will be entered de-identified into REDcap Case Report Forms. Study data collected for BRUNO-PIC is described in Supplemental File 4 along with databank guidelines. Although stored information will be de-identified it will be re-identifiable to delegated members of the study team for data entry and integrity purposes.

For Bayesian estimation, de-identified participant data will be entered into the NextDose server, securely hosted by the University of Otago, New Zealand.

### 3.2. Data management

Participants will be allocated a unique individual identifier prior to their de-identified data being entered into REDCap. All data is protected within Murdoch Children’s Research Institute (MCRI) using the REDCap database by a triple encryption process between the remote user, the server and the REDCap Database. No data can leave the database in an identifiable format.

### 3.3. Governance for Data Management and reporting

The Trial Monitoring Group consists of the Principal Investigators (RC and DM), academic Pharmacist/study coordinator (DK) and associate investigators (TS, JK, JM, EB, NH). The TMG will contribute to the organisation of Pharmacogenomic Steering Committee (PSC) meetings and will assess adverse events, serious adverse events and serious unexpected adverse events (SUSAR) with reporting in line with NHMRC standards. The TMG is also responsible for maintenance of the database, budget administration, HREC updates and amendments.

### 3.4. Statistics methods

#### Outcomes analysis

Comparison between the prospective intervention arm and the control arm (retrospective historical comparator) will be utilised for analyses where comparable data is available. A stabilised inverse probability weighting (IPW) approach will be used with robust standard errors to control for potential bias between “exposure” groups (i.e. NextDose (intervention) vs retrospective control cohort). IPW is an extension of the propensity score method used to summarise the conditional probability of assignment to an exposure. The weights are the inverse probability of assigning an exposure derived from a logistic model with group as the dependent variable and observed patient-level characteristics as the independent variables. These will include factors that may influence tacrolimus concentrations, including demographic variables (age, sex, height, weight), time after transplant, use of drugs known to interact with tacrolimus, presence of comorbidities known to influence transplant outcomes, presence of liver dysfunction, prednisolone daily dose in mg, and HCT levels (all measured on day of transplant D0).

We will stratify by type of solid organ transplant (heart, liver or kidney). Stabilisation is accomplished by multiplying the “exposure” weights (separately) by a constant, equal to the expected value of being in the intervention or control groups. Each participant is weighted by the inverse of the estimated probability of the exposure received. We will then use IPW regression models weighted with exposure to estimate the adjusted associations between exposure and each outcome.

The primary outcome analysis will compare the proportion of participants with a *Cssavg* concentration within acceptable range at DD4, week 3 and week 8. This will be using a risk difference estimated using a marginalised IPW mixed-effects logistic regression model including a random effect for the intercept (to allow for clustering of repeated measures within participants), and a fixed effect for dosing type (control vs intervention), time-period (DD4, week3, week8), SOT type (stratification factor) and other factors known to influence tacrolimus concentrations.

*Cssavg* will be calculated using a maximum *a posteriori* (MAP) approach, using drug concentrations and doses on days DD4, week3 and week8, along with participant clinical covariate information and the published popPK model within NextDose.

For secondary outcome, time to an acceptable *Cssavg* target, we will compare groups using a hazard ratio and 95% confidence interval estimated similarly to primary outcomes via a IPW adjusted Cox proportional hazards model. Between-group difference in mean percentage time within acceptable range over the first 8-weeks will be compared using IPW adjusted linear regression. For secondary outcomes where *Cssavg* is used, in the control group *Cssavg* will be calculated using measured concentrations and MAP estimation as above.

## 4. METHODS: MONITORING

### 4.1. Trial monitoring committee

Safety monitoring will be coordinated by the Trial Steering Committee, chaired by PIs and the study research assistant. Serious adverse events or drug reactions, and significant safety issues, are as defined in the National Health and Medical Research Council (NHMRC) Guidance: *Safety monitoring and reporting in clinical trials involving therapeutic goods* (2016). The TSC and broader research group meet fortnightly to oversee trial progress and review any adverse events. Reported adverse events will be assessed and reported as per NHMRC guidance using the trial Expedited Safety Report Form, Supplementary File 2.

## 5. ETHICS AND DISSEMINATION

### 5.1. Research ethics approval

BRUNO-PIC has been approved by the Sydney Children’s Ethics Committee (2023/ETH02699), and letter of authorisation from The Royal Children’s Hospital Research Governance Office (SSA/105019/RCHM-2024).

### 5.2. Protocol amendments

The study will be conducted in accordance with currently approved study protocol, with amendments to the protocol requiring HREC approval prior to implementation.

### 5.3. Consent

Eligible participants and their parents/caregivers will be introduced to the trial and if interested provided with the Participant Information Sheet and Consent Form (Supplementary Files 3). At subsequent hospital visit, a member of the trial team will discuss the trial in detail and provide sufficient time for questions prior to informed consent being attained.

Waiver of consent has been granted for the retrospective cohort by the approving ethics committee.

### 5.4. Confidentiality

Participant identity and clinical information, along with trial-related documents and data, will remian strictly confidential and accessible to delegated trial staff. No information concerning the trial, or the data will be released to any unauthorized third party, without prior written approval of the sponsoring institution. MCRI may inspect participant medical and pharmacy records with hospital consent. All evaluation forms will use Participant IDs for anonymity.

### 5.6. Dissemination policy

At the conclusion of the trial, participants will receive trial results and an overview of findings.

## Supporting information

Protocol

Spirit 2025 checklist

Data Information and Databank Guidelines

PICF

## Data Availability

All data produced in the present study are available upon reasonable request to the authors

## ACKNOWLEDGEMENTS

RC is supported by the Kids Cancer Project, The Royal Children’s Hospital Foundation, Victorian Paediatric Cancer Consortium, Medical Research Future Fund and holds a Murdoch Children’s Research Institute (MCRI) Clinician Scientist Tier 2 Fellowship and VESKI FAIR Fellowship. DK is supported by Medical Research Future Fund (MRF/2024900). CM is supported by a Melbourne University Research Training Program Strategic Scholarship. RSK is supported by a Fellowship from the National Health and Medical Research Council of Australia (APP2033152). The Chair in Genomic Medicine awarded to JC is generously supported by The Royal Children’s Hospital Foundation. Prof Nick Holford (NH) is the developer of NextDose, which is hosted by the University of Otago, New Zealand. NH assisted with protocol development and NextDose implementation.

## CONTRIBUTUTORS

DK, DM and RC wrote the main manuscript text; DK prepared the figures and DK and DM prepared the tables. All authors contributed to refinement of the study protocol and approved the final manuscript.

## DISCLAIMER

The funding source had no role in the design of this study and will not have any role during its execution, analysis, interpretation of the data, or decision to submit results.

## PATIENT CONSENT FOR PUBLICATION

Not applicable.

## 6.0 SUPPLEMENTARY

**Supplementary 1: SPIRIT checklist**

**Supplementary 2: Protocol**

**Supplementary 3: Informed Consent Forms (PICFS)-Participant and Parent/Guardian**

**Supplementary 4: Data Endpoint and Databank Guidelines**

## REFERENCES

1. Cooper JE. Evaluation and Treatment of Acute Rejection in Kidney Allografts. Clin J Am Soc Nephrol 2020;15(3):430–38. doi: 10.2215/cjn.11991019 [published Online First: 2020/02/19]

2. Neuberger JM, Bechstein WO, Kuypers DR, et al. Practical Recommendations for Long-term Management of Modifiable Risks in Kidney and Liver Transplant Recipients: A Guidance Report and Clinical Checklist by the Consensus on Managing Modifiable Risk in Transplantation (COMMIT) Group. Transplantation 2017;101(4S Suppl 2):S1–S56. doi: 10.1097/TP.0000000000001651

3. O’Connell PJ, Kuypers DR, Mannon RB, et al. Clinical Trials for Immunosuppression in Transplantation: The Case for Reform and Change in Direction. Transplantation 2017;101(7):1527–34. doi: 10.1097/tp.0000000000001648 [published Online First: 2017/02/17]

4. Nickerson PW, Rush DN. Begin at the Beginning to Prevent the End. J Am Soc Nephrol 2015;26(7):1483–5. doi: 10.1681/asn.2014111115 [published Online First: 2015/01/04]

5. Abdel-Kahaar E, Winter S, Tremmel R, et al. The impact of CYP3A4* 22 on tacrolimus pharmacokinetics and outcome in clinical practice at a single kidney transplant center. Frontiers in genetics 2019;10:871.

6. Ekberg H, Tedesco-Silva H, Demirbas A, et al. Reduced exposure to calcineurin inhibitors in renal transplantation. The New England journal of medicine 2007;357(25):2562–75. doi: 10.1056/NEJMoa067411 [published Online First: 2007/12/21]

7. Holford N, Petric Z. The Rational Basis for Personalized Treatment Using Concentration-Guided Dosing. Ther Drug Monit 2025;online [published Online First: 2025-06-10]

8. Riva N, Woillard JB, Distefano M, et al. Identification of Factors Affecting Tacrolimus Trough Levels in Latin American Pediatric Liver Transplant Patients. Liver Transpl 2019;25(9):1397–407. doi: 10.1002/lt.25495 [published Online First: 2019/05/19]

9. Barbarino JM, Staatz CE, Venkataramanan R, et al. PharmGKB summary: cyclosporine and tacrolimus pathways. Pharmacogenetics and genomics 2013;23(10):563.

10. Holford N, Ma G, Metz D. TDM is dead. Long live TCI! British Journal of Clinical Pharmacology 2022;88(4):1406–13. doi: 10.1111/bcp.14434

11. Israni AK, Riad SM, Leduc R, et al. Tacrolimus trough levels after month 3 as a predictor of acute rejection following kidney transplantation: a lesson learned from DeKAF Genomics. Transpl Int 2013;26(10):982–9. doi: 10.1111/tri.12155 [published Online First: 2013/07/25]

12. Wiebe C, Rush DN, Nevins TE, et al. Class II Eplet Mismatch Modulates Tacrolimus Trough Levels Required to Prevent Donor-Specific Antibody Development. Journal of the American Society of Nephrology : JASN 2017;28(11):3353–62. doi: 10.1681/ASN.2017030287 [published Online First: 2017/07/20]

13. Beland MA, Lapointe I, Noel R, et al. Higher calcineurin inhibitor levels predict better kidney graft survival in patients with de novo donor-specific anti-HLA antibodies: a cohort study. Transpl Int 2017;30(5):502–09. doi: 10.1111/tri.12934 [published Online First: 2017/02/12]

14. Wadström J, Ericzon B-G, Halloran PF, et al. Advancing Transplantation: New Questions, New Possibilities in Kidney and Liver Transplantation. Transplantation 2017;101(2):S1–S42. doi: 10.1097/tp.0000000000001563

15. Davis S, Gralla J, Klem P, et al. Lower tacrolimus exposure and time in therapeutic range increase the risk of de novo donor-specific antibodies in the first year of kidney transplantation. Am J Transplant 2018;18(4):907–15. doi: 10.1111/ajt.14504 [published Online First: 2017/09/20]

16. Girerd S, Schikowski J, Girerd N, et al. Impact of reduced exposure to calcineurin inhibitors on the development of de novo DSA: a cohort of non-immunized first kidney graft recipients between 2007 and 2014. BMC Nephrol 2018;19(1):232–32. doi: 10.1186/s12882-018-1014-2

17. Brunet M, van Gelder T, Åsberg A, et al. Therapeutic Drug Monitoring of Tacrolimus-Personalized Therapy: Second Consensus Report. Ther Drug Monit 2019;41(3):261–307. doi: 10.1097/ftd.0000000000000640 [published Online First: 2019/05/03]

18. Barraclough KA, Staatz CE, Johnson DW, et al. Kidney transplant outcomes are related to tacrolimus, mycophenolic acid and prednisolone exposure in the first week. Transplant International 2012;25(11):1182–93.

19. Hu R, Barratt DT, Coller JK, et al. Is There a Temporal Relationship Between Trough Whole Blood Tacrolimus Concentration and Acute Rejection in the First 14 Days After Kidney Transplantation? Therapeutic Drug Monitoring 2019;41(4)

20. Cheng F, Li Q, Cui Z, et al. Tacrolimus Concentration Is Effectively Predicted Using Combined Clinical and Genetic Factors in the Perioperative Period of Kidney Transplantation and Associated with Acute Rejection. J Immunol Res 2022;2022:3129389. doi: 10.1155/2022/3129389 [published Online First: 2022/09/20]

21. Nguyen TVA, Nguyen HD, Nguyen TLH, et al. Higher tacrolimus trough levels and time in the therapeutic range are associated with the risk of acute rejection in the first month after renal transplantation. BMC Nephrol 2023;24(1) doi: 10.1186/s12882-023-03188-0

22. Jennings DL, Salerno D, Lange N, et al. Faster Time-to-Therapeutic Tacrolimus Level is Associated with Lower Risk of Cellular Rejection Early after Heart Transplantation. The Journal of Heart and Lung Transplantation 2020;39(4):S502. doi: 10.1016/j.healun.2020.01.101

23. Francke MI, Andrews LM, Le HL, et al. Avoiding Tacrolimus Underexposure and Overexposure with a Dosing Algorithm for Renal Transplant Recipients: A Single Arm Prospective Intervention Trial. Clin Pharmacol Ther 2021;110(1):169–78. doi: 10.1002/cpt.2163 [published Online First: 2021/01/17]

24. Yang H, Sun Y, Yu X, et al. Clinical Impact of the Adaptation of Initial Tacrolimus Dosing to the CYP3A5 Genotype After Kidney Transplantation: Systematic Review and Meta-Analysis of Randomized Controlled Trials. Clin Pharmacokinet 2021;60(7):877–85. doi: 10.1007/s40262-020-00955-2 [published Online First: 2021/03/23]

25. Lloberas N, Grinyó JM, Colom H, et al. A prospective controlled, randomized clinical trial of kidney transplant recipients developed personalized tacrolimus dosing using model-based Bayesian Prediction. Kidney Int 2023;104(4):840–50. doi: 10.1016/j.kint.2023.06.021 [published Online First: 2023/07/01]

26. Anderson BJ, Holford NH. Understanding dosing: children are small adults, neonates are immature children. Archives of disease in childhood 2013;98(9):737–44. doi: 10.1136/archdischild-2013-303720

27. Min S, Papaz T, Lafreniere-Roula M, et al. A randomized clinical trial of age and genotype-guided tacrolimus dosing after pediatric solid organ transplantation. Pediatr Transplant 2018;22(7):e13285. doi: 10.1111/petr.13285 [published Online First: 2018/09/05]

28. Kausman JY, Patel B, Marks SD. Standard dosing of tacrolimus leads to overexposure in pediatric renal transplantation recipients. Pediatric transplantation 2008;12(3):329–35. doi: 10.1111/j.1399-3046.2007.00821.x [published Online First: 2008/04/26]

29. Holford NHG, Anderson BJ. Allometric size: The scientific theory and extension to normal fat mass. Eur J Pharm Sci 2017;109s:S59–s64. doi: 10.1016/j.ejps.2017.05.056 [published Online First: 20170525]

30. Staatz CE, Tett SE. Clinical pharmacokinetics and pharmacodynamics of tacrolimus in solid organ transplantation. Clin Pharmacokinet 2004;43(10):623–53. doi: 10.2165/00003088-200443100-00001 [published Online First: 2004/07/13]

31. Birdwell KA, Decker B, Barbarino JM, et al. Clinical Pharmacogenetics Implementation Consortium (CPIC) Guidelines for CYP3A5 Genotype and Tacrolimus Dosing. Clin Pharmacol Ther 2015;98(1):19–24. doi: 10.1002/cpt.113 [published Online First: 2015/03/25]

32. Aranda JavdA, JN. Yaffe and Aranda’s Neonatal and Pediatric Pharmacology: Therapeutic Principles in Practice. 5 ed. Lippincott Williams & Wilkins Wolters Kluwer 2021.

33. van Gelder T, Meziyerh S, Swen JJ, et al. The Clinical Impact of the C0/D Ratio and the CYP3A5 Genotype on Outcome in Tacrolimus Treated Kidney Transplant Recipients. Frontiers in Pharmacology 2020;11(1142) doi: 10.3389/fphar.2020.01142

34. Abdullah-Koolmees H, van Keulen AM, Nijenhuis M, et al. Pharmacogenetics Guidelines: Overview and Comparison of the DPWG, CPIC, CPNDS, and RNPGx Guidelines. Front Pharmacol 2020;11:595219. doi: 10.3389/fphar.2020.595219 [published Online First: 2021/02/12]

35. Kim JS, Shim S, Yee J, et al. Effects of CYP3A4*22 polymorphism on trough concentration of tacrolimus in kidney transplantation: a systematic review and meta-analysis. Frontiers in pharmacology 2023;Volume 14 - 2023 doi: 10.3389/fphar.2023.1201083

36. Woillard JB, Mourad M, Neely M, et al. Tacrolimus Updated Guidelines through popPK Modeling: How to Benefit More from CYP3A Pre-emptive Genotyping Prior to Kidney Transplantation. Front Pharmacol 2017;8:358. doi: 10.3389/fphar.2017.00358 [published Online First: 20170608]

37. Li D, Lu W, Zhu JY, et al. Population pharmacokinetics of tacrolimus and CYP3A5, MDR1 and IL-10 polymorphisms in adult liver transplant patients. J Clin Pharm Ther 2007;32(5):505–15. doi: 10.1111/j.1365-2710.2007.00850.x

38. Moes DJ, van der Bent SA, Swen JJ, et al. Population pharmacokinetics and pharmacogenetics of once daily tacrolimus formulation in stable liver transplant recipients. Eur J Clin Pharmacol 2016;72(2):163–74. doi: 10.1007/s00228-015-1963-3 [published Online First: 20151031]

39. Huang L, Assiri AA, Wen P, et al. The CYP3A5 genotypes of both liver transplant recipients and donors influence the time-dependent recovery of tacrolimus clearance during the early stage following transplantation. Clin Transl Med 2021;11(10):e542. doi: 10.1002/ctm2.542

40. Shao J, Wang C, Fu P, et al. Impact of Donor and Recipient CYP3A5*3 Genotype on Tacrolimus Population Pharmacokinetics in Chinese Adult Liver Transplant Recipients. Ann Pharmacother 2020;54(7):652–61. doi: 10.1177/1060028019897050 [published Online First: 20191230]

41. Ji E, Kim MG, Oh JM. CYP3A5 genotype-based model to predict tacrolimus dosage in the early postoperative period after living donor liver transplantation. Ther Clin Risk Manag 2018;14:2119–26. doi: 10.2147/TCRM.S184376 [published Online First: 20181025]

42. Liu J, Man K. Mechanistic Insight and Clinical Implications of Ischemia/Reperfusion Injury Post Liver Transplantation. Cellular and Molecular Gastroenterology and Hepatology 2023;15(6):1463–74. doi: 10.1016/j.jcmgh.2023.03.003

43. Ghonem N, Yoshida J, Murase N, et al. Treprostinil Improves Hepatic Cytochrome P450 Activity during Rat Liver Transplantation. Journal of Clinical and Experimental Hepatology 2012;2(4):323–32. doi: 10.1016/j.jceh.2012.09.002

44. Guo Y, Hu B, Xie Y, et al. Regulation of drug-metabolizing enzymes by local and systemic liver injuries. Expert Opin Drug Metab Toxicol 2016;12(3):245–51. doi: 10.1517/17425255.2016.1139574 [published Online First: 20160128]

45. Thörn M, Lundgren S, Herlenius G, et al. Gene expression of cytochromes P450 in liver transplants over time. Eur J Clin Pharmacol 2004;60(6):413–20. doi: 10.1007/s00228-004-0786-4 [published Online First: 20040610]

46. Aksoy F, Gurluler E, Celik F, et al. Evaluation of the Trend of Biochemical Functions in the Early Period After Cadaveric Liver Transplantation. Transplant Proc 2023;55(5):1252–56. doi: 10.1016/j.transproceed.2023.03.040 [published Online First: 20230425]

47. Tannuri U, Tannuri ACA. Postoperative care in pediatric liver transplantation. Clinics 2014;69:42–46. doi: 10.6061/clinics/2014(Sup01)08

48. Dong Y, Xu Q, Li R, et al. CYP3A7, CYP3A4, and CYP3A5 genetic polymorphisms in recipients rather than donors influence tacrolimus concentrations in the early stages after liver transplantation. Gene 2022;809:146007.

49. Leino AD, Park JM, Pasternak AL. Impact of CYP3A5 phenotype on tacrolimus time in therapeutic range and clinical outcomes in pediatric renal and heart transplant recipients. Pharmacotherapy 2021;41(8):649–57. doi: 10.1002/phar.2601 [published Online First: 2021/06/16]

50. Song W, Lao Q, Hu J, et al. Lower tacrolimus time in therapeutic range is associated with inferior outcomes in adult liver transplant recipients. Basic Clin Pharma Tox 2022;132(1):51–59. doi: 10.1111/bcpt.13803

51. Katada Y, Nakagawa S, Itohara K, et al. Association between time in therapeutic range of tacrolimus blood conc entration and acute rejection within the first three months after lung transplantation. J Pharm Health Care Sci 2022;8(1) doi: 10.1186/s40780-022-00256-9

52. Davis S, Gralla J, Klem P, et al. Tacrolimus Intrapatient Variability, Time in Therapeutic Range, and Risk of De Novo Donor-Specific Antibodies. Transplantation 2020;104(4):881–87. doi: 10.1097/tp.0000000000002913 [published Online First: 2020/04/01]

53. Lichtenberg S, Rahamimov R, Green H, et al. The incidence of post-transplant cancer among kidney transplant recipients is associated with the level of tacrolimus exposure during the first year after transplantation. Eur J Clin Pharmacol 2017;73(7):819–26. doi: 10.1007/s00228-017-2234-2 [published Online First: 2017/03/28]

54. Barreda P, Cañamero L, Boya M, et al. Lower Time in Therapeutic Range Relates to a Worse Kidney Graft Outcom e. Transplantation Proceedings 2022;54(9):2446–49. doi: 10.1016/j.transproceed.2022.09.013

55. Han A, Jo AJ, Kwon H, et al. Optimum tacrolimus trough levels for enhanced graft survival and safet y in kidney transplantation: a retrospective multicenter real-world ev idence study. International Journal of Surgery 2024;110(10):6711–22. doi: 10.1097/js9.0000000000001800

56. Yin S, Huang Z, Wang Z, et al. Early Monitoring and Subsequent Gain of Tacrolimus Time-In-Therapeutic Range May Improve Clinical Outcomes After Living Kidney Transplantati on. Therapeutic Drug Monitoring 2021;43(6):728–35. doi: 10.1097/ftd.0000000000000881

57. Ensor CR, Iasella CJ, Harrigan KM, et al. Increasing tacrolimus time-in-therapeutic range is associated with sup erior one-year outcomes in lung transplant recipients. American Journal of Transplantation 2018;18(6):1527–33. doi: 10.1111/ajt.14723

58. Song T, Yin S, Jiang Y, et al. Increasing Time in Therapeutic Range of Tacrolimus in the First Year Predicts Better Outcomes in Living-Donor Kidney Transplantation. Front Immunol 2019;10:2912. doi: 10.3389/fimmu.2019.02912 [published Online First: 20191220]

59. Woillard JB, Saint-Marcoux F, Debord J, et al. Pharmacokinetic models to assist the prescriber in choosing the best tacrolimus dose. Pharmacol Res 2018;130:316–21. doi: 10.1016/j.phrs.2018.02.016 [published Online First: 2018/02/17]

60. Ekberg H, Mamelok RD, Pearson TC, et al. The challenge of achieving target drug concentrations in clinical trials: experience from the Symphony study. Transplantation 2009;87(9):1360–6. doi: 10.1097/TP.0b013e3181a23cb2 [published Online First: 2009/05/09]

61. Storset E, Holford N, Hennig S, et al. Improved prediction of tacrolimus concentrations early after kidney transplantation using theory-based pharmacokinetic modelling. Br J Clin Pharmacol 2014;78(3):509–23. doi: 10.1111/bcp.12361

62. Størset E, Åsberg A, Skauby M, et al. Improved Tacrolimus Target Concentration Achievement Using Computerized Dosing in Renal Transplant Recipients--A Prospective, Randomized Study. Transplantation 2015;99(10):2158–66. doi: 10.1097/tp.0000000000000708 [published Online First: 2015/04/19]

63. Lloberas N, Grinyo JM, Colom H, et al. A prospective controlled, randomized clinical trial of kidney transplant recipients developed personalized tacrolimus dosing using model-based Bayesian Prediction. Kidney Int 2023 doi: 10.1016/j.kint.2023.06.021 [published Online First: 20230628]

64. Marquet P, Cros F, Micallef L, et al. Tacrolimus Bayesian Dose Adjustment in Pediatric Renal Transplant Recipients. Ther Drug Monit 2021;43(4):472–80. doi: 10.1097/ftd.0000000000000828 [published Online First: 2020/11/06]

65. Woillard JB, Debord J, Monchaud C, et al. Population Pharmacokinetics and Bayesian Estimators for Refined Dose Adjustment of a New Tacrolimus Formulation in Kidney and Liver Transplant Patients. Clin Pharmacokinet 2017;56(12):1491–98. doi: 10.1007/s40262-017-0533-5 [published Online First: 2017/04/09]

66. Neely M, Jelliffe R. Practical, individualized dosing: 21st century therapeutics and the clinical pharmacometrician. J Clin Pharmacol 2010;50(7):842–7. doi: 10.1177/0091270009356572 [published Online First: 2010/02/16]

67. Darwich AS, Polasek TM, Aronson JK, et al. Model-Informed Precision Dosing: Background, Requirements, Validation, Implementation, and Forward Trajectory of Individualizing Drug Therapy. Annual Review of Pharmacology and Toxicology 2021;61(1):225–45. doi: 10.1146/annurev-pharmtox-033020-113257

68. TGA. Consultation: Proposed clarification of how Clinical Decision Support System software is regulated 2024 [cited 2025 15/04/2025].

69. Saint-Marcoux F, Woillard JB, Jurado C, et al. Lessons from routine dose adjustment of tacrolimus in renal transplant patients based on global exposure. Ther Drug Monit 2013;35(3):322–7. doi: 10.1097/FTD.0b013e318285e779

70. FDA. Endpoints and trial designs to advance drug development in kidney transplantation 2023 [Available from: https://www.fda.gov/drugs/news-events-human-drugs/endpoints-and-trial-designs-advance-drug-development-kidney-transplantation-11092023 accessed March 11 2025.

71. Xue L, Ma G, Holford N, et al. A Randomized Trial Comparing Standard of Care to Bayesian Warfarin Dose Individualization. Clin Pharmacol Ther 2024;115(6):1316–25. doi: 10.1002/cpt.3207 [published Online First: 20240304]

72. Zhao CY, Jiao Z, Mao JJ, et al. External evaluation of published population pharmacokinetic models of tacrolimus in adult renal transplant recipients. Br J Clin Pharmacol 2016;81(5):891–907. doi: 10.1111/bcp.12830 [published Online First: 2015/11/18]

73. Aarons L. Physiologically based pharmacokinetic modelling: a sound mechanistic basis is needed. Br J Clin Pharmacol 2005;60(6):581–3. doi: 10.1111/j.1365-2125.2005.02560.x [published Online First: 2005/11/25]

74. Germovsek E, Barker CI, Sharland M, et al. Scaling clearance in paediatric pharmacokinetics: All models are wrong, which are useful? Br J Clin Pharmacol 2017;83(4):777–90. doi: 10.1111/bcp.13160 [published Online First: 20161202]

75. Nanga TM, Doan TTP, Marquet P, et al. Toward a robust tool for pharmacokinetic-based personalization of treatment with tacrolimus in solid organ transplantation: A model-based meta-analysis approach. Br J Clin Pharmacol 2019;85(12):2793–823. doi: 10.1111/bcp.14110 [published Online First: 20191217]

76. Itohara K, Yano I, Nakagawa S, et al. Extrapolation of physiologically based pharmacokinetic model for tacrolimus from renal to liver transplant patients. Drug Metab Pharmacokinet 2022;42:100423. doi: 10.1016/j.dmpk.2021.100423 [published Online First: 20211001]

77. Paschier A, Destere A, Monchaud C, et al. Tacrolimus population pharmacokinetics in adult heart transplant patients. Br J Clin Pharmacol 2023;89(12):3584–95. doi: 10.1111/bcp.15857

78. Woillard J-B, de Winter BCM, Kamar N, et al. Population pharmacokinetic model and Bayesian estimator for two tacrolimus formulations – twice daily Prograf® and once daily Advagraf®. Br J Clin Pharmacol 2011;71(3):391–402. doi: 10.1111/j.1365-2125.2010.03837.x

79. van Gelder T, Gelinck A, Meziyerh S, et al. Therapeutic drug monitoring of tacrolimus after kidney transplantation: trough concentration or area under curve-based monitoring? Br J Clin Pharmacol 2025;91(6):1600–06. doi: 10.1111/bcp.16098

80. Undre NA. Pharmacokinetics of tacrolimus-based combination therapies. Nephrol Dial Transplant 2003;18 Suppl 1:i12–5. doi: 10.1093/ndt/gfg1029 [published Online First: 2003/05/10]

81. Mahalati K, Belitsky P, Sketris I, et al. Neoral monitoring by simplified sparse sampling area under the concentration-time curve: its relationship to acute rejection and cyclosporine nephrotoxicity early after kidney transplantation. Transplantation 1999;68(1):55–62. doi: 10.1097/00007890-199907150-00011 [published Online First: 1999/07/31]

82. Undre NA, van Hooff J, Christiaans M, et al. Low systemic exposure to tacrolimus correlates with acute rejection. Transplant Proc 1999;31(1-2):296–8. doi: 10.1016/s0041-1345(98)01633-9 [published Online First: 1999/03/20]

83. van Rossum HH, Press RR, den Hartigh J, et al. Point: A call for advanced pharmacokinetic and pharmacodynamic monitoring to guide calcineurin inhibitor dosing in renal transplant recipients. Clinical chemistry 2010;56(5):732–5. doi: 10.1373/clinchem.2009.141135 [published Online First: 2010/03/09]

84. Meziyerh S, van Gelder T, Kers J, et al. Tacrolimus and Mycophenolic Acid Exposure Are Associated with Biopsy-Proven Acute Rejection: A Study to Provide Evidence for Longer-Term Target Ranges. Clinical Pharmacology & Therapeutics 2023;114(1):192–200. doi: 10.1002/cpt.2915

85. Scholten EM, Cremers SC, Schoemaker RC, et al. AUC-guided dosing of tacrolimus prevents progressive systemic overexposure in renal transplant recipients. Kidney Int 2005;67(6):2440–7. doi: 10.1111/j.1523-1755.2005.00352.x [published Online First: 2005/05/11]

86. Villeneuve C, Humeau A, Monchaud C, et al. Better Rejection-Free Survival at Three Years in Kidney Transplant Recipients With Model-Informed Precision Dosing of Mycophenolate Mofetil. Clin Pharmacol Ther 2024;116(2):351–62. doi: 10.1002/cpt.3206 [published Online First: 20240219]

87. Haverals L, Roosens L, Wouters K, et al. Does the Tacrolimus Trough Level Adequately Predict Drug Exposure in Patients Requiring a High Tacrolimus Dose? Transplant Direct 2023;9(4):e1439. doi: 10.1097/TXD.0000000000001439 [published Online First: 20230329]

88. Woillard JB, Monchaud C, Saint-Marcoux F, et al. Can the Area Under the Curve/Trough Level Ratio Be Used to Optimize Tacrolimus Individual Dose Adjustment? Transplantation 2023;107(1):e27–e35. doi: 10.1097/tp.0000000000004405 [published Online First: 2022/12/13]

89. Rojas L, Neumann I, Herrero MJ, et al. Effect of CYP3A5*3 on kidney transplant recipients treated with tacrolimus: a systematic review and meta-analysis of observational studies. The pharmacogenomics journal 2015;15(1):38–48. doi: 10.1038/tpj.2014.38 [published Online First: 2014/09/10]

90. Schutte-Nutgen K, Tholking G, Steinke J, et al. Fast Tac Metabolizers at Risk (-) It is Time for a C/D Ratio Calculation. J Clin Med 2019;8(5) doi: 10.3390/jcm8050587 [published Online First: 2019/05/01]

91. Trofe-Clark J, Brennan DC, West-Thielke P, et al. Results of ASERTAA, a Randomized Prospective Crossover Pharmacogenetic Study of Immediate-Release Versus Extended-Release Tacrolimus in African American Kidney Transplant Recipients. Am J Kidney Dis 2018;71(3):315–26. doi: 10.1053/j.ajkd.2017.07.018 [published Online First: 2017/11/23]

92. Tholking G, Siats L, Fortmann C, et al. Tacrolimus Concentration/Dose Ratio is Associated with Renal Function After Liver Transplantation. Annals of transplantation 2016;21:167–79. doi: 10.12659/aot.895898 [published Online First: 2016/03/24]

93. Tholking G, Fortmann C, Koch R, et al. The tacrolimus metabolism rate influences renal function after kidney transplantation. PLoS One 2014;9(10):e111128. doi: 10.1371/journal.pone.0111128 [published Online First: 2014/10/24]

94. Egeland EJ, Reisaeter AV, Robertsen I, et al. High tacrolimus clearance - a risk factor for development of interstitial fibrosis and tubular atrophy in the transplanted kidney: a retrospective single-center cohort study. Transpl Int 2019;32(3):257–69. doi: 10.1111/tri.13356 [published Online First: 2018/09/27]

95. Kwiatkowska E, Kwiatkowski S, Wahler F, et al. C/D Ratio in Long-Term Renal Function. Transplant Proc 2019;51(10):3265–70. doi: 10.1016/j.transproceed.2019.08.030 [published Online First: 20191113]

96. Chamoun B, Torres IB, Gabaldón A, et al. Progression of Interstitial Fibrosis and Tubular Atrophy in Low Immuno logical Risk Renal Transplants Monitored by Sequential Surveillance Bi opsies: The Influence of TAC Exposure and Metabolism. JCM 2021;10(1):141. doi: 10.3390/jcm10010141

97. Tholking G, Schmidt C, Koch R, et al. Influence of tacrolimus metabolism rate on BKV infection after kidney transplantation. Sci Rep 2016;6:32273. doi: 10.1038/srep32273 [published Online First: 2016/08/31]

98. Thölking G, Tosun-Koç F, Jehn U, et al. Improved Kidney Allograft Function after Early Conversion of Fast IR-Tac Metabolizers to LCP-Tac. J Clin Med 2022;11(5) doi: 10.3390/jcm11051290 [published Online First: 2022/03/11]

99. Langone A, Steinberg SM, Gedaly R, et al. Switching STudy of Kidney TRansplant PAtients with Tremor to LCP-TacrO (STRATO): an open-label, multicenter, prospective phase 3b study. Clin Transplant 2015;29(9):796–805. doi: 10.1111/ctr.12581 [published Online First: 2015/06/27]

100. Taber DJ, Gebregziabher MG, Srinivas TR, et al. African-American race modifies the influence of tacrolimus concentrations on acute rejection and toxicity in kidney transplant recipients. Pharmacotherapy 2015;35(6):569–77. doi: 10.1002/phar.1591 [published Online First: 2015/05/27]

101. Ting LS, Villeneuve E, Ensom MH. Beyond cyclosporine: a systematic review of limited sampling strategies for other immunosuppressants. Ther Drug Monit 2006;28(3):419–30. doi: 10.1097/01.ftd.0000211810.19935.44 [published Online First: 2006/06/17]

102. Pallet N, Jannot AS, El Bahri M, et al. Kidney transplant recipients carrying the CYP3A4*22 allelic variant have reduced tacrolimus clearance and often reach supratherapeutic tacrolimus concentrations. Am J Transplant 2015;15(3):800–5. doi: 10.1111/ajt.13059 [published Online First: 20150114]

103. Paschier A, Destere A, Monchaud C, et al. Tacrolimus population pharmacokinetics in adult heart transplant patients. Br J Clin Pharmacol 2023;89(12):3584–95. doi: 10.1111/bcp.15857 [published Online First: 20230810]

104. Textor SC, Wiesner R, Wilson DJ, et al. Systemic and renal hemodynamic differences between FK506 and cyclosporine in liver transplant recipients. Transplantation 1993;55(6):1332–9. doi: 10.1097/00007890-199306000-00023 [published Online First: 1993/06/01]

105. Peplow C, Assfalg R, Beyerlein A, et al. Blood draws up to 3% of blood volume in clinical trials are safe in children. Acta Paediatr 2019;108(5):940–44. doi: 10.1111/apa.14607 [published Online First: 2018/10/07]

106. Braun F, Schütz E, Peters B, et al. Pharmacokinetics of tacrolimus primary immunosuppression in kidney transplant recipients. Transplant Proc 2001;33(3):2127–8. doi: 10.1016/s0041-1345(01)01970-4 [published Online First: 2001/05/30]

107. Kuypers DR, Claes K, Evenepoel P, et al. Clinical efficacy and toxicity profile of tacrolimus and mycophenolic acid in relation to combined long-term pharmacokinetics in de novo renal allograft recipients. Clin Pharmacol Ther 2004;75(5):434–47. doi: 10.1016/j.clpt.2003.12.009 [published Online First: 2004/04/30]

108. Wlodarczyk Z, Squifflet JP, Ostrowski M, et al. Pharmacokinetics for once-versus twice-daily tacrolimus formulations in de novo kidney transplantation: a randomized, open-label trial. Am J Transplant 2009;9(11):2505–13. doi: 10.1111/j.1600-6143.2009.02794.x [published Online First: 2009/08/18]

109. Niioka T, Satoh S, Kagaya H, et al. Comparison of pharmacokinetics and pharmacogenetics of once- and twice-daily tacrolimus in the early stage after renal transplantation. Transplantation 2012;94(10):1013–9. doi: 10.1097/TP.0b013e31826bc400 [published Online First: 2012/10/18]

110. Raj TY, Fernando ME, Srinivasa Prasad ND, et al. Efficacy and Outcomes of CYP3A5 Genotype-Based Tacrolimus Dosing Compared to Conventional Body Weight-based Dosing in Living Donor Kidney Transplant Recipients. Indian J Nephrol 2022;32(3):240–46. doi: 10.4103/ijn.IJN_278_20 [published Online First: 20220507]

