## Supplementary material for "Protocol for Bayesian combined multi-genotype and concentration informed tacrolimus dosing in paediatric solid organ transplantation (BRUNO-PIC)": Protocol

### BRUNO – PIC:

*Genotype informed **B**ayesian dosing of tac**r**olim**u**s i**n** solid **o**rgan transplant-*
***P**harmacogenomic **I**mplementation in **C**hildren*

### Version 4.0, 26 May 2025
### HREC: 2023/ETH02699

**CONFIDENTIAL**
This protocol is confidential and is the property of Murdoch Children's Research Institute. No part of it may be transmitted, reproduced, published, or used without prior written authorisation from the institution.

**Statement of Compliance**
This clinical trial will be conducted in compliance with all stipulation of this protocol, the conditions of the ethics committee approval, the NHMRC National Statement on ethical Conduct in Human Research (2007 and all updates), the Integrated Addendum to ICH E6 (R1): Guideline for Good Clinical Practice E6 (R2), dated 9 November 2016 annotated with TGA comments and the NHMRC guidance Safety monitoring and reporting in clinical trials involving therapeutic goods (EH59, 2016).

This clinical trial is not sponsored by any pharmaceutical company or other commercial entity.

**REVISION CHRONOLOGY:**

| **Version Number & Date** | **Summary of changes** |
|---|---|
| 2.0 dated 23rd July 2024 | Removed AI, added two AIs, updated RCH HREC to SCH HREC, Amended Dr to Prof, changed additional samples to be taken at 3 hours instead of 4 hours post tacrolimus |
| 3.0 dated 8 Sept 2024 | Increased detail and adjustment to co-primary outcomes.<br>Change of timing of additional tacrolimus samples beyond SOC.<br>- Limited (4-point) PK profile taken D4, W3 and W8, along with removal of the 8-hourly sampling D3 and 4 post transplants.<br>- This still equates to no additional venipuncture beyond standard of care, with sampling taken either from CVAD (on D4) or during AUC estimation for mycophenolic acid (part of standard of care).<br>Expansion of relevant scientific background in appendix on the use of genotype (CYP3A4 & CYP3A5), allometric scaling, haematocrit correction, Cavgss and the basis for targets used. These provide additional detail, though do not change the approach from prior ethically approved protocol. |
| 4.0 dated 26 May 2025 | Added a new AI<br>Updated Background information and formatting<br>Increased detail on justification for initial dose; target concentration for tacrolimus. |

| | |
|---|---|
| | Increased detail on pre-validation of NextDose use in cardiac and liver transplant recipients.<br>Updated Retrospective cohort to include all transplanted patients over a 5-year period<br>Updated the SAP and made outcomes clearer<br>Added a new appendix on how to use the Nextdose for dose prediction |

**CONTENTS**

CONTENTS ... 3
PROTOCOL SYNOPSIS ... 6
INVESTIGATOR AGREEMENT ... 14
1. ADMINISTRATIVE INFORMATION ... 15
2. BACKGROUND & RATIONALE ... 16
2.1 Overview ... 16
2.2 Background ... 16
2.2.1 Calcineurin inhibitors in SOT – history & current practice ... 16
2.2.2 Limitations of current dosing and proposed intervention ... 19
2.2.3 NextDose, and applicability to paediatric SOT cohort ... 23
2.3 Summary ... 25
2.4 Risk/Benefit assessment ... 25
2.4.1 Potential benefits. ... 25
2.4.2 Potential risks. ... 26
3. TRIAL OBJECTIVES AND ENDPOINTS ... 26
3.1 Objectives ... 26
3.1.1 Primary objective ... 26
3.1.2 Secondary objectives ... 26
3.2 Endpoints ... 26
3.2.1 Primary endpoint: ... 27
3.2.1 Secondary endpoints ... 27
3.3 Justification for endpoints ... 27
4 TRIAL DESIGN ... 27
4.1 Overall design ... 27
4.2 Justification for dose and target concentration ... 28
4.2.1 Initial dosing (see APPENDIX 2 for details) ... 28
4.2.2 Subsequent dosing (See APPENDIX 3 for details) ... 28
5 MATERIALS AND METHODS ... 29
5.1 Subjects ... 29
5.1.1 Number of subjects ... 30
5.1.2 Inclusion criteria ... 30
5.1.3 Exclusion criteria ... 30

5.1.4 Subject enrollment (phased) ... 30
5.1.5 Recruitment & consent ... 31
5.2 Intervention ... 33
5.2.1 Pre-emptive CYP3A4/5 genotyping ... 33
5.2.2 BRUNOPIC 1st dose ... 34
5.2.3 BRUNOPIC Dose adjustment & Target concentration ... 34
5.2.1 Concomitant therapy ... 35
6 TRIAL VISITS AND PROCEDURES ... 36
6.1 Trial timeline ... 36
6.2 Schedule of assessments ... 37
6.3 Description of procedures ... 38
6.4 Treatment discontinuation, participant withdrawals and losses to follow up ... 39
6.4.1 Withdrawal of consent - participant withdraws from all trial participation. ... 39
6.4.2 Study Closure ... 40
6.4.3 Temporary halt or early termination of a trial ... 40
7 SAFETY MONITORING AND REPORTING ... 40
7.1 Definitions ... 40
7.2 Capturing and eliciting adverse event/reaction information ... 41
7.3 Report of safety events ... 43
8 DATA AND INFORMATION MANAGEMENT ... 43
5.3 8.2 Data management ... 44
8.2 Data confidentiality ... 46
8.3 Quality assurance ... 46
8.1.1 Data sharing ... 46
9 TRIAL OVERSIGHT ... 47
9.1 Governance oversight ... 47
9.1.2 Pharmacogenomic Steering Committee (PSC) ... 47
9.2 Site Monitoring ... 47
9.3 Quality Control and Quality Assurance ... 47
10 STATISTICAL METHODS ... 48
10.1 Sample Size Estimation and statistical analysis plan ... 48
10.2 Handling of missing data ... 49
11 ETHICS AND DISSEMINATION ... 49
11.1 Research Ethics Approval & Local Governance Authorisation ... 49

#### PROTOCOL SYNOPSIS

| | |
|---|---|
| ***STUDY TITLE*** | BRUNO- PIC- Genotype informed Bayesian dosing of tacrolimus in solid organ transplant- Pharmacogenomic Implementation in Children. |
| ***STUDY DESCRIPTION AND BACKGROUND*** | Tacrolimus is a calcineurin inhibitor, a highly effective immunosuppressant used in all solid organ transplantation. Tacrolimus however has a narrow therapeutic index and can vary highly due to individuals and their pharmacokinetic [1,2] leading to incomplete effectiveness, toxicities and suboptimal outcomes [3].<br><br>Tacrolimus is mainly metabolised by cytochrome P450 (CYP) 3A4 and CYP3A5 and studies have found there to be a link between CYP3A5 genotype and tacrolimus dosing [4]. Over 40% of tacrolimus PK variability is predictable, based on CYP3A5 genotype and other measurable covariates [5,6].<br><br>Underexposure in the initial weeks post-transplant is associated with acute rejection [7-9] however genotype-informed dosing algorithms lead to more rapid target concentration attainment overcoming this complication [10-14]. Furthermore, where time outside therapeutic range in the 1st 6-months post-transplant is associated with rejection [15,16], donor-specific antibody formation [17,18], post-transplant malignancy [19] and graft loss [20-22], a genotype-informed maximum *a posteriori* Bayesian dosing [23] can increase time within therapeutic range [24] and potentially overcome these complications.<br><br>In BRUNO-PIC, we will assess for superiority of genotype-informed Bayesian dosing of tacrolimus in attaining and maintaining tacrolimus concentrations within the acceptable range for the first 2 months following kidney, liver and cardiac transplantation in children. Specifically:<br><br>1. First dose will be size scaled by allometry and then adjusted for CYP3A5 and CYP3A4 genotype.<br><br>2. Post-transplant dose adjustment will be guided by NextDose, a Bayesian dosing platform that incorporates genotype, clinical characteristics, measured tacrolimus concentrations and a highly robust population PK model prior to maximally inform dose adjustment recommendations. |

| | |
|---|---|
| | 3. For CYP3A5 expressers, phenotypic rapid metabolisers and any other participants per clinician discretion, dose will be adjusted to drug exposure (tacrolimus Cavgss) rather than the usual trough ($C_0$) target.<br><br>There will be 2 arms to this study, a retrospective cohort compared with the prospective (n=45) cohort.<br>• retrospective SOT recipients<br>• 45 prospective SOT recipients using genotype-informed and Bayesian revised dosing.<br><br>We hypothesize that combined genotype-informed initial dose and Bayesian revised post-transplant dose adjustment dosing will lead to a higher proportion of tacrolimus concentrations within range immediately post-transplant (day 4 of post-transplant dosing), and increased time within therapeutic range in the 2 months post SOT. This will be based on a co-primary outcome of:<br>1. Proportion with acceptable range (80-120% of $C_0$ target) on the 4$^{th}$ day of post-transplant tacrolimus dosing (DD4)<br>2. Proportion in range of over the initial 2 months, based on estimated AUC on DD4, week 3 post-transplant and month 2 post-transplant. |
| ***Objectives*** | To determine if genotype-informed Bayesian dosing of tacrolimus in solid organ transplant recipients leads to better achievement of tacrolimus concentrations compared with a historical control group using standard of care dosing over the first 2 months post-transplant<br>:<br>(i) |

| | |
|---|---|
| ***OUTCOMES AND OUTCOME MEASURES*** | **Co-primary outcome:**<br>1. The proportion of participants within the acceptable range of 80-125% of the treating unit specified Cssavg target on DD4 post-transplant (post-transplant tacrolimus dosing day 4)<br>2. On DD4, Wk 3 & Wk 8 - within 80-125% of Cssavg target (where Cssavg is calculated using Bayesian MAP estimation, from observed concentrations and clinical covariates.<br>**Secondary Outcomes:**<br>1. Time to acceptable tacrolimus concentration (80-125% of Cssavg target, in the immediate post-transplant period<br>2. Time within acceptable range (80-125% of individualised $C_0$ target) over the 2-month period<br>3. Proportion with tacrolimus concentrations within 80-125% of Csstrough on DD4<br>4. Number of dose adjustments of tacrolimus based on TDM and/or Bayesian dosing.<br>5. Safety and feasibility of genotype-informed Bayesian dosing<br>6. Number of negative clinical outcomes: rejection, donor-specific antibody formation and toxicities. |
| ***STUDY SITE AND STUDY POPULATION*** | Renal, Heart and Liver Transplant Units - The Royal Children's Hospital, Melbourne. A combined prospective/retrospective cohort study of 45 prospective and 120 retrospective SOT (3 years recipients recruited from The Royal Children's Hospital Melbourne.<br><br>1. Inclusion criteria<br>• 1- 18yo<br>• Kidney, cardiac or liver transplant recipients (planned or on waiting list) excluding repeat liver transplant.<br>• Amenable to venepuncture and blood draw<br>2. Exclusion criteria<br>• > than 18yo<br>• < than 1yo<br>• Previous liver transplant.<br>• Lung OR Intestinal transplant.<br>• Insufficient time before transplant for pharmacogenomic analysis.<br>• Known hypersensitivity to tacrolimus and/or its formulation.<br>• Slow-release preparation of Tacrolimus (e.g Advagraf XL Brand)<br>• |

| | |
|---|---|
| ***DESCRIPTION OF INTERVENTIONS*** | <u>Tacrolimus</u><br>(Prograf®/ Pharmacor® 0.5mg, 1mg or 5mg IR capsule, Suspension (RCH formula) 1mg/mL<br>• Initial dose based on genotype CYP3A5 as per CPIC guidelines.<br>• Dose(s) as predicted by NextDose Bayesian calculator.<br><u>Genotype testing</u><br>▪ All participants will undergo venepuncture or central line access and a blood draw (3mL for all patients, minimum volume 1mL).<br>• Blood sample sent to VCGS laboratories for targeted CYP3A4/3A5 genotype testing.<br>• A clinical report will be generated within 1 to 2 weeks.<br><u>NextDose</u><br>• A clinical decision support software (CDSS) used to predict Tacrolimus dosage.<br>• Is a web-based Bayesian dose forecasting tool.<br>• Dosing will be led by academic pharmacist and PI. |
| ***TRIAL DURATION*** | Study Duration: 3 years |
| ***PARTICIPANT DURATION*** | Participant Duration: 2 months |
| ***ETHICAL CONSIDERATIONS*** | Research Ethic Approval<br>This study will be reviewed by the Sydney Children's Hospitals Network Human Research Ethics Committee (SCHN HREC) in Sydney, Australia prior to commencing the trial. A letter of authorisation will be obtained from the RGO prior to the commencement of the research at the Royal Children's Hospital.<br>**Informed consent**<br>Patients and parents of patients will be informed of the study with an initial screening during their inpatient stay, or outpatient appointment, and left with a copy of the patient/parent information statement.<br>In accordance with the National Statement, this study will utilise an active consent process. This will involve presenting potential participants or parent/guardian with all the information they need to make an informed decision about participation. When the eligible participant or parent/guardian is invited to participate, they will be provided with detailed information on the purpose of the study, the extent of their |

| | |
|---|---|
| | involvement, the risks associated and the option to refuse participating at all times.<br>Prior to performing any trial-specific procedure (including screening procedures to determine eligibility, signed consent(s) form will be obtained for each participant.<br>There will be 2 cohort of patients; Retrospective and Prospective cohort.<br>For our *retrospective arm* cohort: Waiver of consent.<br>A waiver of the consent to be applied as it impracticable to obtain an individual's explicit consent to the use of their information and the purpose of the research. Tacrolimus doses and concentrations will be assessed in a retrospective manner from patients who have had a solid organ transplant over the last 5 years at the RCH. These results will have no impact on patient. It is purely from a collection data to compare with our prospective data. Data will be collected from the laboratory results available within the electronic medical record (EMR).<br>For our *prospective arm* cohort: An Informed consent will be required. |

| | |
|---|---|
| ***DATA MANAGEMENT*** | BRUNO-PIC results will be kept on the Murdoch Children's Research Institute REDCap database. The database is secure, and password protected. Only researchers involved in the project will have access codes. These access codes will be changed at regular intervals to ensure integrity of the database. All paper files and consents will be kept locked away securely in a filing cabinet within MCRI.<br>Patient parameters along with a unique identifier will be entered into the secure online platform NextDose, along with tacrolimus dosing and concentrations. |
| ***QUALITY AND ASSURANCE*** | The Sponsor-Investigator will have responsibilities in relation to quality management. The Sponsor-Investigator will develop SOPs that identify, evaluate and control risk for all aspects of the trial, (i.e., trial design, source data management, training, eligibility, informed consent and adverse event reporting). Quality control (QC) procedures, which will include the data entry system and data QC checks will also be implemented. Any missing data or data anomalies will be communicated for clarification/resolution. |
| ***STATISTICAL ANALYSIS*** | A sample size was calculated based on comparing proportion of patients within each group with a $C_0$ concentration within therapeutic range on day 4 post transplantation based on prior studies. We assumed 27% of the retrospective control cohort would have a tacrolimus $C_0$ within target range on DD4. 165 participants (120 control and 45 intervention participants) would |

| | |
|---|---|
| | provide us 80% power to detect a risk difference of 24% assuming a two-sided alpha of 0.05. |

GLOSSARY OF ABBREVIATIONS

| ABBREVIATION | TERM |
|---|---|
| AE | Adverse Event |
| ANOVA | Analysis of Variance |
| AR | Adverse Reaction |
| AUC | Area under curve |
| BRF | Biobank Registration Form (MCRI) |
| CDSS | Clinical decision support software |
| CNI | Calcineurin Inhibitors |
| CRF / eCRF | Case Report Form / electronic Case Report Form |
| CPIC | Clinical Pharmacogenomic International Consortium |
| CYP | Cytochrome P450 |
| DMC SMC | Data Monitoring Committee / Safety Monitoring Committee |
| DDI | Drug-Drug Interaction |
| DD0 | Post-transplant tacrolimus Dosing Day 0 (day 0 is the first dosing day) |
| DT0 | Day post transplant 0 (day 0 is the day of transplant) |
| eGFR | Estimated Glomerular Filtration Rate |
| EMR | Electronic Medical Record |
| FDA | Food and Drug Administration |
| GCP | Good Clinical Practice |
| GLP | Good Laboratory Practices |
| GMP | Good Manufacturing Practices |
| HREC | Human Research Ethics Committee |
| IB | Investigator's Brochure |
| ICH | International Conference on Harmonisation |
| IMP | Investigational Medicinal Product |
| ISO | International Organisation for Standardisation |
| ITT | Intention To Treat |
| LFT | Liver Function Test |
| MAPBE | Maximum a posteriori Bayesian estimation |
| MCRI | Murdoch Children's Research Institute |
| MedDRA | Medical Dictionary for Regulatory Activities |
| MSDS | Material Safety Data Sheet |
| NHMRC | National Health and Medical Research Council |
| PK | Pharmacokinetic |
| PI / CPI | Principal Investigator / Coordinating or Chief Principal Investigator |
| PI | Product Information (available for an approved drug or device) |
| QA | Quality Assurance |
| QC | Quality Control |
| RGO | Research Governance Office |
| RCH | Royal Children's Hospital (Melbourne) |
| SAE | Serious Adverse Event |
| SAP | Statistical Analysis Plan |
| SAR | Serious Adverse Reaction |
| SMC | Safety Monitoring Committee |
| SoA | Schedule of Assessments |
| SOP | Standard Operating Procedure |
| SOT | Solid Organ Transplant |

| | |
|---|---|
| SSI | Significant Safety Issue |
| SUSAR | Suspected Unexpected Serious Adverse Reaction |
| TCI | Target Concentration Intervention |
| TDM | Therapeutic Drug Monitoring |
| TGA | Therapeutic Goods Administration |
| UAR | Unexpected Adverse Reaction |
| USM | Urgent Safety Measure |
| VCGS | Victorian Clinical Genetics Services |

**INVESTIGATOR AGREEMENT**

I have read the protocol entitled "***BRUNO-PIC***".

By signing this protocol, I agree to conduct the clinical trial, after approval by a Human Research Ethics Committee or Institutional Review Board (as appropriate), in accordance with the protocol, the principles of the Declaration of Helsinki and the good clinical practice guidelines adopted by the TGA [Integrated Addendum to ICH E6 (R1): Guideline for Good Clinical Practice E6 (R2), dated 9 November 2016 annotated with TGA comments].

Changes to the protocol will only be implemented after written approval is received from the Human Research Ethics Committee or Institutional Review Board (as appropriate), with the exception of medical emergencies.

I will ensure that trial staff fully understand and follow the protocol and evidence of their training is documented on the trial training log.

| **Name** | **Role** | **Signature and date** |
|---|---|---|
| A/Prof Rachel Conyers | Co–PI | |
| Dr David Metz | Co–PI | |
| Ms Dhrita Khatri | Associate Investigator | |
| Ms Tayla Stenta | Associate Investigator | |
| Prof Nick Holford | Associate Investigator | |
| A/Prof Joshua Kausman | Associate Investigator | |
| Dr Jacob Mathew | Associate Investigator | |
| Dr Elizabeth Bannister | Associate Investigator | |
| Julia Netylko | Associate Investigator | |

### 1. ADMINISTRATIVE INFORMATION

- **Trial registration**

*ClinicalTrials.gov* NCT 06529536

- **Sponsor**

| | |
|---|---|
| **Trial Sponsor** | Murdoch Children's Research Institute |
| **Contact name** | Kate Scarff (Melbourne Children's Trials Centre) |
| **Address** | Royal Children's Hospital, 50 Flemington Road, Parkville VIC 3052 |

- **Contributorship**

| **Name** | **Summary of contribution** |
|---|---|
| Dr David Metz | Protocol design, background and rationale, patient and parent information statements, ethics modifications and submissions.<br>Lead for NextDose interpretation. |
| A/Prof Rachel Conyers | Protocol design, background and rationale, patient and parent information statements, patient recruitment, ethics modifications and submissions |
| Ms Dhrita Khatri | Protocol design, background and rationale, patient and parent information statements, patient recruitment, ethics modifications and submissions |
| Prof Nick Holford | Protocol design, background and rationale, NextDose development and oversight. |
| Ms Tayla Stenta | REDCap build, ethics modifications and submission |
| A/Prof Joshua Kausman | Patient identification and recruitment, clinical input. |
| Dr Jacob Mathew | Patient identification and recruitment, clinical input. |
| Dr Elizabeth Bannister | Patient identification and recruitment, clinical input. |
| Julia Netylko | Patient recruitment, ethics modifications and submissions. |

- **Expected duration of study**

3 years from start of recruitment

Participants will be involved from pre-transplant to 8-weeks post SOT.

### 2. BACKGROUND & RATIONALE

#### 2.1 Overview

The aim of BRUNO-PIC is to determine if genotype-informed Bayesian dosing of tacrolimus in solid organ transplant recipients increases therapeutic (safe and effective) drug exposure and ameliorates transplant complications related to systemic under- and overexposure to tacrolimus.

#### 2.2 Background

Patient outcomes from transplantation underwent major advances in the last 2 decades of the 20th century, with introduction of highly effective immunosuppressant drugs like tacrolimus, mycophenolate, and anti-lymphocyte antibody induction [26]. Heavy immunosuppression in the initial 6 months post-transplant has dramatically reduced early rejection and graft loss [26], with some of the toxicity burden offset by prophylactic agents against severe and opportunistic infection. Nevertheless, despite dramatic improvement in short term outcomes by the turn of the century, the burden of incomplete effectiveness (rejection) and substantial toxicities remain [27]. In addition, beyond the first 12 months post-transplant improvement in outcomes have been more modest, with limited improvement in immune-mediated graft attrition (in part set up by early alloimmune activity) and problematic cumulative morbidity from immune suppressing drugs.

Successful transplantation requires a balance between providing sufficient pharmacological immunosuppression to prevent organ rejection – continued for the life of the allograft – whilst simultaneously minimising dose-dependent acute and cumulative toxicities [27,28]. These serious toxicities include severe and opportunistic infection [29]; cardiovascular morbidity [30-32]; kidney scarring; and development of cancer [33,34], quantitatively linked with dose, duration and exposure [19,33,35]. Achieving the optimal balance of safety and effectiveness of immunosuppression is substantially hampered by interindividual dose-response variability [27]. For narrow therapeutic index drugs like tacrolimus, used in almost all solid organ transplant recipients, wide between-subject variability in drug exposure at a given dose translate to treatment failure in some individuals, unacceptable toxicities in others, underlying the need to individualise dosing [36].

This dose-response variability is certainly partially ameliorated by the longstanding practice of therapeutic drug monitoring (TDM), with empiric dose adjustment to attain trough (pre-dose) tacrolimus concentrations within a therapeutic range. Nevertheless, the consequences of both underexposure and treatment failure in some [15,18,37-40], unacceptable toxicities in others, remain substantial with the current TDM approach [27,28,36].

##### 2.2.1 Calcineurin inhibitors in SOT – history & current practice

The calcineurin inhibitors (CNI), first cyclosporine (CsA), now superseded by tacrolimus (Tac), are a core component of almost all solid-organ transplant regimens, alongside an anti-inflammatory steroid (e.g., prednisolone/ prednisone), an anti-metabolite (e.g., mycophenolic acid) and in many instances anti-lymphocyte antibody induction. Together, these make up the contemporary "quadruple therapy" drug regimen.

The introduction of cyclosporine in 1978 [41] was transformative for graft and patient outcomes, taking kidney transplantation from experimental to standard of care [42].The superiority of tacrolimus over cyclosporine was most definitively demonstrated in the Elite-Symphony trial in kidney transplant recipients [43].

However, dose-response variability with CNIs remains a major challenge. At the turn of the century the degree to which CNIs were themselves causing kidney scarring, leading to transplant allograft failure, was identified. This led to calls their replacement e.g "cyclosporine is unsuitable as a universal, long-term immunosuppressive agent for kidney transplantation" [44]. The subsequent decade saw attempts to minimise or withdraw CNIs completely from transplant regimen, leading to a major increase in antibody mediated rejection and graft loss.

CNIs are now accepted as a necessary part of transplantation despite this toxicity [45]. However, this further underlines the importance in optimal precision in use of these agents.

- **Tacrolimus in SOT: current dosing practice and clinical need**

Tacrolimus is typically administered orally twice daily, with a starting dose scaled linearly to body weight (mg/kg). Dose is then adjusted empirically in the individual based on measured steady-state trough (pre-dose) whole blood tacrolimus concentrations, to bring to within a desired "therapeutic range". However, this dosing strategy remains associated with incomplete effectiveness and toxicities in a substantial proportion of recipients (3), related to under- or over-exposure respectively.

There are three situations where effectiveness of dosing strategy remains suboptimal. First, in choice of initial dose, noting that some recipients take 2 weeks or more to attain optimal exposure, with underexposure associated with acute rejection. Second, in the initial post-transplant months, where the combination of substantial between-occasion variability in Tac PK and the "trial and error" approach of TDM (as distinct from more targeted approaches) lead to significant time outside therapeutic range, associated with rejection and graft loss as well as toxicities. Third, trough-based dosing in individuals who are pharmacogenetic "rapid metabolisers" leads to excessive toxicities, related to either excessive peak or AUC to achieve target trough concentration.

These three moments of suboptimal precision could be overcome with a target concentration intervention (TCI) strategy, using genotype-informed Bayesian dosing. Indeed, Bayesian dosing has already been proven superior to clinician based TDM by randomised controlled trial in 2 adult kidney transplant cohorts [24,46] though this has not been shown in the paediatric population. After reviewing the pharmacology and pharmacogenetics of tacrolimus, we describe in detail the potential role of genotype-informed Bayesian dosing in improving patient outcomes.

- **Pharmacology and ontogeny of relevance to tacrolimus**

Tacrolimus, a calcineurin inhibitor, diffuses into T-cells where through inhibition of calcineurin activation it prevents NFAT dephosphorylation, required for NFA translocation into the cell nucleus where they promote IL-2 transcription. It thus inhibits a key step in T-cell activation and proliferation following engagement of antigen with the T-cell receptor.

Tacrolimus itself displays a wide between-subject variability in bioavailability and metabolism. An oral dose is typically rapidly absorbed, with peak concentration 30-60 minutes after dose, though efflux transporters [47] and 1st pass metabolism lead to a low and highly variable bioavailability, mean of 25% with range 4 to 89% [48]. In blood, tacrolimus is extensively bound to erythrocytes, as well as plasma protein binding primarily to alpha-1 acid glycoprotein. These impact interpretation of whole blood tacrolimus concentrations, which co-vary with patient haematocrit [49].

Tacrolimus undergoes metabolism by CYP3A4 and, in expressers, CYP3A5 [50,51]. These drug metabolising enzymes reside predominantly in the liver, but also in enterocytes, contributing to 1st pass metabolism as well as drug clearance. All individuals express CYP3A4, with expression increasing from minimal levels at birth, increasing over infancy with "full adult values reached by 1 year of age" [52].

On the other hand, CYP3A5 is only expressed in a proportion of the population, however for those who do it is associated with increased metabolism of tacrolimus, requiring on average 1.5 to 2.5 times the typical dose for equivalent exposure [51]. Expression of CYP3A5 is 10-15% in Caucasian populations, though much higher in African (40-50%) and Asian (50-70%) populations [53]. There is consensus on need for higher initial dosing of tacrolimus for CYP3A5 expressers when genotype is known [51], though quantitative differences. Specifically, the Clinical Pharmacogenetics Implementation Consortium (CPIC) and the French National Network of Pharmacogenetics (RNPGx) recommend 1.5-2 times standard dosing for both heterozygous and homozygous expressors, whereas the Dutch Pharmacogenetics Working Group (DPWG) recommends 1.5 times normal dose for heterozygous expressors, and 2.5 times normal dose for homozygous expressors [54].

Pharmacogenetic variants in CYP3A4 have been variably reported as impacting tacrolimus disposition and clinical utility remains debated. However, this may in part relate to the prevalence of CYP3A4 variants in the populations studied, with the variant CYP3A4*22 having greater prevalence in European cohorts and clinical data supporting impact on dose requirement from this region [55]. The French National Network of Pharmacogenetics recommends testing for this pharmacogenetic variant, though accepting that the evidence is less robust [54].

In liver transplantation it would be expected that the donor not recipient genotype would be relevant for tacrolimus disposition. Importantly, however, it has been shown that it is the recipient genotype that is relevant in the initial weeks post liver transplant, hence useful for initial dosing recommendation [56].

Although studies have found that whole-blood tacrolimus and tacrolimus doses correlates with CYP3A5 polymorphism, patients listed for a transplant at RCH do not routinely undergo genotype testing. The tacrolimus dose is based on a linearly scaled body weight (mg/kg) and target concentrations are reached based on a “trial and error” approach as opposed to a targeted approach.

Other important covariates that have been shown to contribute to between-subject and between-occasion variability in tacrolimus disposition are size (allometric scaling of free fat mass), steroid dose, and haematocrit [57], though the importance of the latter is in interpretation of whole blood concentrations rather than impacting intrinsic hepatic clearance of tacrolimus [49].

##### 2.2.2 Limitations of current dosing and proposed intervention

- **Initial dose**

Despite initial "quadruple therapy" induction, tacrolimus underexposure in the initial weeks post-transplant remains linked with acute rejection [7-9]. This reflects either insufficient initial dosing, e.g., in "rapid metabolisers" who require on average a 1.5 to 2-fold higher dose than the typical individual [58], or failure to rapidly adjust to achieve target exposure, some studies reporting up to 3 weeks before reaching target using standard dosing and TDM practice [25].

Genetic "rapid metabolisers" take longer to reach target concentration [4] unless status is known a priori [10], and CYP3A5 expression has been linked with negative outcomes including higher rates of donor specific antibody formation [59], rejection, interior graft survival and inferior patient survival [22]. Importantly, genotype-informed initial dosing leads to more rapid target concentration attainment, based on meta-analysis of RCTs [10-14]. However, although genotype contributes substantially to dose requirement in expressers, it is not the full picture, with the addition of patient covariates increasing the predictable component of tacrolimus PK from 16.3 % to 43.4 % [5]. This may explain why the recent meta-analysis did not see improvement in clinical outcomes despite faster time to target concentration attainment [13]. The authors thus concluded that future trials should look at "dosing algorithm that include demographic and clinical factors plus multiple genetic variants" [13], a key aspect to the current trial.

- **Maximizing time within therapeutic range in the early months**

The second shortcoming in current tacrolimus dosing strategy is the practical difficulty, in clinical practice, to consistently maintain drug concentrations within the therapeutic range over the initial post-transplant months. This has been demonstrated both under controlled trial conditions [60] and in clinical practice [61,62], with reports of in some instances *less than 60% of tacrolimus concentrations within therapeutic range* over the initial months [62]. Crucially, time within therapeutic range has been clearly linked with outcomes, including acute rejection [15,16], donor-specific antibody formation [17,18], post-transplant malignancy [19] and graft loss [20-22].

There are 2 aspects to difficulty in maintaining safe and effective exposure in the initial post-transplant months. First, the empiric titration of therapeutic drug monitoring is both highly user-dependant and less accurate than more modern techniques [63]. Standard implementation of concentration-controlled dosing is to measure tacrolimus drug concentration in whole blood immediately prior to the morning dose (trough concentration) at steady state. If the concentration is outside of the "therapeutic range", the clinician makes an empiric dose adjustment to push concentration into range. We have referred to this practice as "therapeutic drug monitoring" (TDM), to differentiate from the more precise TCI [63]. The latter involves a concentration target (rather than therapeutic range), and a specific PK-guided intervention to maximise likelihood of attaining target drug exposure. Its utility across a range of drugs, both in trial conditions and clinical practice, is well established [63,64], including for tacrolimus in randomised controlled trial [24,46].

Second, there is considerable between-occasion variability in PK over this time period, with changing tacrolimus disposition related to haematocrit, concurrent prednisolone dose, time-post transplant, and other unpredictable variability [49,57]. Thus, relationship between dose and concentration at a given time post-transplant will systematically change with time post-transplant. Crucially, predictable components of this between-subject variability can be incorporated into population PK models used for Bayesian dose adjustment (see below), leading to improved target concentration attainment as demonstrated both in virtual external validation datasets [57] as well as randomised controlled interventional trial [24,46].

- **Avoiding unnecessary overexposure in rapid metabolisers**

Using trough concentration to assess drug exposure is a common strategy for most Narrow therapeutic index drugs. Importantly, however, for most drugs - including CNIs [7,65] - trough concentration is a surrogate for systemic exposure (area under the concentration-time curve, AUC), rather than being directly linked with drug effect.  Nevertheless, the use of trough concentration in this setting is pragmatic and practical in clinical care, compared to multiple samples to estimate AUC. The trough (pre-dose) concentration also has the advantage that it is relatively insensitive to timing, given flattest segment of concentration-time curve between doses [66].

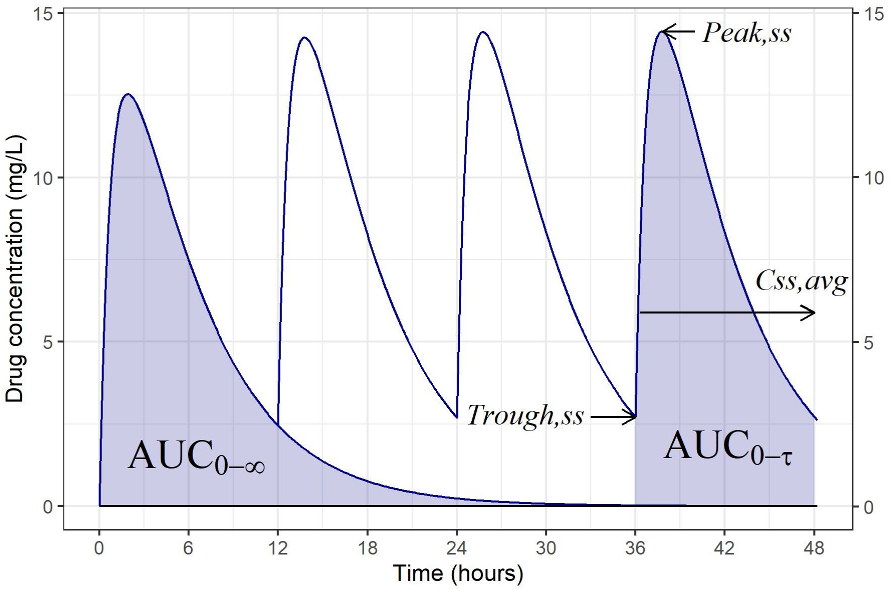

**Figure 1: The time course of concentrations in blood following multiple oral drug doses.** AUC = area under the concentration-time curve, Cav,ss = average concentration at steady state, Peak,ss = maximum (peak) concentration at steady state, Trough,ss = minimum concentration at steady state, ∞ = infinity, t = tau, dosing interval**.**

Whilst trough concentration may be an “adequate” surrogate for total drug exposure for many recipients (correlation coefficient up to 0.91) [66], variability remains, as shown in kidney transplant recipients where despite trough in the “therapeutic range” of 5-10 mcg/L, AUC can range 3-fold (from 75–225 mcg/L.h) [67]. Thus, some expert groups have thus called for AUC monitoring over use of trough concentration alone [36,67].

More importantly, the greatest potential benefit would be expected in those in whom tacrolimus PK

is substantially different to the typical population, e.g., so-called “rapid metabolisers”. These individuals with substantially higher tacrolimus metabolism, due either to CYP3A5 expression [68], or other as yet unidentified source of variability [69], require substantially higher doses to achieve the same trough concentrations as individuals with more typical PK [22]. Yet this in turn often translates to higher peak concentrations and higher overall tacrolimus systemic exposure (AUC) [53,70], predictable from PK principle and shown in clinical PK studies [70]. This higher tacrolimus exposure, whilst achieving the same trough concentration, explains the higher nephrotoxicity [68,71-73] and other toxicities e.g. BK virus nephropathy [74] in tacrolimus rapid metabolisers. Proof of principle for higher peak concentrations contributing to tacrolimus nephrotoxicity has been observed in a trial of early conversion of rapid metabolisers to LCP-Tac, a slow-release product with flattened peak concentration [75], as well as improvement in tremor [76]. This adds to the observational evidence of toxicities in rapid metabolisers dosed to the typical trough range [22,53,77].
Thus, “rapid metabolisers” are at risk of rejection if fail to achieve therapeutic concentrations (see initial dosing, above), and nephrotoxicity (and other toxicities) if they do consistently achieve them [77].

Given the consistent supportive evidence from pharmacokinetic-pharmacodynamic and clinical data [22,53], it has been proposed that rapid metabolisers would be better served by dose adjustment to an AUC target (with corresponding trough therapeutic range) [78]. Another group has shown durability of the relationship between trough and AUC over time for an individual [21], supporting an approach of AUC target (and corresponding lower trough target) for such recipients. Finally, there is published precedence for tacrolimus dosing to target AUC as far back as 2005, using Bayesian estimation [79], and basis for AUC targets derived from very large cohort studies linking trough concentration ranges with corresponding AUC ranges in typical populations. Identification of rapid metabolisers (through genotype or measured concentrations) and targeting AUC rather than trough concentration in these individuals through MAPBE can reduce systemic overexposure [79] and thereby should reduce toxicities like BK virus nephropathy and CNI nephrotoxicity.

Given the above, and facilitated by use of Bayesian estimation software, on identification of rapid metaboliser participants we will in this subgroup use an AUC target rather than trough, to avoid unnecessary overexposure and excess toxicities [53].

- **Genotype-informed Bayesian estimation to optimize initial & ongoing dosing.**

Genotype-informed Bayesian estimation, along with detailed understanding of tacrolimus PKPD, provides an opportunity to address all 3 of the above shortcomings in current tacrolimus dosing practice. This is through:

- Optimising initial tacrolimus dose to the individual based on pharmacogenotype, accurate size-based dosing using allometric scaling to free-fat mass, and other identified covariates impacting tacrolimus PK, computed through a Bayesian dosing platform.
- Increasing time within therapeutic range over the first 12 weeks post-transplantation by Bayesian adaptive dosing to a target concentration, accounting for predictable between-occasion variability over the initial months.
- Avoiding unnecessary systemic overexposure to tacrolimus, and consequent toxicities, in

individuals who rapidly metabolise tacrolimus due to pharmacogenetic differences, by identifying these individuals and changing the target to estimated $C_{av,ss}$ rather than trough concentration, at a $C_{av,ss}$ equivalent to that targeted in individuals with typical PK.

**Maximum *a posteriori* Bayesian estimation (MAPBE)**, also referred to in the literature as "model-informed precision dosing"[80] and "target concentration intervention" [63], is an established pharmaco-statistical technique that leverages quantitative information of drug pharmacokinetic behaviour and variability (the Bayesian prior) with an individual's characteristics and measured drug concentrations, to accurately predict dose required to attain target concentration [81].

It is used in clinical care, though uptake varies globally [80,81]. Key barriers to implementation including unclear regulatory framework (though guidance now available through TGA since 2021); the choice of platforms and model validation; clinician understanding and acceptance of the complex technique; and comparative clinical trial evidence [80].

Bayesian dosing software estimates an individual's drug pharmacokinetic (PK) parameters by combining knowledge about a drugs PK behavior with information from the individual: clinical characteristics (e.g. weight, age, renal function, genotype) and measured drug concentrations.

- A population PK model is a mathematical-statistical model developed in a real patient population which describes the drug's typical pharmacokinetics, how this varies between individuals and over time, and factors that influence this variability.
- By leveraging prior knowledge about a drug's pharmacokinetics with individual patient characteristics and drug concentrations, an accurate estimate of an individual's PK characteristics can be determined. Furthermore, additional drug concentrations over time (subsequent days) iteratively improve accuracy of individual PK estimates.

The Bayesian prior, a ***population pharmacokinetic (PPK) model,*** is a mathematical model developed using dose, drug concentration and covariate data from a population receiving the relevant drug, and contains quantitative information on "how the drug behaves": its basic compartmental characteristics and typical PK parameters; how these PK parameters vary within a population; and the covariate or patient characteristics that influence the drugs PK. PK modelling has an accepted role and is used extensively in drug development to describe a drugs PK, simulate, and predict its behaviour in special populations, and thereby develop safe and effective dosing recommendations.

NextDose is a clinical decision support software. The NextDose output gives a dose recommendation, as well as providing a visual output of the predicted concentration-time curve and how this relates to the observed concentrations. This allows clinical review of recommendation and decision on whether to implement.

Tacrolimus Bayesian dose adjustment has, in some regions of the world, longstanding precedence for use [82-84]. The underlying pharmacometric technique is well validated, and MAPBE allows incorporation of the range of covariates that influence its PK to better predict initial dose and allow accurate Bayesian dose adaption including through the early weeks and months of higher between-occasion variability. This has been shown in real world data, and crucially now in ***2 randomised controlled trials in adult kidney transplant recipients***, with superior target concentration attainment than current clinical practice [24,46]. This has not been shown in children to date.

##### 2.2.3 NextDose, and applicability to paediatric SOT cohort

- **NextDose for Tacrolimus MAPBE**

There are a range of platforms for clinical implementation of MAPBE [62,80], some commercial at cost, some academically driven and free to use, NextDose in the latter category.

Developed and first introduced into clinical care in New Zealand in 2012 (for Busulfan), it has expanded over the past decade to now include Bayesian estimation for 15 different medicines, with the population PK model behind tacrolimus developed and externally validated in 2014 (62). This model is currently in use at multiple centres in New Zealand. It has also been used by PI Metz on occasion over the past 5 years for kidney transplant recipients displaying variable pharmacokinetics and has been found to be consistently accurate in target concentration attainment [57].

Crucially, there is published evidence and independent external validation supporting superiority of the tacrolimus model to others currently available.

- First, the model used by NextDose is the largest model of its kind (excluding those that only contain trough concentrations, a major limitation), developed from a population of 242 kidney transplant recipients and 3100 tacrolimus whole blood concentrations, with 64 full PK profiles (> 8 concentrations per occasion), over 153 limited sampling profiles and 1546 trough concentrations [57].
- Second, the model was unique in using principles of mechanistic modelling for both structure and covariate introduction [57,85], a principle not applied consistently applied in population PK modelling however with clear basis in pharmacometrics (and statistical modelling more generally) to reduce bias and enhance external validity [86].
- Third, it was developed by combining two separate populations, and subsequently outperforming these models in external validation [57]. Notably, one of the populations used to develop the model was that subsequently used in the one of the successful RCTs of tacrolimus Bayesian dosing [24].

  Finally, its superiority has been reported in published systematic review and external validation, e.g. *"the model by Størset et al, which was comparatively the best of all 16 published models"* [85].

- **Use of NextDose in children**

The NextDose tacrolimus model was developed in an adult kidney transplant population. However, though it is true that "children are not small adults" (85), it is important to note that there are many known similarities and predictable differences that this historical idiom obscures. This prompted the call by Gillis J et al in 2007 (86):

> *"it is time to go beyond the 'not just small adults' conceit. There is no doubt that the phrase has served paediatrics well in helping argue for the development of specialist services and, in the broader context, for the protection of children from exploitation. But when trying to convey to fellow health professionals and students the important differences in paediatric*

*practice, this conceit has the potential of becoming vacuous and precious; and possibly deprives child medicine of significant developments."*

Most importantly for the current trial, which involves dose prediction from population PK, it has been well demonstrated that children *are* small adults, infants and toddlers immature children, in relation to pharmacokinetics from infancy to adulthood (87). Specifically, once the mechanisms of drug clearance have matured – for tacrolimus, by 1-2 years of age (89) (90) - drug clearance increases predictably with increasing size from childhood to adulthood (87, 91). This underlies the application allometric size scaling to predict dose requirement in children using adult population PK data (87).

In addition to this well-established PK principle, there is confirmatory clinical data for tacrolimus. Indeed, whilst published tacrolimus PK models have been either in children or adults, it is notable that paediatric models range from early childhood to late adolescence, despite the inarguable reality that PK in adolescence and adulthood is the same. Secondly, where the same research groups have developed models both in children and adults separately, the PK parameters and concentration-time course are equivalent with scaling for size. By way of example, the Limoges group tabulated adult and paediatric exposure measures relating trough and AUC (Table 1), near identical between adults and children [82].

**Table 1: Relationship between tacrolimus trough concentration and AUC, adults and children.**
Adapted from Marquet et al 2021(82)

| | *0-3 months Post-transplant* | *3–12 months Post-transplant* | *Beyond 12 months* |
|---|---|---|---|
| *$C_0$ range: 3–7 ng/ml* | | | |
| *Proposed paediatric $AUC_{0-12h}$ range* | 85–155 | 80–140 | 70–130 |
| *Proposed adult $AUC_{0-12h}$ range* | 75–140 | 80–140 | 75–130 |
| *$C_0$ range: 5–10 ng/ml* | | | |
| *Proposed paediatric $AUC_{0-12h}$ range* | 120–200 | 110–190 | 100–170 |
| *Proposed adult $AUC_{0-12h}$ range* | 110–190 | 110–180 | 100–170 |
| *$C_0$ range: 8–12 ng/ml* | | | |
| *Proposed paediatric $AUC_{0-12h}$ range* | 170–240 | 155–220 | 140–200 |
| *Proposed adult $AUC_{0-12h}$ range* | Na | 150–210 | 140–200 |

- **NextDose use in nonrenal transplant recipients**

The PPK model for NextDose was developed in a large, adult kidney transplant population. However, for use in other organ transplant groups, pooled models from renal and nonrenal transplant recipients have been published [84,87,88], showing relatively consistent PK *apart from liver transplant recipients*, where tacrolimus clearance is substantially lower in the initial weeks [87].

This is suggested to be due to reduced metabolism in the recovering liver allograft. However, it has been shown that, by adding a parameter to reduce clearance in liver transplant recipients – or in the case of physiology-based PK models by inputting mildly impaired hepatic function – models initially developed in kidney transplant recipients can accurately predict tacrolimus concentrations in liver transplant recipients [88]. This data will be used to add an additional parameter for reduced metabolic clearance in NextDose, to allow for first dose prediction in liver transplant recipients.

We will aim to complete this study in two phases:

**Phase 1**: Enrolling renal transplant recipients.

Concurrently, data from published popPK models in nonrenal organ transplant recipients will be used to add a covariate factor for organ type to the NextDose model, followed by external validation using retrospective data from the prior 5 years at RCH (see below).

**Phase 2:** Enrolling other SOT recipients (heart and liver).

The second phase will commence following validation of the adjusted NextDose model for nonrenal organ transplant recipients – see section 5.1.4

#### 2.3 Summary

The BRUNO-PIC study will determine whether Bayesian forecasting performs well when applied to make subsequent dose predication across all SOT. Bayesian forecasting may lead to improved patient outcomes by more rapid target concentration attainment immediately post-transplant; maximising time within therapeutic range in the initial 8 weeks post-transplant; and targeting an estimate of AUC (rather than trough concentration) in individuals with rapid tacrolimus metabolism, identified by genotype or Dose/Concentration ratio.

#### 2.4 Risk/Benefit assessment

##### 2.4.1 Potential Benefits.

Participating in this study will have several potential benefits.

- Potential direct benefits from intervention:
  1. Receipt of a pharmacogenetic test for CYP3A5 & CYP3A4 prior to transplantation, which *has been shown to improve time to therapeutic exposure* in both adult and paediatric solid organ transplant recipients. This could reduce risk of acute rejection in pharmacokinetic outliers without increasing toxicities.
  2. Use of genotype-informed Bayesian dosing, which has been shown by RCT in adult kidney transplant recipients to improve time within therapeutic range in the early post-transplant period (and would be expected to extrapolate to the paediatric population down to 1-2 years of age). This has potential to reduce risk of acute and chronic rejection, tacrolimus toxicities, and graft failure.
  3. Adjusting dose to estimated $C_{av,ss}$ rather than trough in "rapid metaboliser" participants, as supported by mechanistic and observational evidence with precedence in adult transplant recipients, could decrease kidney scarring and toxicities associated with overexposure in this population.
- Potential reduced costs to the healthcare system. Bayesian dosing can help to reduce the cost of tacrolimus therapy by reducing the need for dose adjustments.
- Facilitate the expansion of evidence-based pharmacogenomic therapy in paediatric populations, especially in children receiving a solid organ transplant.

- Assistance in future drug development, improvement, and safety.
- Potential contributions to workforce training and development.
- Possible improvement, interoperability, and integration of health care record systems.

###### 2.4.2 Potential risks.

Participants in BRUNO-PIC will receive the same immunosuppressant regimen as standard care, including tacrolimus. Tacrolimus concentration target will be consistent with unit practice, or equivalent Cavgss (AUC/tau) based on published population data.

Participants will have tacrolimus concentrations taken consistent with standard practice. In addition, a limited sampling profile will be taken on day 4 of tacrolimus dosing (DD4), and on weeks 3 and 8 post-transplant. For kidney transplant recipients these are consistent with current practice (at time of weeks 3 and 8 mycophenolic acid sampling profiles), with the addition of day 4 profile though noting this will be from central line thus no additional venipuncture required.

Bayesian dosing in kidney transplantation has proven benefit in adults (superior exposure control) based on RCT data, and NextDose has long use precedence. Further details on NextDose, the tacrolimus PPK model used in the Bayesian estimation step, and applicability to paediatric kidney transplant recipients is outlined in section 2.2.6.

Finally, the NextDose output will be overseen by  academic pharmacist and/or CRA and Dr David Metz (paediatric nephrologist and clinical pharmacologist experienced with use of Bayesian dosing software and PPK modeling).

***See Appendix 1 for additional potential risks and mitigations.***

#### 3. TRIAL OBJECTIVES AND ENDPOINTS

##### 3.1 Objectives

###### 3.1.1 Primary objective

In paediatric SOT recipients, to determine whether genotype-informed Bayesian dosing of tacrolimus leads to better achievement of tacrolimus concentrations compared with a historical control group using standard of care dosing over the first 2 months post-transplant.

###### 3.1.2 Secondary objectives

1. To determine the safety and efficacy of using a genotype-informed Bayesian dosing of tacrolimus in SOT within the initial 8-weeks post-transplant. (prospective arm only, descriptive)
2. To determine the impact of using a genotype-informed dosing of tacrolimus compared to a historical control group using standard of care dosing in our paediatric SOT cohort in reaching and maintaining therapeutic concentrations.
3. To evaluate the feasibility of using NextDose in the context of paediatric SOT.

##### 3.2 Endpoints

##### 3.2.1 Primary endpoint:

Proportion of cohort with tacrolimus concentration within 80-125% of concentration target:

a) On post-transplant dosing day 4 (DD4) - within 80-125% of  Cssavg target

b) On DD4, Wk 3 & Wk 8 - within 80-125% of Cavgss target (where Cssavg is calculated using Bayesian MAP estimation, from observed concentrations and clinical covariates )

##### 3.2.1 Secondary endpoints

a) Time to acceptable range (80-125% of Cssavg) in the immediate post-transplant period.

b) Time within acceptable range (80-125% of individualised $C_0$ target) in the first 8-weeks post-transplant.

c) Proportion  with tacrolimus concentrations within 80-125% of Csstrough on DD4

d) Number of dose adjustments of tacrolimus based on TDM and/or Bayesian.

e) Safety and feasibility of genotype-informed Bayesian dosing

f) Number of clinical outcomes: rejection, donor-specific antibody formation and toxicities.

##### 3.3 Justification for endpoints

Highly effective immunosuppression in the early post-transplant period has led to low rates of hard outcomes early post-transplant (acute rejection, graft loss, death), albeit with suboptimal long-term outcomes. In addition, despite excellent short-term gains, the low prevalence of hard end-points has made testing of new interventions very difficult without trials with huge numbers of participants, to give adequate power to detect superiority.[27,28] This has led to academic and now regulatory calls for innovative approaches including surrogate biomarkers of long-term outcome.

There exists robust data linking tacrolimus underexposure and acute rejection, as well as time outside of acceptable range in the first 3 and 6-months post-transplant and negative short and long-term outcomes (rejection, de novo DSA formation, graft loss), with clear and causal mechanistic explanation. Thus, evidence for superior tacrolimus concentration control, with increased time within therapeutic range, can reasonably be extrapolated to better long-term outcomes.

#### 4 TRIAL DESIGN

##### 4.1 Overall design

Prospective, open-label single arm intervention trial with retrospective standard care comparator.

Forty-five prospective paediatric recipients undergoing kidney, liver or cardiac transplantation will be recruited to BRUNO-PIC and compared to  120 retrospective recipients (taken from the immediate prior 5 years).

For all eligible SOT patients, the initial tacrolimus dose will be determined by allometric size scaling from adult dose, with adjustment based on genotype (CYP3A4 & CYP3A5). Subsequently, dose adjustment to measured tacrolimus concentrations in the individual will be adjusted using Bayesian dosing to a target concentration. This will be to a metric of overall exposure (Cavgss, equivalent to

AUC/tau) in all kidney transplant recipients (for which good evidence for exposure target exists) and, for other transplant recipients, in rapid metabolisers (phenotypic or CYP3A5 expressers).

Comparison between prospective and retrospective arm (using standard mg/kg dosing plus therapeutic drug monitoring) will be based on proportion in therapeutic range immediately post-transplant (DD4) and over the initial 8 weeks (assessed at D4, W3 and W8).

#### 4.2 Justification for dose and target concentration

The initial dose of tacrolimus will be determined using CYP3A5 & CYP3A4 genotype and allometric scaling for size. Subsequent dose adjustments (beyond DD4) will be guided by NextDose, which uses a tacrolimus population PK *prior*, the participants clinical characteristics (see below) and dose and concentration data to estimate an individual's pharmacokinetic parameters and hence optimal dose.

##### 3.2.2 Initial dosing (see APPENDIX 2 for details)

***Initial dose*** is determined by mechanism-based size scaling using free fat mass and allometry, as opposed to linear scaling of weight (mg/kg), combined with genotype. For detailed justification see APPENDIX 2: Rationale for initial dose determination.

Allometric scaling more accurately describes the relationship between size and drug clearance compared to e.g. linear functions (mg/kg), as well as having robust theoretic underpinnings.[96,97] For tacrolimus, fat free mass is the best size descriptor used with allometric scaling.[57]

In Australian and New Zealand, dosing data from all kidney transplant episodes is collected within a registry, ANZDATA. Analysis of this data shows that the median initial dose of tacrolimus is 5 mg/70 kg/dose twice daily (unpublished from ANZDATA, Metz PhD thesis)[93] Dose size in BRUNO-PIC will be scaled by allometry from a dose of 5 mg twice daily in a 70 kg adult.

Next, the quantitative impact of genotype (CYP3A4 and CYP3A5) will be added to allometric scaled dose. The effect of genotype was derived from CPIC and from a review of popPK literature (see APPENDIX 2). Proportional adjustment based on genotype is:

- Non-expresser: No change
- CYP3A4*22 expresser: Dose x 0.74
- CYP3A5*1 expresser (hetero- or homozygous): Dose x 1.59
- Both CYP3A5*1 & CYP3A4*22: Dose x 1.18

There are several notable recent publications with genotype informed 1$^{st}$ dose. Francke et al 2021[12], in a single arm prospective trial in adult kidney transplant recipients, achieved 58% of recipients within therapeutic range on day 3 post kidney transplantation, using an algorithm containing both CYP3A5 and CYP3A4 status (expresser status leading to x1.62 of dose and 0.814 of dose respectively). Their algorithm was from a popPK model published in 2018 by Andrews et al.[101]

Lloberas et al 2024[46] also improved early exposure using CYP3A5 and CYP3A4 genotype, though those expressing CYP3A5*1/*1 and poor metaboliser CYP3A4*22 led to similar dose as SOC in effect cancelling each other out.

##### 3.2.3 Subsequent dosing (See APPENDIX 3 for details)

***Subsequent dosing*** will be by Bayesian estimation using NextDose. Bayesian dosing aims to increase safety & effective of tacrolimus by more accurate individualised dosing.[63] Bayesian forecasting of individual pharmacokinetics is a statistical process that leverages an individual's characteristics, measured concentrations and dosing schedule with a previously developed population PK model that describes the drugs compartmental kinetics and quantifies PK parameter variability and the covariates that influence them. Maximum likelihood estimation is used to determine the "best fit" concentration time course to observed concentrations and thus predict the individuals drug clearance. This allows determination of the dose required to achieve target concentration (92) (93) (94).

The NextDose target concentration will be either an average **steady state concentration (Cssavg)** or a **Csstrough** (the more standard of care practice). **Cssavg** will be used in all CYP3A5 expressers, as well as other "phenotypic" rapid metabolisers defined as $C_0$/Dose < 1.05.[53] For other participants, either **Cavgss** or **Csstrough** can be used, determined in advance based on unit agreement.

The Csstrough target will be the mid-point of the unit-specified therapeutic window. The acceptable range will be the same as the therapeutic windows.

**Cavgss** is equal to the $AUC_{0\text{-}DI}$ divided by the dosing interval (DI). It is the pharmacological metric better linked with pharmacodynamic effect, and for rapid metabolisers in particular avoids unnecessary overexposure (excessive $AUC_{0\text{-}12h}$ and Cmax to attain desired $C_0$). The specific **Cavgss** target is obtained from steady state AUCs over a dosing interval (AUCssDI). The C0 associated AUCssDI provides a guide to show that on average the **Cssavg** corresponds to the widely using C0 targets (see **Determining Cavgss target**).

The target concentrations are standardized to a haematocrit of 45% (HCT45, table below). See APPENDIX 3 for further explanation.

From this, target concentrations for the stage 1 (kidney transplant) cohorts are shown below, including calculations (see Table 1). The values used for calculation have been shaded grey, with the final targets in the final 2 columns.

***Table 1: Conversion from protocol C0 target to Cavgss.***

| Kidney Tx | HCT non-standardized | | | | | HCT45 | | |
|---|---|---|---|---|---|---|---|---|
| Post KTx month | C0 low | C0 target | C0 high | Equiv. $AUC_{0\text{-}12h}$[a] | Equiv. Cavgss[b] | Trough target[c] | $AUC_{0\text{-}12h}$ target | Cavgss target[c] |
| M1 | 8 | 10 | 12 | 175 | 14.6 | 13.6 | 240 | 20 |
| M2 | 6 | 8 | 10 | 140 | 11.7 | 10.9 | 180 | 15 |
| M3 | 6 | 7 | 8 | 122.5 | 10.2 | 9 | 158 | 13.125 |

[a]See protocol for determination AUC from C0
[b]Equal to AUC0-12h/12
[c]HCTnonstd multiplied by 45/33 (month 1) or 45/35 (month 2)

#### 5 MATERIALS AND METHODS

##### 5.1 Participiants

##### 4.1.1 Number of participants

BRUNO-PIC will prospectively recruit 45 children treated at RCH undergoing kidney, heart or liver transplantation to a single-arm, open label trial.

##### 4.1.2 Inclusion criteria

- 1- 18yo
- Kidney, cardiac or liver transplant recipients (planned or on waiting list) excluding repeat liver transplant.
- Amenable to venepuncture and blood draw

##### 4.1.3 Exclusion criteria

- > than 18yo
- < than 1yo
- Previous liver transplant.
- Lung OR Intestinal transplant.
- Insufficient time before transplant for pharmacogenomic analysis.
- Known hypersensitivity to tacrolimus and/or its formulation.
- Slow-release preparation of Tacrolimus (e.g Advagraf XL Brand)
- 

##### 4.1.4 Subject enrollment (phased)

BRUNO-PIC involves a single, prospective open label treatment arm, with retrospective standard of care comparator.

45 patients will be recruited prospectively to the trial, with phased opening of cohorts based on SOT type:

**Phase 1**: Enrolling renal transplant recipients, being the population in whom the PPK model used by NextDose was developed.

**Phase 2:** Enrolling other SOT recipients (heart and liver).

- This second phase will commence following validation of the adjusted NextDose model for nonrenal organ transplant recipients. This will involve two steps.
    - First, using published popPK literature to inform PK parameter quantitative differences (and any other necessary changes), Prof Nick Holford will work to develop the necessary parameter adjustments for application of the NextDose tacrolimus model to liver and cardiac transplant recipients.
        - This will primarily be based on already developed meta-models across organ types {Itohara, 2022 #76;Nanga, 2019 #75} . In addition, our group has extracted all published popPK models of tacrolimus in solid organ transplantation. This includes:
            - 43 published popPK models in kidney transplant recipients (37 adult, 6 paediatric)
            - 21 published popPK models in liver transplant recipients (20 adult, 11

paediatric)
- 8 published popPK models in cardiac transplant recipients (7 adult, 1 paediatric)

- Next, external evaluation of the popPK literature derived adjustments to the NextDose Model for use with cardiac and liver recipients will be performed using a Bayesian forecasting procedure on the retrospective data[57] from the prior 5 years in RCH liver and cardiac transplant recipients.

    - For each retrospective subject, the first measured tacrolimus concentration after transplantation will be predicted based on the actual dose administered, popPK standard values and covariates. The second measured tacrolimus concentration after transplantation will then be predicted using continued dosing information, updated covariate values and revised individual pharmacokinetic parameters by balancing population standard values and observed concentrations in the individual. This will be continued for the *n+1*$^{th}$ measured concentration, using information from the *first to n*$^{th}$ concentration.

    - The ability of the transplant-organ specific model to predict the next tacrolimus concentration will then be assess by calculating the median percentage prediction error (MPE%, measure of bias and imprecision)[89] per below equation, whereby Concpred is the model predicted concentration and Concobs is the observed concentration:

$$MPE\% = median(100\% \times \frac{(Conc_{pred} - Conc_{obs})}{Conc_{obs}}$$

- Predictive performance will be assessed over the first 3 post-transplant weeks using retrospective data. The 95% confidence intervals (CI) of the MPE% were will be generated from the 2.5th to 97.5th percentile of the medians in 10 000 non-parametric bootstrap replicate datasets.

##### 4.1.5 Recruitment & consent

Patients and parents of patients in the prospective cohort will be informed of the study by the Study PI assigned to this study following a screening of potentially eligible patients from the electronic medical record and weekly/monthly multi-disciplinary meetings with treating transplant teams.

Patients and their parents will be approached during routine outpatient appointments or, on the ward, if currently an inpatient. Patients and their parents will have the study explained to them and any questions will be answered. A screening of eligibility will be carried out and following this the parent will be left with a copy of the PICF.

There will be 2 arms to this study, a retrospective arm cohort compared with the prospective arm cohort. The approach to informed consent for BRUNO-PIC is as follows:

- **Prospective Cohort Consent**

In accordance with the National Statement, this study will utilize an active consent process for the prospective arm. This will involve presenting potential participants with all the information they need to make an informed decision about participation. When the eligible participant or parent/guardian is invited to participate, they will be provided with detailed information on the purpose of the study, the extent of their involvement, the risks associated and the option to refuse participation at any time during the study. They will be informed that enrolment into the study is a voluntary process.

- Patients and patients of patients will be informed of the study with an initial screening during their inpatient stay, or outpatient appointment, and left with a copy of the patient/parent information statement.
- Prior to performing any trial-specific procedure (including screening procedures to determine eligibility), a signed consent(s) form will be obtained for each participant.
- The investigator or delegated member of the study team will discuss the study with relevant family members: parent/legal guardian and where appropriate the adult participant.
- The investigator will provide the Participant Information and Consent Form to the parent/legal guardian and, where appropriate, to the child/adolescent. This document will describe the purpose of the study, the procedures to be followed, and the risks and benefits of participation.
- Potential participants as well as enrolled participants will be able to contact the study research assistant anytime and have the ability to ask questions and request clarifications at any time prior to or during the study via email.
- Participants who successfully pass the eligibility criteria will proceed through the consent process.
- Where the patient is < 12 years of age, the patient will be provided with the Child Information form and the parent/guardian will be provided with the parent/guardian consent. The Parent/guardian will be asked to read and sign the study informed consent (PICF) if they agree and are willing for their child to participate.
- Where the patient is >12 years of age, the patient will be provided with the Participant PICF. An adolescent information sheet will also be provided. Both parents/guardians and patient will be asked to read and sign the respective consents (PICFs) if they agree and are willing to participate.
- A copy of the signed study consent will be given to the parent guardian or participant upon completion of the consent form. The participant will be considered 'enrolled' from this point and the study interventions will begin thereafter.

- After consenting to participate, the study coordinator will verify the patient's demographics and medical history using the EMR or alternatively by asking the participant if the information cannot be determined from their medical record.

- **Retrospective Cohort: waiver of consent**

- No consent is required as per the National Statement on Ethical Conduct in Human Research as it is considered low risk in this arm.

- A waiver of the consent to be applied as it impracticable to obtain an individual's explicit consent to the use of their information for the purpose of this research. Tacrolimus doses and concentrations will be assessed over a 12-week period in a retrospective manner from patients who have had a SOT from the previous 5 years at RCH from the data that ethics approval is given.

- The RCH is a major base for kidney transplantation and the national base for liver and cardiac transplant patients and caters for patients needing transplant from all over Australia. It is impractical to reach out to these patients who may no longer reside in Melbourne in whom the results will be of no benefit to the patient. The benefit from this study will not cause harm to the patient hence can justify the need to not require consent.
- Patient will not be identifiable when results are analysed. These results will have no impact on patients' current health. It is purely from a collection data to compare with our prospective data.
- Study coordinators may not need to go back this far and will stop once the target review of 45 retrospective patients is met.

#### 5.2 Intervention

##### 4.2.1 Pre-emptive CYP3A4/5 genotyping

Methodology of genotyping and phenotype determination

(i) Genotyping

Patients in the prospective (intervention) arm will undergo genotyping using Illumina's genome-wide genotyping array (Infinium Global Screening Array). The results for the CYP3A4 and CYP3A5 genes will be automatically extracted. Genotype will be assigned per CPIC and other relevant literature, as follows:

- Genotype CYP3A4: Normal metaboliser *1/*1
- Genotype CYP3A4: Poor metaboliser *22
- Genotype CYP3A5: Extensive expresser *1/*1 or *1/*3
- Genotype CYP3A5: Normal expresser *3/*3

(ii) Assignment of phenotype

The determined diplotypes for CYP3A5 will be matched with predicted phenotypes using the CPIC proposed genotype-to-phenotype translation table. The assignment of the phenotype is outlined in the CPIC guidelines. In addition, the influence of CYP3A4 will be incorporated based on recent literature and interventional trials, see Appendix 1 for detail.

###### 4.2.2 BRUNOPIC 1st dose

Initial dose in BRUNO-PIC will use allometric size scaling from adult dose, with adjustment based on genotype (CYP3A4 & CYP3A5).

The first dose calculation is performed using **BRUNOPIC_1stDOSE.xlxs**. The calculations are based on **the following:**

1. In kidney transplantation, dose will be scaled from a dose of 5 mg twice daily in a 70 kg adult.
2. Size scaling by allometry, with fat free mass (FFM) as size descriptor, using this equation:

$$DoseRate_{Child} = DoseRate_{Adult} \times \left( {}^{FFM_{Child}}/_{FFM_{STD}} \right)^{3/4}$$

*where $FFM_{STD}$ is 56.1 kg, based on a 70 kg 176 cm male adult.*

- See **Allometric scaling** for basis for size scaling.

3. After size scaling, proportional adjustment based on genotype (see **Genotype**) is:
   - CYP3A4 *1/*1 normal metabolizer & CYP3A5 *3/*3 non-expresser: No change
   - CYP3A4*22 expresser: Dose x 0.74
   - CYP3A5*1 expresser (hetero- *1/*3 or homozygous *1/*1): Dose x 1.59
   - Both CYP3A5*1 & CYP3A4*22: Dose x 1.18

Tacrolimus is dosed twice daily, around 8am and 8pm. The BRUNOPIC first dose should commence following the transplantation as soon as able and/or consistent with unit guidelines. In living donor kidney transplantation tacrolimus commences on day-3, and thus if available the BRUNOPIC first dose may commence earlier (note the dose immediately prior to transplant is 50% of maintenance dose, given the known concentration-dependent vasoconstrictive effect of tacrolimus on renal microvasculature[90]).

###### 4.2.3 BRUNOPIC Dose adjustment & Target concentration

- **NextDose implementation**

***Bayesian dosing using NextDose will commence after dosing day 4 of tacrolimus post-transplant***

***dosing (DD4)***, on which day the first limited PK profile will be performed. Notably, DD0 is used to mean the first day of *post-transplant tacrolimus dosing*. In kidney transplantation, DD0 is also the day of transplantation (which is designated DTX0). This difference is stipulated as post-transplant tacrolimus dosing (DD0) is delayed for several days post day of transplant (DTX0) in liver transplant recipients.

Tacrolimus concentrations will be measured in routine biochemistry samples per unit protocol. In kidney transplantation these are typically drawn around 6:00 am ("daily trough concentrations"). Tacrolimus concentrations are measured daily for the first month post kidney transplant (prior to the morning dose), and then thrice weekly for the second month. Additional tacrolimus concentrations will be measured for BRUNOPIC participants, on D4 post-transplant, and then at 3 weeks post-transplant and 8 weeks post-transplant. These samples will be the time of morning dose, and then 1, 2 and 3h post dose. The time of sampling (to the closest minute) should be recorded. Samples should be sent to the lab for measurement after each sample is taken.

Notably, Bayesian AUC estimation using a limited sampling strategy out to 3h post dose has been shown accurate for tacrolimus.[78,79,82]

Patient parameters along with a unique identifier will be entered into NextDose, along with the tacrolimus dosing and concentrations. The predicted dose as per NextDose will be communicated with the transplant treatment team via phone or in-person. The tacrolimus dose will be pending on EMR and documented. Tacrolimus concentrations will then be taken at the specified time intervals in addition to the standard testing as per protocol. NextDose implementation and dosing recommendations will be led by academic pharmacist and CRA in consultation with PI Dr David Metz.

- **Target concentrations (and derivation) for tacrolimus**

##### 5.2.1 Concomitant therapy

There are several medications that can interact significantly if taken concomitantly with tacrolimus. This may require dose adjustments as per evidence-based recommendation (see Appendix 6). Subsequently, NextDose predictions will be refreshed (i.e. concentrations prior to major DDI not used) whilst the interacting medication is having impact.

### 6 TRIAL VISITS AND PROCEDURES

#### 6.1 Trial timeline

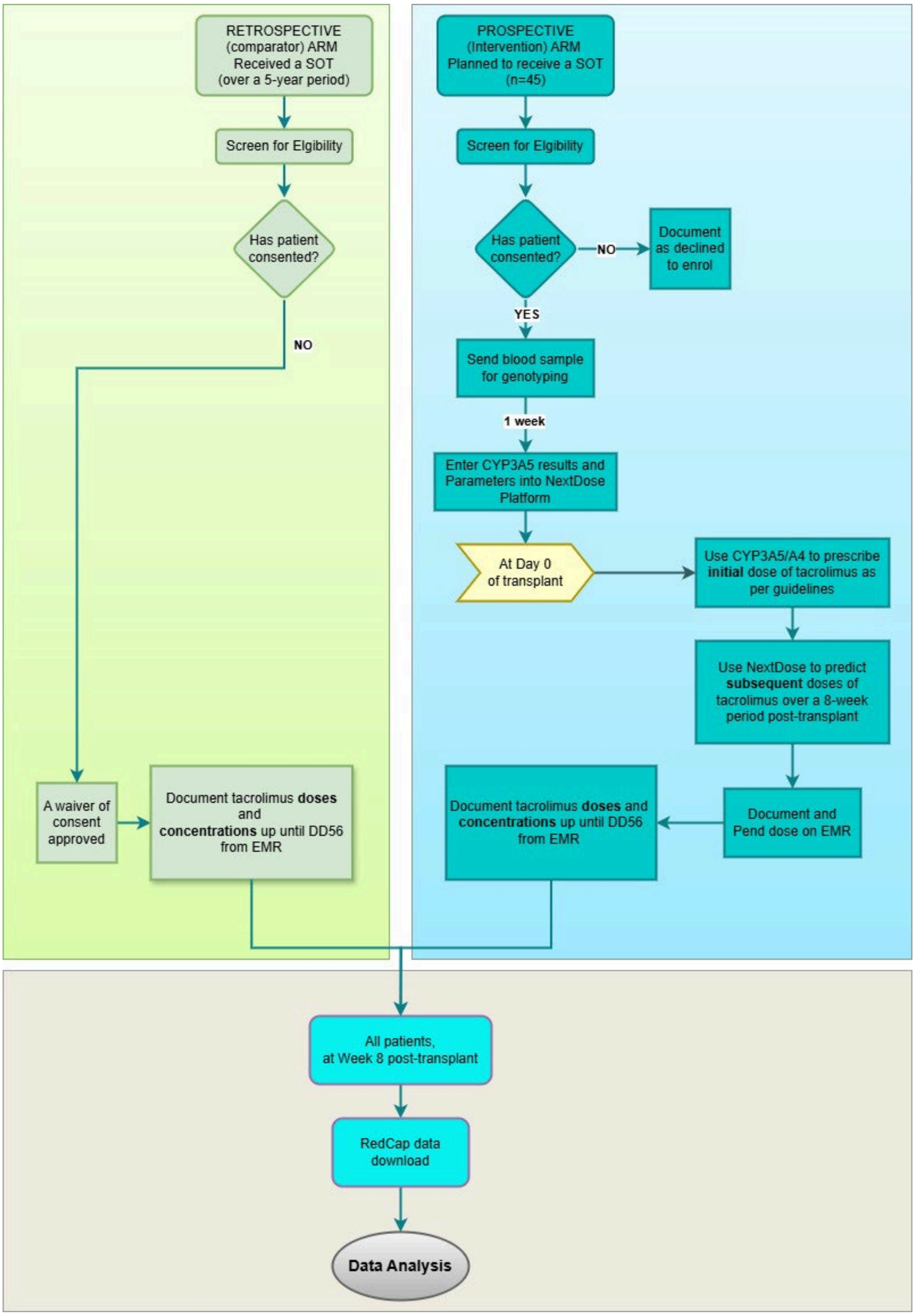

**7**

* For Retrospective arm: Tacrolimus doses, adjustments and concentrations will be taken from the EMR. If data is missing at those time points, +2day (on D4) or ±7days (from D4 to D56) will be use. Any time points not within this range will be deemed as “lost to follow-up”. NOTE: DD0 is the first dosing day of tacrolimus -. For kidney transplantation, DD0 will be same as DT0

### For Prospective arm: Tacrolimus doses, adjustments and concentrations will be recorded from the EMR at set time points and as per transplant policy.

##### 7.1 Schedule of assessments

| Evaluation | Screening and Eligibility | Consent and Enrolment | DTO | DT1 | DT3 | DT4 | DT21 | DT56 |
|---|---|---|---|---|---|---|---|---|
| TIME POINT at Week | **-4** | **-4 to -1** | | **1** | | **1** | **3** | **8** |
| Eligibility criteria | **X** | | | | | | | |
| Informed Consent | | **X** | | | | | | |
| Baseline Data | | **X** | | | | | | |
| Medication History | | **X** | | At all timepoints | | | | |
| Blood Test for PGx testing* | | **X** | | | | | | |
| 8 hourly Tacrolimus level on D3 | | | | | **X** | | | |
| Additional* Tacrolimus levels on D4, W3 and W8: 4-point AUC; trough, +1 hr, +2hr, +3hr post tacrolimus dose | | | | | | **X** | **X** | **X** |
| Add new patient to NextDose and REDCap database | | **X** | | | | | | |
| Record observations and tacrolimus concentration# | | | | | Daily from DT1 to DT56 | | | |
| Record tacrolimus dose given# | | | | | Daily from DT1 to DT56 | | | |

| | | | | | | |
|---|---|---|---|---|---|---|
| Use NextDose to predict tacrolimus dose * | | | | | | Daily from DD4 to DD56 |

#For Retrospective arm: Tacrolimus doses, adjustments and concentrations will be taken from the EMR. If data is missing at those time points those timepoints will be deemed as “lost to follow-up”.
For Prospective arm: Tacrolimus doses, adjustments and concentrations will be recorded from the EMR at set time points and as per transplant policy. This includes daily tacrolimus concentrations in the early post-transplant weeks. Additional tacrolimus concentrations will be taken on at D3,D4, W3 and W8:
8-hourly on D3 and 4-point AUC on others will be performed alongside mycophenolic acid AUC, where collected as part of clinical care.
NOTE: DT0: Day post-transplant 0 (day 0 is the day of transplant), DD0 post-transplant tacrolimus Dosing Day 0 (day 0 is the first dosing day)

*Performed on prospective arm upon enrolment by the academic pharmacist in conjunction with the PI.

#### 7.2 Description of procedures

##### 7.2.1 Patient Eligibility (Inclusion/exclusion criteria)

Refer to section 4.4 Eligibility criteria and 4.6 Recruitment and identification of potential participants.

##### 4.3 Baseline Data collection

| Baseline characteristics collected upon study enrolment |
|---|
| Age |
| Gender |
| Weight and height |
| Ethnicity |
| Transplant details: type of transplant, time post-transplant, primary diagnosis, donor (living or post-mortal). |
| Laboratory Measurements: hematocrit, creatine and estimated eGFR, albumin, LFTs. |

##### 7.2.2 Medication History

There are several medications that can interact significantly if taken concomitantly with tacrolimus. This may require dose adjustments as per evidence-based recommendation. A thorough medication history needs to be taken at the set time intervals when tacrolimus concentrations and doses are being calculated. Medication history can be taken from the EMR and confirmed with patient and or their carer.

- **Blood Test for PGx testing**

All participants will undergo venepuncture or central line access and a blood draw (3mL for all patients, minimum volume 1mL) to be done by a trained healthcare professional.
Blood sample sent to VCGS laboratories for targeted sequencing then a clinical report will be generated within 1-2 weeks by a Bioinformatician.

- **Blood Test for tacrolimus concentration**

All participants will undergo venipuncture or central line access and a blood draw for monitoring tacrolimus concentrations. All tacrolimus concentrations collected are as part of standard of care, apart from the additional samples on days 4 and weeks 3 and 8, which are performed at times blood is taken as part of clinical care already, with addition of tacrolimus measurement only. Thus, no venipuncture occurs beyond standard of care, with additional samples taken either via central line (DD4) or at times samples are being taken already as part of standard care.

Typically, blood testing will occur daily during the first 1-2 weeks post-transplant and occur less often (i.e., weekly) for a period of time. The frequency and timing of tacrolimus blood test monitoring will depend on the type of organ transplanted, the transplant protocol, and individual patient factors.

Tacrolimus concentrations are measured daily for the first month post kidney transplant (prior to the morning dose), and then thrice weekly for the second month.

Additional tacrolimus concentrations will be measured for BRUNOPIC participants, on D4 post-transplant, and then at 3 weeks post-transplant and 8 weeks post-transplant. These samples will be the time of morning dose, and then 1, 2 and 3h post dose, with addition of 12h post dose on D4 (whilst inpatient).

  - For weeks 3 and 8 these will be timed with standard care (MPA profiles taken at these times).
  - On D4, these are in addition to standard care, taken from in situ central line.
  - One final sample on D4 will be taken at ~12h post dose, when bloods are taken as part of clinical care.

Notably, Bayesian AUC estimation using a limited sampling strategy out to 3h post dose has been shown accurate for tacrolimus.[78,79,82]

##### 7.3 Treatment discontinuation, participant withdrawals and losses to follow up

Participants and or their parent/legal guardian can withdraw consent for trial participation at any time during the study. We do not anticipate any loss to follow-up as all transplant recipients are followed closely in the 1st three months post-transplant by their treating units.

###### 7.3.1 Withdrawal of consent - participant withdraws from all trial participation.

Patients or parents of patients can withdraw their child from the study at any time. Patients or parents of patients who have previously consented to the study can withdraw from the study by notifying the study PI or research assistants from the team

Withdrawing from the trial will not affect their access to standard treatment or their relationship with the hospital and affiliated health care professionals.

###### 7.3.2 Study Closure

A participant is considered to have completed the trial if he or she has completed all phases of the trial including the last visit or the last scheduled procedure shown in the Schedule of Assessments.

The end of the trial is defined as completion of the last visit or procedure shown in the Schedule of Assessments in the trial at all sites. At this stage, the Sponsor-Investigator will ensure that all HRECs and RGOs are noted.

###### 7.3.3 Temporary halt or early termination of a trial

This trial may be temporarily suspended or prematurely terminated if there is sufficient reasonable cause. If the trial is prematurely terminated or suspended, the Principal Investigator will promptly inform trial participants, HREC and RGO, the funding and regulatory bodies, providing the reason(s) for the termination or suspension. Circumstances that may warrant termination or suspension include, but are not limited to:

- Determination of an unexpected, significant, or unacceptable risk to participants that meets the definition of a Significant Safety Issue (SSI).
- Insufficient compliance to protocol requirements.
- Determination that the primary endpoint has been met.
- Data that is not sufficiently complete and/or evaluable.

In the case of concerns about safety, protocol compliance or data quality, the trial may resume once the concerns have been addressed to the satisfaction of the sponsor, HREC, RGO, funding and/or regulatory bodies.

#### 8 SAFETY MONITORING AND REPORTING

Study safety monitoring will be coordinated by the PI and the study coordinator. These reported adverse events will be assessed and classified individually and in consultation.

Major risks in undertaking a clinical trial can be broadly categorised into:

- ***PART 1*** *- Risks to the safety and rights of the study participants (the risks and other unintended effects of trial interventions or trial conduct)*

- ***PART 2*** *- Risks to the successful conduct of the study (e.g. inadequate funding, poor recruitment, poor quality data/samples, inadequate accountability of the investigational product).*

##### 7.1 Definitions

<u>Adverse Event (AE):</u> Any untoward medical occurrence in a patient or clinical trial participant administered a medicinal product and does not necessarily have a causal relationship with this treatment.

Adverse Drug Event (ADE): An injury resulting from the use of a drug. This includes harm caused by the drug (adverse drug reactions and overdoses) and harm from the use of the drug (including dose reductions, discontinuations of drug therapy and interactions)

Serious Adverse Event (SAE): Any adverse even reaction that results in death, is life threatening, requires hospitalisation or prolongation of existing hospitalisation, results in persistent or significant disability or incapacity or is a congenital anomaly or birth defect.

Note: Life-threatening refers to an event in which the participant was at risk of death at the time of the event. It does not refer to an event that hypothetically might have caused death if it were more severe. Medical and scientific judgement should be exercised in deciding whether an adverse event should be classified as serious in other situations. **Important medical events** that are not immediately life-threatening or do not result in death or hospitalisation but may jeopardise the participant or may require intervention to prevent one of the other outcomes listed in this definition should also be considered serious.

Suspected Unexpected Serious Adverse Reaction (SUSAR): An adverse reaction that is both serious and unexpected.

Significant Safety Issue (SSI): A safety issue that could adversely affect the safety of participants or materially impact on the continued ethical acceptability or conduct of the trial.

Urgent Safety Measure (USM): A measure required to be taken in order to eliminate an immediate hazard to a participant's health or safety. Note: This is a type of SSI that can be instigated by either the investigator or sponsor and can be implemented before seeking approval from HRECs or institutions.

##### 7.2 Capturing and eliciting adverse event/reaction information

All AEs or ADEs, whether or not related to the prospective study arm will be collected by the PI, academic pharmacist or research assistant during the scheduled time points of tacrolimus data collection, as well as at any time point during the study where the participant or their representative contacts the Study Team to report such an event. The study team will record the details of each AE including, but not limited to, information such as date of event and duration of event. AEs or ADEs will be followed up until resolution or stabilisation, or 30 days after the completion of the patient's last visit – whichever occurs earlier.

- **Adverse events related to this study.**

The following occurrences *are* regarded as AEs:

- Bruising or due to having blood test performed.
- Vomiting or spitting out of tacrolimus (within 2 hours of administration).
- Allergic reaction to tacrolimus (within 2 hours of administration).
- High tacrolimus concentrations and/or toxicity symptoms as a result of NextDose predicting a higher tacrolimus dose. Signs of toxicity can include nephrotoxicity, nausea, tremors, and elevated liver enzyme levels.

Side-effects of Tacrolimus

| **Common (> 1%)** | **Infrequent (0.1 – 1%)** | **Rare (< 0.1%)** |
|---|---|---|

| | | |
|---|---|---|
| Nephrotoxicity*, hypertension, hypercholesterolaemia, neurotoxicity#, raised bilirubin, raised aminotransferases, hypomagnesaemia, hyperkalaemia, opportunistic infection, diarrhoea, hyperglycaemia, diabetes | muscle weakness, myopathy | haemolytic uraemic syndrome |

*dose-related and reversible with dose reduction or withdrawal

### Tremor and headache are most frequent; other effects include paraesthesia, confusion, seizures, coma, psychosis. Reversible posterior leukoencephalopathy syndrome has also been reported.

The following occurrences *are not* to be regarded as AEs:

- Underlying (pre-existing) symptoms or diseases unless there is an increase in severity or frequency during the course of the investigation.
- Conditions that are present at screening and do not deteriorate will not be considered adverse events.
- Abnormal laboratory values will not be considered adverse events unless deemed clinically significant by the investigator and documented as such.

All adverse events are subject to clinical and scientific judgment and will be subject to assessment as per the TMG.

- **Documentation of AEs**

For the purposes of this study the investigator is responsible for recording all Adverse Events, regardless of their relationship to trial drug, with the following exceptions:

- Conditions that are present at screening and do not deteriorate will not be considered adverse events.
- Abnormal laboratory values will not be considered adverse events unless deemed clinically significant by the investigator and documented as such.

The AE will be described in the source documents (e.g. medical record) and captured on the REDCap database and will include:

- A description of the AE
- The onset date, duration, date of resolution
- Severity (mild, moderate or severe and with reference to the Common Terminology Criteria for Adverse Events (CTCAE v.5) produced by the National Cancer Institute (U.S) https://ctep.cancer.gov/protocoldevelopment/electronic_applications/ctc.htm

AND

The severity of an Adverse Event will be assessed as follows:

- **Mild**: Events that require minimal or no treatment and do not interfere with the participant's daily activities.
- **Moderate**: Events that cause sufficient discomfort to interfere with daily activity and/or require a simple dose of medication.
- **Severe:** Events that prevent usual daily activity or require complex treatment.
    - Seriousness (i.e., is it an SAE?).
    - Any action taken, (i.e., treatment, follow-up tests).

- The outcome (recovery, death, continuing, worsening) and management as per study.

- **Assessing the seriousness of a participant's AE**

The seriousness of an AE will be assessed by an investigator according to the definition in in the preceding section on definitions with the following exception(s):

- Hospitalisation due to progression of disease will not be considered an SAE for the purposes of this trial.

The seriousness of an AE will be assessed by an investigator according to the definition in Section 8.1, with the following exceptions:

- Hospitalisation due to progression of disease will not be considered an SAE for the purposes of this trial.
- Elective surgery planned at the time of enrolment.

- **Assessing the relatedness (causality) of a participant's AE**

The relationship of the event to the BRUNO-PIC study will be assessed as follows:

- **Unrelated:** There is no association between the BRUNO-PIC study and the reported event. AEs in this category do not have a reasonable temporal relationship to exposure to the test product or can be explained by a commonly occurring alternative aetiology.
- **Possible:** The event could have cause or contributed to the AE. AEs in this category follow a reasonable temporal sequence from the time of exposure to the intervention and/or follow a known response pattern to the test article but could also have been produced by other factors.
- **Probable:** The association of the event with BRUNO-PIC study seems likely. AEs in this category follow a reasonable temporal sequence from the time of exposure to the test product and are consistent with the known pharmacological action of the drug, known or previously reported adverse reactions to the drug or class of drugs, or judgement based on the investigators clinical experience.
- **Definite:** The AE is a consequence of administration of the BRUNO-PIC study. AEs in the category cannot be explained by concurrent illness, progression of disease state or concurrent medication reaction. Such events may be widely documented as having an association with the test product or that they occur after rechallenge.

- **Assessing the expectedness of a participant's AE**

The TMG will be responsible for determining whether an AE is expected or unexpected. An AE will be considered unexpected if the nature, severity, or frequency of the event is not consistent with the risk information previously described for the study intervention.

##### 7.3 Report of safety events

The Principal Investigator is responsible for reporting SAEs (including SUSARs) to the Sponsor-Investigator as soon as possible but within 24 hours of the first knowledge of the event. These reports should be submitted using the trial Expedited Safety Report Form (Appendix 2) and emailed to. Adverse effects related specifically to this study are outlined in section 7.3.

#### 9 DATA AND INFORMATION MANAGEMENT

BRUNO-PIC results will be kept on a REDCap database. This is a secure database run by the Murdoch Children's Research Institute. All paper files and consents will be kept locked away securely in a research office. The Principal Investigator is responsible for storing essential trial documents relevant to data management and maintaining a site-specific record of the location(s) of the site's data management-related Essential Documents.

The Principal Investigators will also maintain accurate case report forms (CRFs) (i.e. the data collection forms) and be responsible for ensuring that the collected and reported data is accurate, legible, complete, entered in a timely manner and enduring. To maintain the integrity of the data, any changes to data (hardcopy and electronic) must be traceable, must not obscure the original entry, and must be explained where this is necessary.

Any person delegated to collect data, perform data entry or sign for data completeness will be recorded on the delegation log and will be trained to perform these trial-related duties and functions.

##### 4.4 8.2 Data management

Data will be collected in an identifiable manner. Participants will then be allocated a unique individual identifier prior to their de-identified data being added to the database. If any data is removed from the REDCap database the patient identifiable information will be removed and the unique individual identifier will be applied to the data prior to extraction.

Thus, although the information entered will be de-identified it will be re-identifiable to the researchers involved. Only the PI and AI's of the study will have access to re-identifiable information of individuals in the database for data entry purposes.

All data is protected within Murdoch Children's Research Institute using the REDCap database by a triple encryption process between the remote user, the server and the REDCap Database. No data can leave the database in an identifiable format. All data extraction is de-identified for all users at all times, thus privacy and confidentiality will be maintained.
Database information collection Data generation (source data)

In this trial, the following types of data will be collected:

- Study Number
- Initials of patient
- Year of birth
- Ethnicity
- Height and weight
- Email details for correspondence with participant/parent
- 
- SOT type
- Donor status
- CYP3A4 and 3A5 genotype
- ComorbiditiesTacrolimus concentrations at set time points
- Tacrolimus dose at set time points
- NextDose tacrolimus dose calculations
- Urea, Electrolytes & creatinine, liver function tests and Haematocrit at each timepoint if performed as part of routine clinical care.
- Medication reconciliation at each study time point

- **Source Document Plan**

The source documents for this trial include:
- RCH EMR
- Laboratory reports
- Signed parent/guardian (and, where applicable participant) information and consent forms.

Will be signed and dated by the Principal Investigator and will be prepared prior to recruitment of the first participant. It will be available from the Investigator Site File.

- **Use of the data**

The data will be used for the analyses specified in the protocol and Statistical Analysis Plan.

Following the completion and analysis of the trial, the data will be retained long-term following the mandatory archive period for use in future research projects.

- **Storage and access**

Hard copy data will be stored in a locked cabinet in a secure location, accessible to the research team only. Electronic data will be securely stored in MCRI's REDCap database system and in files stored in MCRI's network file servers, which are backed up nightly. Files containing private or confidential data will be stored only in locations accessible only by appropriate designated members of the research team.

REDCap is hosted on MCRI infrastructure and is subject to the same security and backup regimen as other systems (e.g. the network file servers). Data is backed up nightly to a local backup server, with a monthly backup taken to tape and stored offsite. REDCap maintains an audit trail of data create/update/delete events that is accessible to project users who are granted permission to view it. Access to REDCap will be provided via an MCRI user account or (for external collaborators) via a REDCap user account created by the MCRI system administrator. The permissions granted to each user within each REDCap project will be controlled by, and will be the responsibility of, the trial team delegated this task by the Principal Investigator. REDCap has functionality that makes adding and removing users and managing user permissions straightforward. All data transmissions between users and the REDCap server are encrypted. The instructions for data entry to REDCap must be read and the training log signed prior to personnel commencing data entry on REDCap.

Authorised representatives of the sponsoring institution as well as representatives from the HREC, Research Governance Office and regulatory agencies may inspect all documents and records required to be maintained by the Investigator for the participants in this trial.

- **Disclosure**

The trial protocol, documentation, data and all other information generated will be held in strict confidence. No information concerning the trial or the data will be released to any unauthorised third party, without prior written approval of the sponsoring institution. Clinical information will not be released without written permission of the participant, except as necessary for monitoring by the HREC, Research Governance Office or regulatory agencies.

- **Archiving - Data and document retention**

All data stored on the REDCap database will be archived for at least 15-years post-trial completion (TGA) or until child aged 25 years (whichever is the later). All genetic data will be stored within a dedicated VCGS network drive to store this data with access to it. Those that have access will be the PI together with the bio-informatician of the Pharmacogenomics Team. This will be stored until the completion of the study. After study completion, data will be retained in a secure offline archive utilising MCRI data backup infrastructure, to comply with publication and research data retention requirements and also to enable future reuse in accordance with patient consent.

- **Destruction**

All hardcopy and electronic data will be destroyed in accordance with the most up to date Australian National statement on data and regulation policies.

##### 8.2 Data confidentiality

Participant confidentiality is strictly held in trust by the Principal Investigator, participating investigators, research staff, and the sponsoring institution and their agents. This confidentiality is extended to cover testing of biological samples and genetic tests in addition to the clinical information relating to participating participants.

To preserve confidentiality and reduce the risk of identification during collection, analysis and storage of data and information, the following will be undertaken:

**(1)** The number of private/confidential variables collected for each individual has been minimised. The data collected will be limited to that required to address the primary and secondary objectives.

**(2)** Participant identifiers will be stored separately to the data collected; documents with identifiers will be stored separately to participant data. Participant data and samples will be identified through use of a unique participant trial number/code assigned to the trial participant ("re-identifiable"). The Site Principal Investigator is responsible for the storage of a master-file of names and other identifiable data with the participant ID; access to this document will be restricted to the site trial team and authorised persons as listed previously. The master file should be stored securely, and separately, from trial data in locked/ password-protected databases with passwords kept separately.

**(3)** Separation of the roles responsible for management of identifiers and those responsible for analysing content. The data will be analysed by the statistician, who will be provided with anonymised data identified only by the unique participant trial ID.

##### 8.3 Quality assurance

The PI will have responsibilities in relation to quality management. The PI will develop SOPs that identify, evaluate and control risk for all aspects of the trial, (i.e. trial design, source data management, training, eligibility, informed consent and adverse event reporting). Quality control (QC) procedures, which will include the data entry system and data QC checks will also be implemented. Any missing data or data anomalies will be communicated for clarification/resolution.

###### 8.1.1 Data sharing

Results may also be presented publicly in the form of scientific talks or publication in internationally available journals. This may include publication of information including age, family history, physical description relative to their disorder (including images) diagnosis and treatment. Only information relevant to the disorder will be published and in this case the patient will also be coded and unidentifiable.

### 10 TRIAL OVERSIGHT

#### 9.1 Governance oversight

##### 10.1.1 Trial Management Group (TMG)

The Principal Investigator is responsible for supervising any individual or party to whom they have delegated tasks at the trial site. They must provide continuous supervision and documentation of their oversight. To meet this GCP requirement, a small group will be responsible for the day-to-day management of the trial and will include at a minimum the PI, Academic Pharmacist and Research Assistant. The group will closely review all aspects of the conduct and progress of the trial, ensuring that there is a forum for identifying and addressing issues. Meetings must be minuted with attendees listed, pertinent emails retained, and phone calls documented.

##### 10.1.2 Pharmacogenomic Steering Committee (PSC)

The Pharmacogenomics Steering Committee will meet every 2 to 4 weeks to review the pharmacogenomic data for patients enrolled in the study. This group will consist of the Study PI, Academic Pharmacist, Clinical Pharmacologist (as required) and Bioinformatician. The meeting will serve the purpose of generating purpose-built clinical reports of pharmacogenomic results.

#### 10.2 Site Monitoring

Trial site monitoring is conducted to ensure that the rights and well-being of trial participants are protected, that the reported trial data is accurate, complete, and verifiable, and that the conduct of the trial is in compliance with the current approved protocol and amendment(s), good clinical practice and applicable regulatory requirements.

Full details of trial site monitoring are documented in the Clinical Monitoring Plan (CMP). The CMP describes in detail who will conduct the monitoring, at what frequency monitoring will be done, at what level of detail monitoring will be performed, and the distribution of monitoring reports.

Monitoring for this trial will be performed by a designated trial study coordinator. On-site monitoring will be conducted after the enrolment of 5 patients initially, and throughout the trial thereafter. Monitoring will involve review of 100% of original signed consent forms, trial eligibility data and data related to primary outcome, safety and other key data variables; review of all withdrawals from trial treatment and/or trial follow-up.

The study coordinator will provide direct access to all trial related source data/documents, and reports for the purpose of monitoring and auditing by the sponsor, and inspection by local and regulatory authorities.

#### 10.3 Quality Control and Quality Assurance

The Sponsor-Investigator will have responsibilities in relation to quality management. The Sponsor-Investigator will develop SOPs that identify, evaluate and control risk for all aspects of the trial (i.e. trial design, source data management, training, eligibility, informed consent and adverse event reporting). The Sponsor-Investigator will also implement quality control (QC) procedures, which will include the data entry system and data QC checks. Any missing data or data anomalies will be communicated for clarification/resolution.

As outlined in the previous section (9.2 Site Monitoring), the trial monitor will verify that the clinical trial is conducted and data is generated, documented (recorded), and reported in compliance with the protocol, good clinical practice and applicable regulatory requirements.

In the event of non-compliance that significantly affects human participant protection or reliability of results, the Sponsor-Investigator will perform a root cause analysis and corrective and preventative action plan (CAPA).

In addition, the study coordinator will perform internal quality management of trial conduct, data and biological specimen collection, documentation and completion. An individualised quality management plan will be developed to describe a site's quality management.

### 11 STATISTICAL METHODS

#### 11.1 Sample Size Estimation and statistical analysis plan

The sample size was calculated based on comparing proportion of patients within each group with a $C_0$ concentration within therapeutic range on day 4 post transplantation., Prior studies in adult kidney transplant recipients have shown 54% vs 24% [14], 54.8% vs 20.8% [91] and 58% vs 37.4% [12] of patients had a tacrolimus $C_0$ within target range on Day 3-5 when treated with the standard, bodyweight-based dosing vs a dosing algorithm.We assumed 27% of the retrospective control cohort would have a tacrolimus $C_0$ within target range on DD4. 165 participants (120 control and 45 intervention participants) would provide us 80% power to detect a risk difference of 24% assuming a two-sided alpha of 0.05.

Outcomes:

Comparison between the intervention prospective arm and the control group (retrospective historical comparator) will be utilised for analyses where comparable data is available . We will use a stabilised inverse probability weighting (IPW) approach with robust standard errors to control for potential bias between the "exposure" groups (i.e. NextDose (intervention) vs retrospective control cohort). IPW is an extension of the propnsity score method used to summarise the conditional probability of assignment to an exposure. The weights are the inverse probability of assigning an exposure derived from a logistic model with group as the dependent variable and observed patient-level characteristics as the independent variables. These will include factors that may influence tacrolimus concentrations, including demographic variables (age, sex), use of drugs known to interact with tacrolimus, presence of comorbidities known to influence transplant outcomes, presence of liver dysfunction, height, weight, prednisolone dose in mg, and HCT levels (all measured on day of transplant D0). We will stratify by type of solid organ transplant (heart, liver or kidney). Stabilisation is accomplished by multiplying the "exposure" weights (separately) by a constant, equal to the expected value of being in the intervention or control groups. Each child is weighted by the inverse of the estimated probability of the exposure received. We will then use IPW regression models weighted with exposure to estimate the adjusted associations between exposure and each outcome.

The co-primary outcome analysis will compare"

1. the proportion of participants with a Cssavg concentration within acceptable range

on DD4 between groups will be made using a risk difference, estimated using IPW logistic regression with marginalisation, adjusted for age, gender, SOT type and other factors known to influence tacrolimus concentrations presented with its 95% confidence interval (CI) and p-value. For the primary outcome

2. the proportion of participants with a Cavgss concentration within acceptable range at DD4, week 3 and week 8 will be made using a risk differences estimated using a marginalised IPW mixed-effects logistic regression model including a random effect for the intercept (to allow for clustering of repeated measures within participants), and a fixed effect for dosing type (control vs intervention), time-period (DD4, week3, week8), age, gender and SOT type (stratification factor) and other factors known to influence tacrolimus concentrations.

Ctrough concentrations are only available in the control group, thus Cssavg will be calculated using a maximum *a posteriori* (MAP) approach, using the 4-point tacrolimus concentration profile on days DD4, week3 and week8, participant clinical covariate information and the published popPK model within NextDose.

For secondary outcomes, time to an acceptable Cssavg target we will compare groups using a hazard ratio and 95% confidence interval estimated similarly to primary outcomes via a IPW adjusted Cox proportional hazards model. Between-group difference in mean percentage time within acceptable range over the first 8-weeks will be compared using IPW adjusted linear regression. Proportion within acceptable Ctrough concentration on DD4 will be analysed similarly to the primary outcome, proportion within acceptable Cssavg concentration on DD4.

For secondary outcomes where Cssavg is used, in the control group Cssavg will be calculated using Ctrough concentration by MAP estimation as above.

##### 10.2 Handling of missing data

Any missing data or data anomalies will be communicated to the PIs for clarification/resolution.

#### 12 ETHICS AND DISSEMINATION

##### 12.1 Research Ethics Approval & Local Governance Authorisation

This protocol and the informed consent document and any subsequent amendments will be reviewed and approved by the Sydney Children's Hospitals Network Human Research Ethics Committee (SCHN HREC) prior to commencing the trial. A letter of protocol approval by HREC will be obtained prior to the commencement of the trial, as well as approval for other trial documents requiring HREC review.

A letter of authorisation will be obtained from the RGO prior to the commencement of the research at the Royal Children’s Hospital. Institutional governance authorisation for any subsequent HREC-approved amendments will be obtained prior to implementation.

##### 12.2 Amendments to the protocol

This trial will be conducted in compliance with the current version of the protocol. Any change to the protocol document or Informed Consent Form that affects the scientific intent, trial design, participant safety, or may affect a participants willingness to continue participation in the trial is considered an amendment, and therefore will be written and filed as an amendment to this protocol and/or informed consent form. All such amendments will be submitted to the HREC, for approval prior to being implemented.

##### 12.3 Protocol Deviations and Serious Breaches

All protocol deviations will be recorded in the participant record (source document) and on the CRF and must be reported to the Principal Investigator, who will assess for seriousness.

Those deviations deemed to affect to a significant degree rights of a trial participant or the reliability and robustness of the data generated in the clinical trial will be reported as serious breaches. Reporting will be done in a timely manner (Principal Investigator to report to the Sponsor within 72 hours and to the Site RGO within 7 day; the Sponsor will review and submit to the approving HREC within 7 days).

Where non-compliance significantly affects human participant protection or reliability of results, a root cause analysis will be undertaken and a corrective and preventative action plan prepared.

Where protocol deviations or serious breaches identify protocol-related issues, the protocol will be reviewed and, where indicated, amended.

#### 13 CONFIDENTIALITY

Participant confidentiality is strictly held in trust by the PI, research staff, and the sponsoring institution MCRI and their agents. This confidentiality is extended to cover clinical information relating to participating participants.

The trial protocol, documentation, data and all other information generated will be held in strict confidence. No information concerning the trial or the data will be released to any unauthorised third party, without prior written approval of the sponsoring institution. Authorised representatives of the sponsoring institution may inspect all documents and records required to be maintained by the Investigator, including but not limited to, medical records (office, clinic or hospital) and pharmacy records for the participants in this trial. The Royal Children's Hospital will permit access to such records.

All evaluation forms, reports and other records that leave the site will be identified only by the Participant Identification Number (SID) to maintain participant confidentiality.

Clinical information will not be released without written permission of the participant, except as necessary for monitoring by HREC or regulatory agencies.

#### 14 FINANCIAL DISCLOSURE AND CONFLICTS OF INTEREST

There are identified and declared no financial or other competing interests for investigators for the overall trial and Royal Children's Hospital, Melbourne.

#### 15 DISSEMINATION AND TRANSLATION PLAN

At the end of the project the study team will send a final letter to the participants. This letter will explain what was found during this project – in other words, our project results. The letter will not have any identifiable information or information specific to each participant. MCRI holds the primary responsibility for publication of the results of the study.

### 16 APPENDICES

#### APPENDIX 1: Additional potential risks to participants

| Potential risk | Impact | Likelihood | Mitigation |
|---|---|---|---|
| Bleeding or Infection risk at the site of blood test | High | Unlikely | Only trained and accredited nursing staff to take blood sample to help reduce this risk.<br><br>A numbing cream can be applied on skin to help reduce any discomfort where applicable. In addition, some patients will have a central venous access device in- situ allowing for immediate blood draws. |
| Unable to provide Pharmacogenomic Result | High | Unlikely | Pharmacogenomic variant calling can be difficult. Not all diplotypes are easily identifiable.<br><br>The study team currently has an open PGx trial in paediatric oncology patients (MARVEL-PIC) where 27 gene:drug pairs are examined via whole genome sequencing. To date there has been 100% concordance with genotypes repeated in NATA accredited laboratories (TPMT/NUD 15 genotype) and the team has developed significant experience in calling across a range of metaboliser genes.<br><br>In addition, a standard operating procedure has been developed for how to report variants in the event that a metaboliser state is unclear. This was developed in collaboration with input from senior clinical geneticist Professor Paul James. |
| Data security breach | High | Unlikely | MCRIs data policies and procedures will minimize data security breach concerns. |
| Study staff (PI/RA/PhD Student/Academic Pharmacist) unable to complete enrolment stage. | Medium | Possible | To mitigate this issue, we have multiple staff able to complete the enrolment task. In addition, a SOP will be written to ensure all staff are consistent in completing all tasks.<br><br>Certain tasks can be carried out via electronic platforms (i.e. Zoom) or via |

| | | | |
|---|---|---|---|
| | | | telephone calls which may be enabled particularly if staff are able to work from home.<br>The pharmacogenomics team has a total of 2.2 FTE Academic pharmacists and 1.6 FTE Research Assistants. |
| Internet down | High | Unlikely | A hospital Code yellow is to be announced, and offline backup system can be used or document on paper |
| Computer System (Epic EMR) is offline | High | Unlikely | A hospital Code yellow is to be announced, and offline backup system can be used or document on paper |
| Uncovering other actionable genes responsible for metabolising concomitant medications that has clinical implications | Medium | Unlikely | We will undertake a targeted approach to genotyping the CYP3A5 and CYP3A4 gene and diplotypes therefore it is highly unlikely we will uncover any additional actionable genotypes. |

#### APPENDIX 2: Rationale for initial dose determination

- **Tacrolimus SOC dosing**

**FDA label**

- Adult Kidney Transplant
  - With azathioprine 0.1 mg/kg/dose twice daily
  - With MMF/IL-2RA, 0.05 mg/kg/dose twice daily
- Paediatric Kidney Transplant
  - 0.15 mg/kg/dose twice daily

**Examples, paediatric age-banded dosing:**

- Kausman et al 2008[92]
  - <40 kg–0.15 mg/kg dose twice daily
  - >40 kg–0.1 mg/kg/ dose twice daily (maximum 10 mg/dose).
- Min et al 2018[10]
  - 0.075 mg/kg twice daily for >6yo
  - 0.1 mg/kg twice daily for <6yo
  - 2x dose for expressers
  - Cap dose at 5 mg twice daily.

**Adult contemporary practice**

- In Australian and New Zealand, dosing data from all kidney transplant episodes is collected within a registry, ANZDATA. Analysis of this data shows that the median initial dose of tacrolimus is 5 mg/70 kg/dose twice daily (unpublished from ANZDATA, Metz PhD thesis)[93]
  - See Figure 1, month 0, blue shaded bars (2012-2015) with median daily dose 10 mg).
- There is also precedence for empiric initial dosing (non-weight based) in adult kidney transplant recipients.
  - For example, Meziyerh S et al[94] published their experience between 2009-2018, with typical initial tacrolimus dosing of 5 mg twice daily (all recipients) followed by TDM.
  - Stumpf J et al[95] recently trialled a lower dose size as initial empiric dosing, reporting noninferiority in a trial of 432 adult kidney transplant recipients on quadruple therapy (IL2Ra/Pred/Tac/MMF). Low empiric dosing of 5 mg daily Advagraf for the 1st post-transplant week (~half the typical starting dose immediate release tacrolimus), was compared with usual weight-based followed by TDM tacrolimus dosing.
  - From ANZDATA, adult kidney transplant recipients median = 5 mg twice daily. All kidney transplant recipients (allometric scaling to 70kg), 5 mg/dose twice daily during first month after transplant (Metz PhD thesis[93], see Figure 1).

*Figure 1: Daily dose TAC in kidney transplant recipients (adults & children), scaled to 70 kg adult. By year, for each post-transplant visit. Data as boxplots with IQR, median (black circle) and average (open circle).[93]*

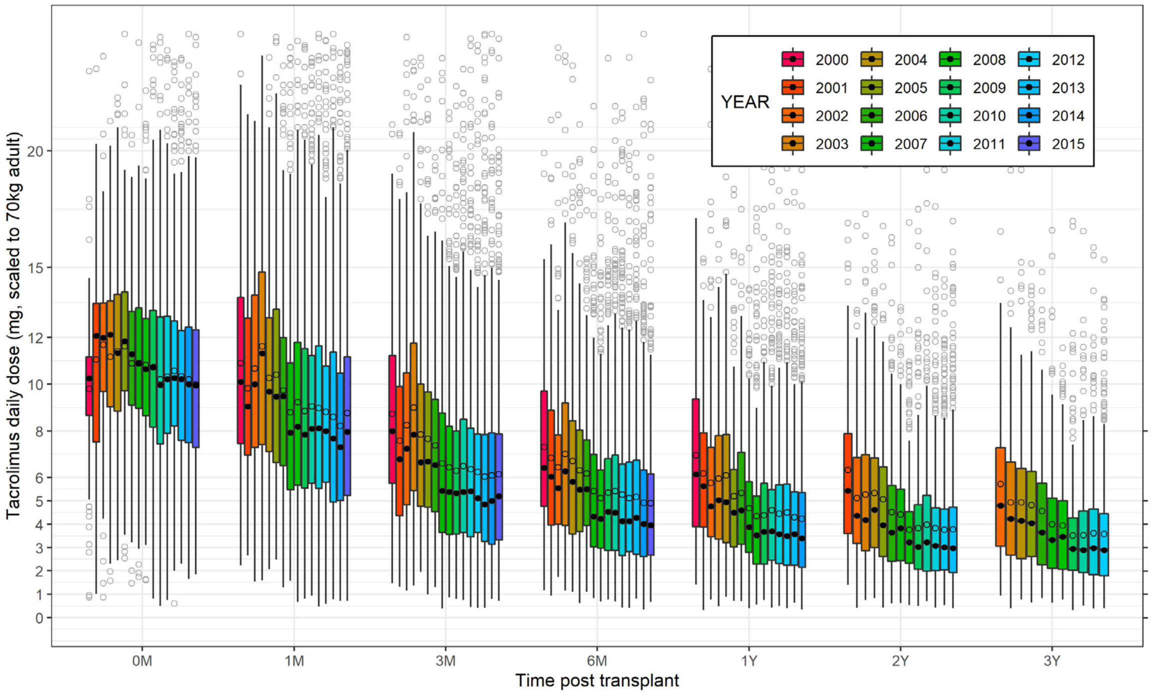

- **Allometric size scaling for tacrolimus dose**

Allometric scaling more accurately describes the relationship between size and drug clearance compared to e.g. linear functions (mg/kg), as well as having robust theoretic underpinnings.[96,97] For tacrolimus, fat free mass is the best size descriptor used with allometric scaling.[57]

The resultant equivalent mg/kg dose achieved with such an approach increases with decreasing weight. This more precisely describes the need for a greater per-kilogram dose in younger children, as has been found empirically by others who have recommended age or weight-based thresholds for dosing younger children. See Figure 3.

- Kausman et al 2008[92]
  - <40 kg–0.15 mg/kg dose twice daily
  - >40 kg–0.1 mg/kg/ dose twice daily (maximum 10 mg/dose).
- Min et al 2018[10]
  - 0.075 mg/kg twice daily for >6yo
  - 0.1 mg/kg twice daily for <6yo
  - 2x dose for expressers
  - Cap dose at 5 mg twice daily.

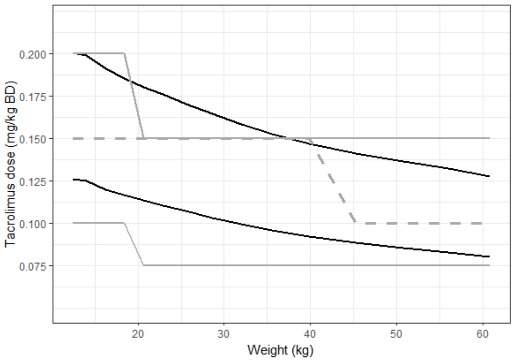

*Figure 2: Equivalent mg/kg dose by weight, allometric scaling of tacrolimus dose (black lines, upper line for CYP3A5 expressers). Grey line per-kilogram dose in Min 2018[10], grey-dashed as proposed by Kausman 2008[98].*

***Table 2 Size descriptor for allometric scaling of tacrolimus***

| Source | Covariate | |
|---|---|---|
| **Janmahasatian S et al 2005[99]**<br>**Anderson BJ, Holford NHG 2009[100]**<br>**Anderson BJ, Holford NHG 2017[97]** | Size descriptor for tacrolimus | Allometric size = $(NFM/NFM_{STD})$^(3/4), where NFM = normal fat mass.<br><br>Fat free mass = (WHSMAX*(HTM^2) *WTKG / (WHS50*HTM^2+WTKG))<br><br>For tacrolimus:<br>$DoseRate_{Child} = DoseRate_{Adult}$ x (FFM / 56.1kg) ^ (3/4) |

- **Genotype assignment and influence**

The quantitative impact on 1st dose is derived from CPIC and literature.
<u>Proportional adjustment based on genotype is:</u>

- Non-expresser: No change
- CYP3A4*22 expresser: Dose x 0.74
- CYP3A5*1 expresser (hetero- or homozygous): Dose x 1.59
- Both CYP3A5*1 & CYP3A4*22: Dose x 1.18

*Table 3: Impact of CYP3A5*1 and CYP3A4*22*

| Study | Cohort & design | Summary | Implementation |
|---|---|---|---|
| **Storset et al[57]** | PPK model | CYP3A5 expressers:<br>• 30% increase of CL &<br>• 18% decrease in oral bioavailability.<br><br>Thus require 1.59x dose rate for equivalent exposure. | Superiority, validation set |
| **Francke et al 2021[12]** | Adult KTx<br>Single arm prospective trial.<br>N=60. | Intervention: algorithm using age, BSA, CYP3A4 and CYP3A5 genotypes, noting:<br>• Dose x1.62 if CYP3A5 expresser (*1/x or *1/*1)<br>• Dose x0.814 if hetero CYP3A4*22<br><br>Tac C0 of 10 mcg/L ~ AUC0-12h of 222 mcg/L.h in this cohort. | Validation cohort (n=304) with rich PK profile n=7 else C0 only.<br><br>Subsequent prospective trial. |

| | | | |
|---|---|---|---|
| **Andrews et al 2018**[101] | | 1stDose = <br>$=TargetConc*CL_{Tac}*(1.62^{CYP3A51exp})*(0.814^{CYP3A422exp})*((Age/56)\hat{}(-0.5))*((BSA/1.93)\hat{}(0.72))/1000$<br><br>Algorithm derived from PPK model, n=337, obs = 4527.<br>Rotterdam cohort (n=237), C0 only.<br>Leiden cohort (n=100), PK profile at median 2wk: C0, 1, 2, 3, 4, 5, 6h. | Day 3 post-tx<br>-34/59 (58%) within range)<br>-7% <5.0 mcg/L<br>-3% > 20 mcg/L |
| **Pallet et al 2015**[102] | Adult KTx prospective cohort, n=272. Retrospective analysis PK data (trough). | For patients heterozygous for CYP3A4*22<br>• Dose x0.67 [0.54; 0.84] cf CYP3A4*1/*1 carriers. | |
| **Lloberas et al 2024**[46]<br>**Lloberas et al 2017**<br>**Andreu et al 2017**[103]<br>**Andreu et al 2015**[104] | | <u>Genotype clusters:</u><br>Poor metaboliser (PM)<br>• CYP3A4*22 carrier & CYP3A5*3/*3<br>Intermediate metaboliser (IM)<br>• CYP3A4*1/*1 & CYP3A5 *3/*3 or<br>• CYP3A4*22 carrier & CYP3A5*1 carrier<br>Extensive metaboliser<br>• CYP3A4*1/*1 & CYP3A5*1 carrier.<br><u>Lloberas RCT (target trough 8)</u><br>CYP3A5*1, non-CYP3A4*22 = high metaboliser<br>• 0.057 mg/kg BD<br>CYP3A5*1, CYP3A4*22 = intermediate metaboliser<br>• 0.0428 mg/kg BD<br>CYP3A5*3/*3, CYP3A4*22 = poor metaboliser<br>• 0.028 mg/kg BD | Prospective trial, improved exposure attainment |
| **Elens et al 2017**[105]<br><br>**Elens et al 2013**[106]<br><br>**Elens et al 2011**[107] | N=96, with PK profile n=59.<br><br>N=185. Dose for target C0 33% lower with CYP3A4*22 | <u>Standard = 0.1 mg/kg BD</u><br><u>Proposed update:</u><br>CYP3A5*3 & CYP3A4*22 (allele freq: Cau 5%, Afr 0%)<br>• 0.07 mg/kg BD (0.7x of SOC)<br>CYP3A5*3/*3 & 3A4*1/*1 (af: Cau 84%, Afr 4%)<br>• 0.1-0.125 mg/kg BD (??)<br>CYP3A5*1 & CYP3A4*1/*1 (af: Cau 11%, Afr 96%)<br>• 0.15-0.175 mg/kg BD (1.5-1.75x SOC) | |
| **De Jonge et al 2013**[108] | | CYP3A4, CYP3A5, haematocrit | |

- **Prospective trials with genotype-informed 1st dose – recent literature**

There are several notable recent publications with genotype informed 1st dose. Francke et al 2021[12], in a single arm prospective trial in adult kidney transplant recipients, achieved 58% of recipients within therapeutic range on day 3 post kidney transplantation, using an algorithm containing both CYP3A5 and CYP3A4 status (expresser status leading to x1.62 of dose and 0.814 of dose respectively). Notably, they did have 2 recipients with tacrolimus C0 over 20 mcg/L.h. Their algorithm was from a popPK model published in 2018 by Andrews et al.[101]

Lloberas et al 2024[46] also improved early exposure using CYP3A5 and CYP3A4 genotype, though those expressing CYP3A5*1/*1 and poor metaboliser CYP3A4*22 led to similar dose as SOC in effect cancelling each other out.

Min et al 2018[10] also successfully improved early target attainment, in a paediatric cohort, using an age threshold for differing linear per-kilogram dosing, which is consistent in direction with allometric scaling, and doubling of dose for CYP3A5 expressers.

*Table 4 Genotype Effect Data*

| Study | Cohort & design | Summary | Implementation |
|---|---|---|---|
| **Francke et al 2021[12]**<br><br>**Andrews et al 2018[101]** | Adult KTx<br>Single arm prospective trial.<br>N=60. | Intervention: algorithm using age, BSA, CYP3A4 and CYP3A5 genotypes, noting:<br>• **Dose x1.62 if CYP3A5 expresser** (*1/x or *1/*1)<br>• **Dose x0.814 if hetero CYP3A4*22**<br><br>Tac C0 of 10 mcg/L ~ AUC0-12h of 222 mcg/L.h in this cohort.<br>1stDose = TargetConc × $CL_{Tac}$ × ($1.62^{CYP3A51exp}$) × ($0.814^{CYP3A422exp}$) × ((Age/56)^(-0.5)) × ((BSA/1.93)^(0.72))/1000 | **Validation cohort** (n=304) with rich PK profile n=7 else C0 only.<br><br>Subsequent prospective trial.<br>Day 3 post-tx<br>-34/59 (58%) within range)<br>-7% <5.0 mcg/L<br>**-3% > 20 mcg/L** |
| **Min et al 2018[10]** | Paediatric RCT | Tac dose (capped at maximum 5 mg/dose twice daily).<br>Age >6yo:<br>• CYP3A5 expresser: 0.15 mg/kg/dose twice daily<br>• CYP3A5 nonexpresser: 0.075 mg/kg/dose twice daily<br>Age ≤6yo<br>• CYP3A5 expresser: 0.2 mg/kg/dose twice daily<br>• nonexpresser: 0.1 mg/kg/dose twice daily<br>SOC: 0.1 mg/kg/dose twice daily | Prospective trial, improved exposure attainment on D4 |
| **Lloberas et al 2024[46]**<br>**Lloberas et al 2017**<br>**Andreu et al 2017[103]**<br>**Andreu et al 2015[104]** | | <u>Lloberas RCT (target trough 8)</u><br>CYP3A5*1, non-CYP3A4*22 = high metaboliser<br>• 0.057 mg/kg BD<br>CYP3A5*1, CYP3A4*22 = intermediate metaboliser<br>• 0.0428 mg/kg BD<br>CYP3A5*3/*3, CYP3A4*22 = poor metaboliser<br>• 0.028 mg/kg BD | Prospective trial, improved exposure attainment on D4 |

#### APPENDIX 3: Rationale for tacrolimus concentration target including haematocrit standardization

- **Haematocrit standardization of TAC concentration**

NextDose standardizes haematocrit concentrations to 45%, which corrects for the random between-occasion variability due to changes in haematocrit (see below). The tacrolimus target concentration (standardized to haematocrit of 45%) is calculated from the accepted target for time post-transplant, then corrected to 45% from the typical haematocrit for time post-transplant (e.g. in kidney transplant recipients, target concentration adjusted from the average haematocrit of 33% in the initial month) (48).

Tacrolimus concentration is measured in whole blood. A substantial proportion of tacrolimus in whole blood is partitioned into erythrocytes, though only unbound drug in plasma represents the "effective" concentration. Thus, the red cell mass (reflected in whole blood haematocrit) represents an important source of predictable variability in whole blood tacrolimus concentration associated with the unmeasured unbound concentration.

Therapeutic target concentrations have typically been derived from whole blood tacrolimus concentrations unstandardized for haematocrit. The haematocrit seen in the early post-kidney transplant period (i.e. 33% [57]) may be used to standardise literature value to a haematocrit of 45%.

This source of between-occasion variability also applies to an individual's haematocrit, and can be accounted for, and used within a Bayesian estimation platform, by standardizing WB concentration target using the measured haematocrit.

The use of tacrolimus standardised concentrations has been shown to improve target achievement [109].

*Table 5 Use of Haematocrit*

| Source | Covariate | |
|---|---|---|
| **Storset, Holford et al 2014[57], Staatz et al. 2015[49,57,110]** | Haematocrit | Adult kidney transplant cohort<br>Important predictable BOV<br>- Haematocrit |
| **Schijvens et al 2019[111]** | Haematocrit | Paediatric kidney transplant cohort |
| **Limsrichamrern et al 2016 [112]** | Haematocrit | Liver transplant cohort |
| **Sikma et al 2020[113]** | Haematocrit | Cardiac & thoracic transplant cohorts |

See also below online calculator for further reference:
[https://kidney.wiki/tacro-calculator/](https://kidney.wiki/tacro-calculator/)

- **Evidence for tacrolimus target concentration in absence of haematocrit standardisation**

Evidence for tacrolimus target concentration in contemporary regimen (IL2/Pred/Tac/MPA) is based on observational data rather than randomized concentration-controlled trial (RCCT).

When first introduced in the mid-90s, an open-label, randomized concentration-control trial was performed in KTRs alongside anti-lymphocyte globulin, azathioprine and prednisolone, with optimal safety and effectiveness within the range of 5-15 mcg/L $HCT_{NONSTD}$.[114-116]. Later, an RCCT alongside contemporary immunosuppression, the 2017 Elite-Symphony trial, cemented the superiority of tacrolimus over cyclosporine.[43] However, Symphony had just the one tacrolimus arm, alongside two cyclosporine concentration target arms, and thus whilst showing superiority of the agent did not help in defining the optimal target concentration. Furthermore, planned tacrolimus concentration range (3-7 mcg/L $HCT_{NONSTD}$) was not achieved, concentrations achieved were higher: median trough concentration was just above 7 ng/ml $HCT_{NONSTD}$ for the first 2 months, then gradually reducing to a mean of 6.4 ng/ml by 12-months (with interquartile range of trough concentrations always above 5 ng/ml $HCT_{NONSTD}$) [43].

Observational data has helped affirm and narrow the therapeutic range from early RCCT. Efforts to minimise tacrolimus exposure have been driven be association with various serious toxicities, including a 2005 meta-regression reporting less new onset diabetes after transplant with trough concentrations below 10 mcg/L $HCT_{NONSTD}$[117] and from Lichtenberg et al an association between time-weighted average tacrolimus trough concentration above 11 ng/mL at 6 or 12 months and an increased cancer risk.[19] Regarding low concentrations and rejection, a 2013 pooled analysis of 3 RCTs did not show relationship between C0 concentration and acute rejection early post-transplant.[118] This may, however, relate to noise and smaller signal in current regimen, more recent studies showing relationship between haematocrit corrected tacrolimus concentration and acute rejection in the first 2 weeks,[119] along with other dedicated studies.[9] In addition, over the past decade, a number of studies have shown relationship between time below trough concentrations of 7-8 mcg/L in the early months and inefficacy, including acute rejection, chronic rejection and de novo donor specific antibody formation.[15,18,37-40]

Broadly accepted therapeutic goal early post kidney transplant is to maintain C0 concentrations within 8-12 mcg/L $HCT_{NONSTD}$ initially, a recent review suggesting 7–12 ng/mL $HCT_{NONSTD}$ for the first 3 months then 5-8 mcg/L for month 3-12, then 5-7 thereafter.[120]

Whether rapid attainment of therapeutic target (e.g. 10 mcg/L $HCT_{NONSTD}$ trough) is required whilst covered with monoclonal IL2Ra and pulse methylprednisolone, as opposed to e.g. by day 5-7, has not been determined. Notably some adult transplant units start all recipients on an e.g. 5 mg twice daily dose,[94] and elsewhere non-inferiority shown in an RCT (n=432) comparing SOC with an initial lower fixed dose (advagraf 5 mg daily) followed by TDM.[94] Nevertheless, as above, there is clear advantage to time within therapeutic range over the initial weeks.

- **Determining Cavgss target from AUC and trough (C0)**

The proposed concentration target for Css avg is obtained by dividing $AUC_{ssDI}$ by the dosing interval e.g. 12 h for twice daily dosing. Css avg has the advantage over $AUC_{ssDI}$ in that it can be used with any dosing interval. An AUCssDI of 175 mcg/L.h HCT 33 corresponds to a $C_0$ of 10 mcg/L HCT 33 in patients who are CYP3A4 normal metabolisers and CYP3A5 non-expressers.

To determine Cavgss target that is consistent with accepted literature $C_0$ targets of in typical PK individuals, a targeted review extracted studies using an AUC target, large datasets correlating the typical trough concentration with AUC, and expert opinion, are summarised below. In addition, we extracted datasets correlating typical AUC and trough and calculated a weighted average for a more precise estimate of the correlation (see Table 6, below). For calculations, all TAC concentrations and AUC are interpreted as being at HCT 33 in the first month. This assumption is based on the typical HCT in the first 3 months after transplant.

*Table 6: Literature values and weighted average Cavgss target*

| Source | N (profiles) | $C_0$ (raw) | $C_0$ (HCT45) | $AUC_{0-12}$ (central) | | $AUC_{0-12}$ at $C_0$ (raw) | Cavgss (HCT45) |
|---|---|---|---|---|---|---|---|
| **Saint-Marcoux 2013** | 113 | 7.5 | 10.2 | 152.0 | | 202.7 | 23.0 |
| **Saint-Marcoux 2013** | 95 | 12.5 | 17.0 | 218.0 | | 174.4 | 19.8 |
| **Scholten 2005** | 33 | 11.9 | 16.2 | 181.0 | | 152.1 | 17.3 |
| **Braun 2001** | 20 | 9.3 | 12.7 | 159.3 | | 171.3 | 19.5 |
| **Kuypers 2004** | 100 | 10.3 | 14.0 | 168.5 | | 163.6 | 18.6 |
| **Kuypers 2004** | 100 | 9.0 | 12.3 | 154.7 | | 171.9 | 19.5 |
| **Wlodarczyk 2009** | 32 | 10.1 | 13.8 | 180.7 | | 178.6 | 20.3 |
| **Wlodarczyk 2009** | 32 | 10.0 | 13.7 | 171.8 | | 171.5 | 19.5 |
| **Wlodarczyk 2009** | 32 | 12.1 | 16.4 | 191.3 | | 158.6 | 18.0 |
| **Niokka 2012** | 47 | 9.3 | 12.7 | 164.8 | | 177.2 | 20.1 |
| **Sum or weighted average** | **604** | **10.2** | **13.9** | **174.2** | **$AVG_{WGTD}$ (raw)** | **175.6** | |
| **Weighted average, HCT45** | | | | | **$AVG_{WGTD}$ (HCT45)** | **239.5** | **19.96** |

The average CssAVG in Table 3 is 19.96 mcg/L HCT45. This is based on variety of sources with different AUCssDI and C0 ranges, with a high level of consistency, as well as consistency with consensus and precedence (see below). A target CssAvg of 20 mcg/L HCT45 is thus proposed.

The above data-driven estimates are also consistent with precedence and published expert opinion:

1. Expert opinion
    a. A 2009 consensus paper[121] (update 2019[36]) on tacrolimus personalized therapy suggested an $AUC_{0\text{-}12h}$ TDM "therapeutic window" of 150-200 mcg/L (thus mid-point 175 mcg/L.h) in the early post-transplant period
2. Correlation between $C_0$ and $AUC_{0\text{-}12h}$.
    a. The largest dataset to determine correlation between $C_0$ and $AUC_{0\text{-}12h}$ was by Saint-Marcoux et al,[83] using data to their online Bayesian dosing platform (ISBA).
        i. They analysed 2030 tacrolimus PK profiles from 1000 kidney transplant recipients, and among other analyses used regression analysis to determine the relationship between $C_0$ and $AUC_{0\text{-}12h}$.
        ii. From this they determined that, in the early post-transplant period (first 3 months),
            1. a $C_0$ range of 8-12 mcg/L with 140-210 mcg/L (average 175 mcg/L)
    b. In a paediatric analysis from the same group, with 1935 PK profiles from 419 paediatric recipients, they determined that in the first 3 months,
        i. a $C_0$ range of 8-12 mcg/L correlated with an $AUC_{0\text{-}12h}$ of 170-240 mcg/L.h (mid-averaget 205 mcg/L.h).[82]
    c. An earlier group (based on 20 paediatric transplant recipients) suggested
        i. A tacrolimus $C_0$ range of 10-15 mcg/L.h was reported as equivalent to an $AUC_{0\text{-}12h}$ of 175-250 mcg/L in children.[122]
3. Practice
    a. In a 2005 study adult kidney transplant study, tacrolimus was dosed to an AUC0-12h target, alongside prednisolone, mycophenolate and IL2Ra blockade.[79] Target concentration was
        i. 210 mcg/L.h for the first 6 wks post-transplant ("corresponding to a trough of 12.5 mcg/L") and then

ii. 125 mcg/L.h thereafter ("corresponding to a C0 of 7.5 mcg/L.h").

b. Most recently, Meziyerh et al[94] reported on their experience with AUC-guided dosing of MPA and tacrolimus. Tacrolimus was commenced at an empiric dose of 5 mg twice daily, with subsequent titration to
   i. an AUC0-12h of 160-180 mcg/L.h (mid-point 170 mcg/L.h) for the first 6-weeks, which they suggested equivalent to a Tac C0 of 8-12 mcg/L.h.

Expanding this to the second and third post-transplant month yields the following:

***Table 7: Conversion from protocol C0 target to Cavgss.***

| Kidney Tx | HCT non-standardized | | | | HCT45 | | | |
|---|---|---|---|---|---|---|---|---|
| Post KTx month | C0 low | C0 target | C0 high | Equiv. $AUC_{0-12h}$[a] | Equiv. Cavgss[b] | Trough target[c] | $AUC_{0-12h}$ target | Cavgss target[c] |
| M1 | 8 | 10 | 12 | 175 | 14.6 | 13.6 | 240 | 20 |
| M2 | 6 | 8 | 10 | 140 | 11.7 | 10.9 | 180 | 15 |
| M3 | 6 | 7 | 8 | 122.5 | 10.2 | 9 | 158 | 13.125 |

[a]See protocol for determination AUC from C0
[b]Equal to AUC0-12h/12
[c]HCTnonstd multiplied by 45/33 (month 1) or 45/35 (month 2)

#### APPENDIX 4: Standard Operating Procedure for using NextDose

##### Standard Operating Procedure: NextDose for BRUNO-PIC

##### Summary

1. **Enter all patient details,** including genotype, dosing record and observation record.
2. **Perform dose estimation**
   a) **Model**:
      - Choose **PK Storset 2024 AVG** or **PK Storset 2024 BTEL AVG**
         - If using concentrations from initial 5-7 days, use **BTEL AVG**

   b) **PK concentration metric**
      - If CYP3A5 expresser, phenotypic rapid metaboliser ($C_0$/Dose < 1.05), [53] or by clinician choice, choose **Cssavg**. This is the preferred PK metric, used in all kidney transplant recipients.
      - For a target trough concentration ($C_0$), choose **Csstrough**.

   c) **Target concentration** (see full BRUNOPIC protocol for determination of targets)
      - For kidney transplant recipients

| Post KTx month | Csstrough target (HCT45) | Cavgss target (HCT45) |
|---|---|---|
| M1 | 13.6 | 20 |
| M2 | 10.9 | 15 |

3. **NextDose Output**
   a) Dose recommendation is given in bold green, average of Bayesian estimates
      - Proposed PO maintenance dose XXX mg every 12.0 hours (Average)

   b) Accuracy of *input* can be reviewed by:
      - inspecting concentration-time course
      - separate inspection of dosing record and observation record (including time post dose) on the results PRINT page.

   c) This results from an estimation can be printed e.g. in pdf format as a full record of the information used to propose the dose to achieve the target whole blood concentration.

##### Background

NextDose is an online Bayesian dosing platform that sits on a secure server currently hosted by the University of Auckland.

Bayesian dosing software ***estimates an individual's drug pharmacokinetic (PK) parameters*** by combining knowledge about a drugs PK behavior with information from the individual: ***clinical characteristics*** *(e.g. weight, age, renal function, genotype) and* ***measured drug concentrations***.

- A population PK model is a mathematical-statistical model developed in a real patient population which describes the drug's typical pharmacokinetics, how this varies between individuals and over time, and factors that influence this variability.

- By leveraging prior knowledge about a drug's pharmacokinetics with individual patient characteristics and drug concentrations, an accurate estimate of an individual's PK characteristics can be determined. Furthermore, additional drug concentrations over time (subsequent days) iteratively improve accuracy of individual PK estimates.

***NextDose*** is a ***clinical decision support software***. The ***NextDose*** output gives ***a dose recommendation***, as well as providing a ***visual output of the predicted concentration-time curve*** and how this relates to the observed concentrations. This allows clinical review of recommendation and decision on whether to implement.

#### NextDose online manual

The basic online manual (below) walks through the process of creating a new patient record, then entering their data and finally making a dose estimation:
https://www.nextdose.org/manual
In addition, specific detail on Bayesian dosing of tacrolimus using NextDose is detailed here:
https://wfn.sourceforge.net/clinpharmacol/nextdose/medicines/tacrolimus.htm
Further information on Bayesian dosing and NextDose are listed here:
https://wfn.sourceforge.net/clinpharmacol/nextdose/index.htm

Specific information for use of NextDose for BRUNO-PIC follows.

#### NextDose for tacrolimus dose recommendation in BRUNO-PIC

#### NextDose is an online platform:

**https://www.nextdose.org/**

BRUNOPIC participants are entered under the same username:

#### Creating a new participant and entering data

1. A new participant is created and saved within NextDose
   a) Click New Patient tab
   b) In the **Patient Details** tab, fill in NextDose ID, age and sex, genotype.
   c) The relevant genotypes for tacrolimus are CYP3A4 & CYP3A5 (see Figure 1)
      - CYP3A4: Normal metaboliser *1/*1
      - CYP3A4: Poor metaboliser *22
      - CYP3A5: Nonexpresser *3/*3
      - CYP3A5: Expresser *1/*1 or *1/*3
   d) Save changes.

*Figure 3: Genotype Selection*

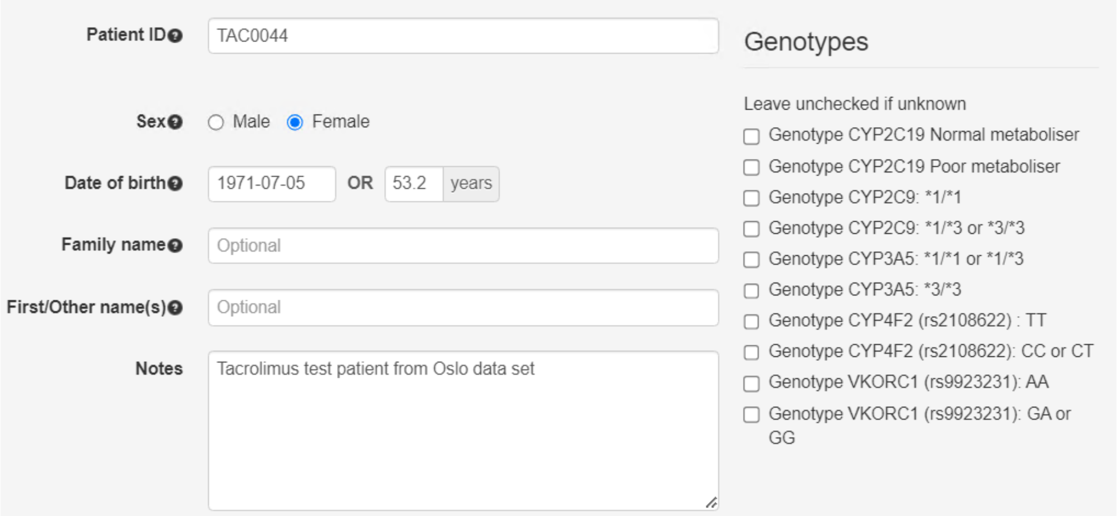

2. Click **Add a new medicine** and choose **Tacrolimus**.
3. In the **Doses & Observations** tab, the following details will need entering (further details in point 4)
   **a) The observations (clinical characteristics & measurements for the individual)**
      i. The important covariates for tacrolimus Bayesian estimation are:
         - **Genotype**: entered once, on the patient details page
         - **Date of Transplant**: entered once
         - **Height** (cm) & **Weight** (kg): enter pre-transplant measurement, then update if a change has occurred.
         - **Prednisolone daily dose** (mg/day). Update this every time the dose changes.
         - **Hematocrit (%)**: crucial source of between-occasion variability, warrants updating with each hematocrit measurement.
         - **Serum creatinine** (umol/L). Update this every time Scr is measured. Note that Scr has no influence on tacrolimus dose prediction but it is important for assessing the change in renal function during treatment.

      ii. Each observation entry requires a **time** & **date**. For some this will be known, others should be guessed at by the user to provide a plausible value.
      iii. It is important to ensure that the **observations are updated** (with the most appropriate value) **prior to the next drug concentration** measurement.

   **b) The tacrolimus drug concentrations**
      i. Enter all measured tacrolimus concentrations, with date & time
      ii. Ensure appropriate covariates, especially hematocrit, updated at or prior to tacrolimus concentration date & time (see above).
         - PK parameter estimation is performed each time there are measured drug concentrations within a dosing interval. Thus, all relevant **observations are updated in NextDose** where applicable (with the most appropriate value) **prior to the next drug concentration** measurement, unless the prior observation remains relevant.
         - The timing of hematocrit relative to concentration measurement needs to be carefully considered. **For example, if there is no haematocrit measurement** at the time of a tacrolimus concentration measurement, however, it was measured 1h later with a value of 34%, and the prior measurement was a month earlier at value of 27%, then **the value of 34% should be entered prior to the tacrolimus concentration timepoint**. Otherwise, the prior haematocrit (27%) will be carried forward for the estimation, biasing the output.

**c) The tacrolimus dosing record**

i. Enter each tacrolimus dose, including dose size (mg), and date & time of administration

ii. Note if the same dose is repeated multiple times at a constant interval

- Check the Repeat checkbox.
- Enter the dose interval and the number of doses.
- Alternatively, "at steady state" can be used where the patient can be assumed at steady state at the time date/time specified

**<u>Determining target concentration</u>**

4. Move to the **<u>Result</u>** tab, where will appear the heading **Dose Prediction Options**
5. Fill in details as follows

**a) Model**

- Choose **PK Storset 2024 AVG** or **PK Storset 2024 BTEL AVG**
- If using concentrations from initial 5-7 days, use **BTEL AVG**

b) **PK concentration metric**

- First determine the units (mcg/L) and PK metric.
  - If CYP3A5 expresser, phenotypic rapid metaboliser (C0/D < 1.05) or by clinician choice, choose **Css avg** (see Figure 2). **This is the preferred PK metric**, used in all kidney transplant recipients.
  - For a target trough concentration ($C_0$), choose Css trough (see Figure 3).
- Then enter the target.

c) **Target concentration** (see full BRUNOPIC protocol for determination of targets)

- For kidney transplant recipients

| Post KTx month | Csstrough target (HCT45) | Cavgss target (HCT45) |
|---|---|---|
| M1 | 13.6 | 20 |
| M2 | 10.9 | 15 |

*Figure 4 Target concentration is Css avg*

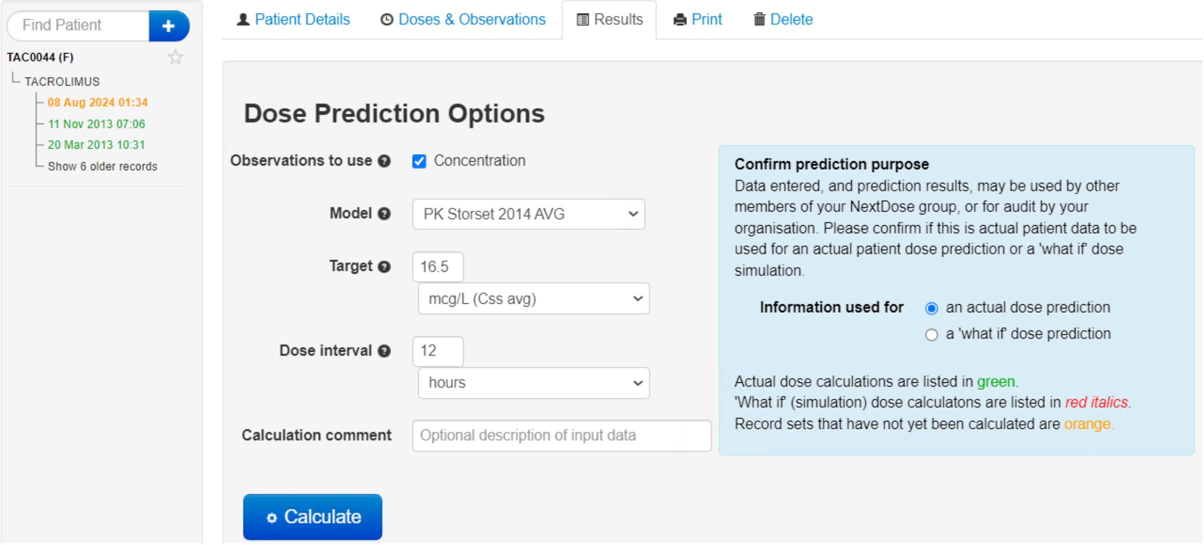

*Figure 5 Target concentration is Csstrough*

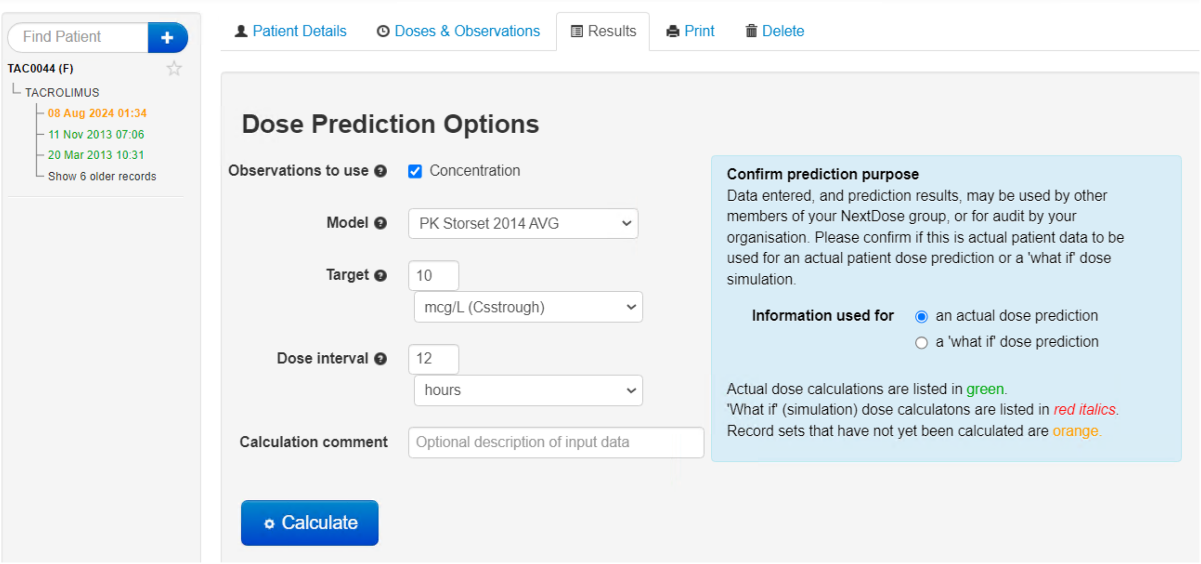

#### <u>Perform estimation to receive dose recommendation</u>

1. Hit **<u>Calculate</u>**. The estimation will occur, followed by detailed output including concentration-time course that can be visualized, and **dose recommendation given**.
2. This information be printed e.g. in pdf format as a full record of the information used to propose the dose to achieve the target whole blood concentration.
3. Correct *entry* of data can be checked by visual inspection of concentration-time curve, and in the printed report, checking separate dosing record, and time-post dose for concentration (and other) observations.

**APPENDIX 5: Expedited Safety Report Form**

| **EXPEDITED SAFETY REPORT FORM** | |
|---|---|
| Reporting requirement: All sites to report to Sponsor-Investigator all *SAEs, SUSARs and USMs within 24 hours of **trial** staff becoming aware of the event.<br>**Except those identified in the protocol as not needing immediate reporting* | |
| **HREC Reference #** | |
| **Project title** | BRUNO-PIC |
| **Section A: To be completed by the Local Site** | |
| **Site:** | |
| **Local Site Principal Investigator:** | |
| **Participant Enrolment OR Randomisation No.:** | |
| **Date the safety event occurred:** | |
| **Date Local Site Principal Investigator became aware of the safety event:** | |
| **Participant's date of birth, age and weight:** | |
| **Event description and management:** | |
| **Event outcome (synopsis):** | |
| **Trial phase** *(amend to reflect protocol)* | ☐ Screening<br>☐ Treatment<br>☐ Follow Up |
| **Relationship to the trial drug** | ☐ Unrelated<br>☐ Unlikely to be related<br>☐ Possibly related<br>☐ Probably related |
| **Expectedness (only complete for SAEs that are probably/possibly related):** | ☐ Not applicable<br>☐ Expected<br>☐ *Unexpected<br>*Report SUSAR to local RGO within 72 hours of becoming aware of event |

| | |
|---|---|
| **Was an Urgent Safety Measure (USM) instigated?** | * ☐ Yes ☐ No |

| | | |
|---|---|---|
| *A measure required to be taken in order to eliminate an immediate hazard to a participant's health or safety.* | *Report to local RGO within 72 hours of becoming aware of event | |
| Name and Signature (of local PI or delegate) | | Date |
| **Section B: To be completed by the Sponsor-Investigator only** | | |
| **Is this event a Significant Safety Issue (SSI)?**<br>*A safety issue that could adversely affect the safety of participants or materially impact on the continued ethical acceptability of the trial. Often SSIs do not fall within the definition of a Suspected Unexpected Serious Adverse Reaction (SUSAR), thus are not reported as SUSARs but require other action such as the reporting of an urgent safety measure (USM), an amendment, a temporary halt or early termination of a trial.* | | *☐ Yes ☐ No<br><br>* Report to TGA, HREC and all site PIs within 15 days of becoming aware of event |
| **Is this event an Urgent Safety Measure (USM)?**<br>*A measure required to be taken in order to eliminate an immediate hazard to a participant's health or safety.* | | * ☐ Yes ☐ No<br><br>*Report to TGA, HREC and all site PIs within 72 hours of becoming aware of event |
| **Is this event a SUSAR?** | | * ☐ Yes ☐ No<br><br>*Report to TGA within 7 days of becoming aware of the event if fatal/life threatening, otherwise report within 15 calendar days |
| **Does the protocol require amending as a result of this safety event?**<br>(If Yes, submit an amended protocol to approving HREC) | | ☐ Yes ☐ No |
| **Do the participant information statements require amending as a result of this safety event?**<br>(If Yes, submit a**n amendment** request to approving HREC and RGOs with the amended forms) | | ☐ Yes ☐ No |
| **Is a temporary halt or early termination of the trial required as a result of this safety event?**<br>(If Yes, ensure actions are taken within 15 days of decision to halt) | | ☐ Yes ☐ No |
| Name and Signature (of Sponsor-Investigator) | | Date |

#### APPENDIX 6: Departmental guidelines

- **RCH Renal Transplant Guidelines**

https://www.rch.org.au/uploadedFiles/Main/Content/nephrology/intranet-only/ktx-transplant-patient-medication-reference-guide.pdf

ktx-transplant-patient
-medication-reference

Tacrolimus concentrations are measured daily for the $1^{st}$ post-transplant month, thrice weekly for the $2^{nd}$ post-transplant month and twice weekly for the $3^{rd}$ post-transplant month.

- **RCH Liver Transplant Guidelines**

https://www.rch.org.au/uploadedFiles/Main/Content/gastro/intranet-only-security/LTxProtocol-s6.1-Medication_list-Feb23.pdf

LTxProtocol-s6.1-Me
dication_list-Feb23.pd

- **RCH Cardiac Transplant Guidelines**

https://www.rch.org.au/uploadedFiles/Main/Content/cardiology/intranet_resources/Heart_Transplant_Guidelines.pdf

Heart_Transplant_Gui
delines.pdf

#### APPENDIX 6: Common Medications Interactions with Tacrolimus

| Interaction: | Medications: | Management |
|---|---|---|
| Drugs that **increase** tacrolimus concentration | • Antimicrobials:<br> ○ Erythromycin, Clarithromycin, Azithromycin, Metronidazole, Chloramphenicol, Doxycycline, Clotrimazole, Fluconazole, Ketonconazole, Itraconazole, Voriconazole, Posaconazole<br>• Cardiovascular:<br> ○ Nifedipine, Diltiazem, Verapamil, Amiodarone, Thiazides<br>• Other:<br> ○ Methylprednisolone, Oral Contraceptives, Metoclopramide, Omeprazole<br>• Antidepressants:<br> ○ Fluoxetine, Fluvoxamine, Sertraline, Venlafaxine, Mirtazapine, Paroxetine | • Monitor concentrations closely following addition, dose changeor discontinuation.<br>• Can decrease effectiveness of oral contraceptive |
| Drugs that **decrease** Tacrolimus concentration | • Dexamethasone<br>• Anticonvulsants:<br> ○ Phenytoin, Phenobarbitone, Carbamazepine<br>• Antimicrobial:<br> ○ Rifampicin, Isoniazid, Rifabutin, Caspofungin, Trimethoprim-sulfamethoxazole | • Monitor concentrations closely following addition, dose changeor discontinuation.<br>• Can cause increased concentration of phenytoin, monitor closely |
| Drugs to use with caution because of **additive toxicity** | **Risk of Nephrotoxicity:**<br>• Vancomycin, Gentamicin, Tobramycin, Amphotericin, Amikacin, Ganciclovir, Cyclosporin, Sulfonamides, Indomethacin, NSAIDs (except low dose aspirin**)**, ACE inhibitors, trimethoprim<br>**Risk of Hyperkalaemia:**<br>• Spironolactone, Potassium Supplements, ACE inhibitors<br>**Risk of neurotoxicity:**<br>• Aciclovir, Ganciclovir, Ciprofloxacin, Imipenem | • Avoid these medications if possible.<br>• Monitor renal function closely.<br>• Monitor serum potassium levels |
| **Other interactions:** | • **Avoid (affect metabolism):**<br> ○ St John's wort and other herbal remedies<br> ○ Grapefruit juice<br>• **Use with caution:**<br> ○ Concurrent use of QT prolonging drugs |  |

Modified from Liver Transplant Guidelines. Ref: Lexicomp, Micromedex, AMH.

#### 17 REFERENCES

1. Riva N, Woillard JB, Distefano M, et al. Identification of Factors Affecting Tacrolimus Trough Levels in Latin American Pediatric Liver Transplant Patients. Liver Transpl 2019;25(9):1397-1407. (In eng). DOI: 10.1002/lt.25495.
2. Barbarino JM, Staatz CE, Venkataramanan R, Klein TE, Altman RB. PharmGKB summary: cyclosporine and tacrolimus pathways. Pharmacogenetics and genomics 2013;23(10):563.
3. Neuberger JM, Bechstein WO, Kuypers DR, et al. Practical recommendations for long-term management of modifiable risks in kidney and liver transplant recipients: a guidance report and clinical checklist by the consensus on managing modifiable risk in transplantation (COMMIT) group. Transplantation 2017;101(4S):S1-S56.
4. Yanik MV, Seifert ME, Locke JE, et al. CYP3A5 genotype affects time to therapeutic tacrolimus level in pediatric kidney transplant recipients. Pediatric transplantation 2019;23(5):e13494.
5. Cheng F, Li Q, Cui Z, Wang Z, Zeng F, Zhang Y. Tacrolimus Concentration Is Effectively Predicted Using Combined Clinical and Genetic Factors in the Perioperative Period of Kidney Transplantation and Associated with Acute Rejection. Journal of Immunology Research 2022.
6. Woillard J-B, Mourad M, Neely M, et al. Tacrolimus updated guidelines through popPK modeling: how to benefit more from CYP3A pre-emptive genotyping prior to kidney transplantation. Frontiers in pharmacology 2017;8:358.
7. Undre NA, van Hooff J, Christiaans M, et al. Low systemic exposure to tacrolimus correlates with acute rejection. Transplant Proc 1999;31(1-2):296-8. (In eng). DOI: 10.1016/s0041-1345(98)01633-9.
8. Hu R, Barratt DT, Coller JK, Sallustio BC, Somogyi AA. Is There a Temporal Relationship Between Trough Whole Blood Tacrolimus Concentration and Acute Rejection in the First 14 Days After Kidney Transplantation? Therapeutic Drug Monitoring 2019;41(4) (https://journals.lww.com/drug-monitoring/Fulltext/2019/08000/Is_There_a_Temporal_Relationship_Between_Trough.14.aspx).
9. Cheng F, Li Q, Cui Z, Wang Z, Zeng F, Zhang Y. Tacrolimus Concentration Is Effectively Predicted Using Combined Clinical and Genetic Factors in the Perioperative Period of Kidney Transplantation and Associated with Acute Rejection. J Immunol Res 2022;2022:3129389. (In eng). DOI: 10.1155/2022/3129389.
10. Min S, Papaz T, Lafreniere-Roula M, et al. A randomized clinical trial of age and genotype-guided tacrolimus dosing after pediatric solid organ transplantation. Pediatr Transplant 2018;22(7):e13285. (In eng). DOI: 10.1111/petr.13285.
11. Anutrakulchai S, Pongskul C, Kritmetapak K, Limwattananon C, Vannaprasaht S. Therapeutic concentration achievement and allograft survival comparing usage of conventional tacrolimus doses and CYP3A5 genotype-guided doses in renal transplantation patients. Br J Clin Pharmacol 2019;85(9):1964-1973. (In eng). DOI: 10.1111/bcp.13980.
12. Francke MI, Andrews LM, Le HL, et al. Avoiding Tacrolimus Underexposure and Overexposure with a Dosing Algorithm for Renal Transplant Recipients: A Single Arm Prospective Intervention Trial. Clin Pharmacol Ther 2021;110(1):169-178. (In eng). DOI: 10.1002/cpt.2163.
13. Yang H, Sun Y, Yu X, et al. Clinical Impact of the Adaptation of Initial Tacrolimus Dosing to the CYP3A5 Genotype After Kidney Transplantation: Systematic Review and Meta-Analysis of Randomized Controlled Trials. Clin Pharmacokinet 2021;60(7):877-885. (In eng). DOI: 10.1007/s40262-020-00955-2.
14. Raj TY, Fernando ME, Srinivasa Prasad ND, et al. Efficacy and Outcomes of CYP3A5 Genotype-Based Tacrolimus Dosing Compared to Conventional Body Weight-based Dosing in Living Donor Kidney Transplant Recipients. Indian J Nephrol 2022;32(3):240-246. DOI: 10.4103/ijn.IJN_278_20.
15. Israni AK, Riad SM, Leduc R, et al. Tacrolimus trough levels after month 3 as a predictor of acute rejection following kidney transplantation: a lesson learned from DeKAF Genomics. Transpl Int 2013;26(10):982-9. (In eng). DOI: 10.1111/tri.12155.
16. Leino AD, Park JM, Pasternak AL. Impact of CYP3A5 phenotype on tacrolimus time in therapeutic range and clinical outcomes in pediatric renal and heart transplant recipients. Pharmacotherapy 2021;41(8):649-657. (In eng). DOI: 10.1002/phar.2601.
17. Davis S, Gralla J, Klem P, Stites E, Wiseman A, Cooper JE. Tacrolimus Intrapatient Variability, Time in

Therapeutic Range, and Risk of De Novo Donor-Specific Antibodies. Transplantation 2020;104(4):881-887. (In eng). DOI: 10.1097/tp.0000000000002913.

18. Davis S, Gralla J, Klem P, et al. Lower tacrolimus exposure and time in therapeutic range increase the risk of de novo donor-specific antibodies in the first year of kidney transplantation. Am J Transplant 2018;18(4):907-915. (In eng). DOI: 10.1111/ajt.14504.
19. Lichtenberg S, Rahamimov R, Green H, et al. The incidence of post-transplant cancer among kidney transplant recipients is associated with the level of tacrolimus exposure during the first year after transplantation. European journal of clinical pharmacology 2017;73(7):819-826. (In eng). DOI: 10.1007/s00228-017-2234-2.
20. van Gelder T, Meziyerh S, Swen JJ, de Vries APJ, Moes D. The Clinical Impact of the C(0)/D Ratio and the CYP3A5 Genotype on Outcome in Tacrolimus Treated Kidney Transplant Recipients. Front Pharmacol 2020;11:1142. (In eng). DOI: 10.3389/fphar.2020.01142.
21. Jouve T, Fonrose X, Noble J, et al. The TOMATO Study (Tacrolimus Metabolization in Kidney Transplantation): Impact of the Concentration-Dose Ratio on Death-censored Graft Survival. Transplantation 2020;104(6):1263-1271. (In eng). DOI: 10.1097/tp.0000000000002920.
22. Schütte-Nütgen K, Thölking G, Steinke J, et al. Fast Tac Metabolizers at Risk ⁻ It is Time for a C/D Ratio Calculation. J Clin Med 2019;8(5) (In eng). DOI: 10.3390/jcm8050587.
23. Kluwe F, Michelet R, Mueller-Schoell A, et al. Perspectives on Model-Informed Precision Dosing in the Digital Health Era: Challenges, Opportunities, and Recommendations. Clinical Pharmacology & Therapeutics 2021;109(1):29-36. DOI: https://doi.org/10.1002/cpt.2049.
24. Størset E, Åsberg A, Skauby M, et al. Improved Tacrolimus Target Concentration Achievement Using Computerized Dosing in Renal Transplant Recipients--A Prospective, Randomized Study. Transplantation 2015;99(10):2158-66. (In eng). DOI: 10.1097/tp.0000000000000708.
25. Francke MI, Andrews LM, Le HL, et al. Avoiding Tacrolimus Underexposure and Overexposure with a Dosing Algorithm for Renal Transplant Recipients: A Single Arm Prospective Intervention Trial. Clin Pharmacol Ther 2021;110(1):169-178. (https://doi.org/10.1002/cpt.2163). DOI: https://doi.org/10.1002/cpt.2163.
26. Cooper JE. Evaluation and Treatment of Acute Rejection in Kidney Allografts. Clin J Am Soc Nephrol 2020;15(3):430-438. (In eng). DOI: 10.2215/cjn.11991019.
27. O'Connell PJ, Kuypers DR, Mannon RB, et al. Clinical Trials for Immunosuppression in Transplantation: The Case for Reform and Change in Direction. Transplantation 2017;101(7):1527-1534. DOI: 10.1097/TP.0000000000001648.
28. Neuberger JM, Bechstein WO, Kuypers DR, et al. Practical Recommendations for Long-term Management of Modifiable Risks in Kidney and Liver Transplant Recipients: A Guidance Report and Clinical Checklist by the Consensus on Managing Modifiable Risk in Transplantation (COMMIT) Group. Transplantation 2017;101(4S Suppl 2):S1-s56. (In eng). DOI: 10.1097/tp.0000000000001651.
29. Karuthu S, Blumberg EA. Common infections in kidney transplant recipients. Clinical journal of the American Society of Nephrology : CJASN 2012;7(12):2058-70. DOI: 10.2215/CJN.04410512.
30. Stoumpos S, Jardine AG, Mark PB. Cardiovascular morbidity and mortality after kidney transplantation. Transplant international : official journal of the European Society for Organ Transplantation 2015;28(1):10-21. DOI: 10.1111/tri.12413.
31. Bamgbola O. Metabolic consequences of modern immunosuppressive agents in solid organ transplantation. Therapeutic advances in endocrinology and metabolism 2016;7(3):110-27. (In eng). DOI: 10.1177/2042018816641580.
32. Miller LW. Cardiovascular toxicities of immunosuppressive agents. Am J Transplant 2002;2(9):807-18. (In eng). DOI: 10.1034/j.1600-6143.2002.20902.x.
33. Chapman JR, Webster AC, Wong G. Cancer in the transplant recipient. Cold Spring Harb Perspect Med 2013;3(7). DOI: 10.1101/cshperspect.a015677.
34. Au E, Wong G, Chapman JR. Cancer in kidney transplant recipients. Nature Reviews Nephrology 2018;14(8):508-520. DOI: 10.1038/s41581-018-0022-6.
35. Engels EA, Pfeiffer RM, Fraumeni JF, Jr., et al. Spectrum of cancer risk among US solid organ transplant recipients. Jama 2011;306(17):1891-901. (In eng). DOI: 10.1001/jama.2011.1592.

36. Brunet M, van Gelder T, Asberg A, et al. Therapeutic Drug Monitoring of Tacrolimus-Personalized Therapy: Second Consensus Report. Ther Drug Monit 2019;41(3):261-307. (In eng). DOI: 10.1097/ftd.0000000000000640.
37. Wiebe C, Rush DN, Nevins TE, et al. Class II Eplet Mismatch Modulates Tacrolimus Trough Levels Required to Prevent Donor-Specific Antibody Development. Journal of the American Society of Nephrology : JASN 2017;28(11):3353-3362. DOI: 10.1681/ASN.2017030287.
38. Beland MA, Lapointe I, Noel R, et al. Higher calcineurin inhibitor levels predict better kidney graft survival in patients with de novo donor-specific anti-HLA antibodies: a cohort study. Transplant international : official journal of the European Society for Organ Transplantation 2017;30(5):502-509. (In eng). DOI: 10.1111/tri.12934.
39. Wadström J, Ericzon B-G, Halloran PF, et al. Advancing Transplantation: New Questions, New Possibilities in Kidney and Liver Transplantation. Transplantation 2017;101(2):S1-S42. DOI: 10.1097/tp.0000000000001563.
40. Girerd S, Schikowski J, Girerd N, et al. Impact of reduced exposure to calcineurin inhibitors on the development of de novo DSA: a cohort of non-immunized first kidney graft recipients between 2007 and 2014. BMC Nephrol 2018;19(1):232-232. DOI: 10.1186/s12882-018-1014-2.
41. Calne RY, White DJ, Thiru S, et al. Cyclosporin A in patients receiving renal allografts from cadaver donors. Lancet 1978;2(8104-5):1323-7. (In eng). DOI: 10.1016/s0140-6736(78)91970-0.
42. Kahan BD. Individualization of cyclosporine therapy using pharmacokinetic and pharmacodynamic parameters. Transplantation 1985;40(5):457-76. (In eng). DOI: 10.1097/00007890-198511000-00001.
43. Ekberg H, Tedesco-Silva H, Demirbas A, et al. Reduced exposure to calcineurin inhibitors in renal transplantation. N Engl J Med 2007;357(25):2562-75. (In eng). DOI: 10.1056/NEJMoa067411.
44. Nankivell BJ, Borrows RJ, Fung CL, O'Connell PJ, Chapman JR, Allen RD. Calcineurin inhibitor nephrotoxicity: longitudinal assessment by protocol histology. Transplantation 2004;78(4):557-65. (In eng).
45. Matas AJ, Gaston RS. Moving Beyond Minimization Trials in Kidney Transplantation. Journal of the American Society of Nephrology : JASN 2015;26(12):2898-2901. (In eng). DOI: 10.1681/ASN.2015030245.
46. Lloberas N, Grinyo JM, Colom H, et al. A prospective controlled, randomized clinical trial of kidney transplant recipients developed personalized tacrolimus dosing using model-based Bayesian Prediction. Kidney international 2023. DOI: 10.1016/j.kint.2023.06.021.
47. Barbarino JM, Staatz CE, Venkataramanan R, Klein TE, Altman RB. PharmGKB summary: cyclosporine and tacrolimus pathways. Pharmacogenet Genomics 2013;23(10):563-85. (In eng). DOI: 10.1097/FPC.0b013e328364db84.
48. Venkataramanan R, Swaminathan A, Prasad T, et al. Clinical pharmacokinetics of tacrolimus. Clin Pharmacokinet 1995;29(6):404-30. (In eng). DOI: 10.2165/00003088-199529060-00003.
49. Størset E, Holford N, Midtvedt K, Bremer S, Bergan S, Åsberg A. Importance of hematocrit for a tacrolimus target concentration strategy. Eur J Clin Pharmacol 2014;70(1):65-77. (In eng). DOI: 10.1007/s00228-013-1584-7.
50. Staatz CE, Tett SE. Clinical pharmacokinetics and pharmacodynamics of tacrolimus in solid organ transplantation. Clin Pharmacokinet 2004;43(10):623-53. (In eng). DOI: 10.2165/00003088-200443100-00001.
51. Birdwell KA, Decker B, Barbarino JM, et al. Clinical Pharmacogenetics Implementation Consortium (CPIC) Guidelines for CYP3A5 Genotype and Tacrolimus Dosing. Clinical pharmacology and therapeutics 2015;98(1):19-24. (In eng). DOI: 10.1002/cpt.113.
52. Aranda JvdA, JN. Yaffe and Aranda's Neonatal and Pediatric Pharmacology: Therapeutic Principles in Practice. 5 ed. Lippincott Williams & Wilkins Wolters Kluwer, 2021.
53. van Gelder T, Meziyerh S, Swen JJ, de Vries APJ, Moes DJAR. The Clinical Impact of the C0/D Ratio and the CYP3A5 Genotype on Outcome in Tacrolimus Treated Kidney Transplant Recipients. Frontiers in Pharmacology 2020;11(1142) (Review) (In English). DOI: 10.3389/fphar.2020.01142.
54. Abdullah-Koolmees H, van Keulen AM, Nijenhuis M, Deneer VHM. Pharmacogenetics Guidelines: Overview and Comparison of the DPWG, CPIC, CPNDS, and RNPGx Guidelines. Front Pharmacol 2020;11:595219. (In eng). DOI: 10.3389/fphar.2020.595219.

55. Woillard JB, Mourad M, Neely M, et al. Tacrolimus Updated Guidelines through popPK Modeling: How to Benefit More from CYP3A Pre-emptive Genotyping Prior to Kidney Transplantation. Front Pharmacol 2017;8:358. DOI: 10.3389/fphar.2017.00358.
56. Dong Y, Xu Q, Li R, et al. CYP3A7, CYP3A4, and CYP3A5 genetic polymorphisms in recipients rather than donors influence tacrolimus concentrations in the early stages after liver transplantation. Gene 2022;809:146007. DOI: https://doi.org/10.1016/j.gene.2021.146007.
57. Storset E, Holford N, Hennig S, et al. Improved prediction of tacrolimus concentrations early after kidney transplantation using theory-based pharmacokinetic modelling. British journal of clinical pharmacology 2014;78(3):509-23. (In eng). DOI: 10.1111/bcp.12361.
58. Egeland EJ, Robertsen I, Hermann M, et al. High Tacrolimus Clearance Is a Risk Factor for Acute Rejection in the Early Phase After Renal Transplantation. Transplantation 2017;101(8):e273-e279. (In eng). DOI: 10.1097/tp.0000000000001796.
59. Friebus-Kardash J, Nela E, Möhlendick B, et al. Development of De Novo Donor-specific HLA Antibodies and AMR in Renal Transplant Patients Depends on CYP3A5 Genotype. Transplantation 2022;106(5):1031-1042. (In eng). DOI: 10.1097/tp.0000000000003871.
60. Ekberg H, Mamelok RD, Pearson TC, Vincenti F, Tedesco-Silva H, Daloze P. The challenge of achieving target drug concentrations in clinical trials: experience from the Symphony study. Transplantation 2009;87(9):1360-6. (In eng). DOI: 10.1097/TP.0b013e3181a23cb2.
61. Leino AD, Park JM, Pasternak AL. Impact of CYP3A5 phenotype on tacrolimus time in therapeutic range and clinical outcomes in pediatric renal and heart transplant recipients. Pharmacotherapy: The Journal of Human Pharmacology and Drug Therapy 2021;41(8):649-657. (https://doi.org/10.1002/phar.2601). DOI: https://doi.org/10.1002/phar.2601.
62. Woillard JB, Saint-Marcoux F, Debord J, Åsberg A. Pharmacokinetic models to assist the prescriber in choosing the best tacrolimus dose. Pharmacol Res 2018;130:316-321. (In eng). DOI: 10.1016/j.phrs.2018.02.016.
63. Holford N, Ma G, Metz D. TDM is dead. Long live TCI! British Journal of Clinical Pharmacology;n/a(n/a). DOI: 10.1111/bcp.14434.
64. Sanathanan LP, Peck CC. The randomized concentration-controlled trial: an evaluation of its sample size efficiency. Control Clin Trials 1991;12(6):780-94. (In eng). DOI: 10.1016/0197-2456(91)90041-j.
65. Mahalati K, Belitsky P, Sketris I, West K, Panek R. Neoral monitoring by simplified sparse sampling area under the concentration-time curve: its relationship to acute rejection and cyclosporine nephrotoxicity early after kidney transplantation. Transplantation 1999;68(1):55-62. (In eng). DOI: 10.1097/00007890-199907150-00011.
66. Undre NA. Pharmacokinetics of tacrolimus-based combination therapies. Nephrology, dialysis, transplantation : official publication of the European Dialysis and Transplant Association - European Renal Association 2003;18 Suppl 1:i12-5. (In eng). DOI: 10.1093/ndt/gfg1029.
67. van Rossum HH, Press RR, den Hartigh J, de Fijter JW. Point: A call for advanced pharmacokinetic and pharmacodynamic monitoring to guide calcineurin inhibitor dosing in renal transplant recipients. Clin Chem 2010;56(5):732-5. (In eng). DOI: 10.1373/clinchem.2009.141135.
68. Rojas L, Neumann I, Herrero MJ, et al. Effect of CYP3A5*3 on kidney transplant recipients treated with tacrolimus: a systematic review and meta-analysis of observational studies. The pharmacogenomics journal 2015;15(1):38-48. (In eng). DOI: 10.1038/tpj.2014.38.
69. Schutte-Nutgen K, Tholking G, Steinke J, et al. Fast Tac Metabolizers at Risk (-) It is Time for a C/D Ratio Calculation. Journal of clinical medicine 2019;8(5) (In eng). DOI: 10.3390/jcm8050587.
70. Trofe-Clark J, Brennan DC, West-Thielke P, et al. Results of ASERTAA, a Randomized Prospective Crossover Pharmacogenetic Study of Immediate-Release Versus Extended-Release Tacrolimus in African American Kidney Transplant Recipients. Am J Kidney Dis 2018;71(3):315-326. (In eng). DOI: 10.1053/j.ajkd.2017.07.018.
71. Tholking G, Siats L, Fortmann C, et al. Tacrolimus Concentration/Dose Ratio is Associated with Renal Function After Liver Transplantation. Annals of transplantation 2016;21:167-79. (In eng). DOI: 10.12659/aot.895898.

72. Tholking G, Fortmann C, Koch R, et al. The tacrolimus metabolism rate influences renal function after kidney transplantation. PloS one 2014;9(10):e111128. (In eng). DOI: 10.1371/journal.pone.0111128.
73. Egeland EJ, Reisaeter AV, Robertsen I, et al. High tacrolimus clearance - a risk factor for development of interstitial fibrosis and tubular atrophy in the transplanted kidney: a retrospective single-center cohort study. Transpl Int 2019;32(3):257-269. (In eng). DOI: 10.1111/tri.13356.
74. Tholking G, Schmidt C, Koch R, et al. Influence of tacrolimus metabolism rate on BKV infection after kidney transplantation. Scientific reports 2016;6:32273. (In eng). DOI: 10.1038/srep32273.
75. Thölking G, Tosun-Koç F, Jehn U, et al. Improved Kidney Allograft Function after Early Conversion of Fast IR-Tac Metabolizers to LCP-Tac. J Clin Med 2022;11(5) (In eng). DOI: 10.3390/jcm11051290.
76. Langone A, Steinberg SM, Gedaly R, et al. Switching STudy of Kidney TRansplant PAtients with Tremor to LCP-TacrO (STRATO): an open-label, multicenter, prospective phase 3b study. Clin Transplant 2015;29(9):796-805. (In eng). DOI: 10.1111/ctr.12581.
77. Taber DJ, Gebregziabher MG, Srinivas TR, Chavin KD, Baliga PK, Egede LE. African-American race modifies the influence of tacrolimus concentrations on acute rejection and toxicity in kidney transplant recipients. Pharmacotherapy 2015;35(6):569-77. (In eng). DOI: 10.1002/phar.1591.
78. Woillard JB, Monchaud C, Saint-Marcoux F, Labriffe M, Marquet P. Can the Area Under the Curve/Trough Level Ratio Be Used to Optimize Tacrolimus Individual Dose Adjustment? Transplantation 2023;107(1):e27-e35. (In eng). DOI: 10.1097/tp.0000000000004405.
79. Scholten EM, Cremers SC, Schoemaker RC, et al. AUC-guided dosing of tacrolimus prevents progressive systemic overexposure in renal transplant recipients. Kidney Int 2005;67(6):2440-7. (In eng). DOI: 10.1111/j.1523-1755.2005.00352.x.
80. Darwich AS, Polasek TM, Aronson JK, et al. Model-Informed Precision Dosing: Background, Requirements, Validation, Implementation, and Forward Trajectory of Individualizing Drug Therapy. Annual Review of Pharmacology and Toxicology 2021;61(1):225-245. DOI: 10.1146/annurev-pharmtox-033020-113257.
81. Neely M, Jelliffe R. Practical, individualized dosing: 21st century therapeutics and the clinical pharmacometrician. Journal of clinical pharmacology 2010;50(7):842-7. (In eng). DOI: 10.1177/0091270009356572.
82. Marquet P, Cros F, Micallef L, et al. Tacrolimus Bayesian Dose Adjustment in Pediatric Renal Transplant Recipients. Ther Drug Monit 2021;43(4):472-480. (In eng). DOI: 10.1097/ftd.0000000000000828.
83. Saint-Marcoux F, Woillard JB, Jurado C, Marquet P. Lessons from routine dose adjustment of tacrolimus in renal transplant patients based on global exposure. Ther Drug Monit 2013;35(3):322-7. (In eng). DOI: 10.1097/FTD.0b013e318285e779.
84. Woillard JB, Debord J, Monchaud C, Saint-Marcoux F, Marquet P. Population Pharmacokinetics and Bayesian Estimators for Refined Dose Adjustment of a New Tacrolimus Formulation in Kidney and Liver Transplant Patients. Clin Pharmacokinet 2017;56(12):1491-1498. (In eng). DOI: 10.1007/s40262-017-0533-5.
85. Zhao CY, Jiao Z, Mao JJ, Qiu XY. External evaluation of published population pharmacokinetic models of tacrolimus in adult renal transplant recipients. Br J Clin Pharmacol 2016;81(5):891-907. (In eng). DOI: 10.1111/bcp.12830.
86. Aarons L. Physiologically based pharmacokinetic modelling: a sound mechanistic basis is needed. Br J Clin Pharmacol 2005;60(6):581-3. (In eng). DOI: 10.1111/j.1365-2125.2005.02560.x.
87. Nanga TM, Doan TTP, Marquet P, Musuamba FT. Toward a robust tool for pharmacokinetic-based personalization of treatment with tacrolimus in solid organ transplantation: A model-based meta-analysis approach. Br J Clin Pharmacol 2019;85(12):2793-2823. (In eng). DOI: 10.1111/bcp.14110.
88. Itohara K, Yano I, Nakagawa S, et al. Extrapolation of physiologically based pharmacokinetic model for tacrolimus from renal to liver transplant patients. Drug Metab Pharmacokinet 2022;42:100423. (In eng). DOI: 10.1016/j.dmpk.2021.100423.
89. Sam WJ, Aw M, Quak SH, et al. Population pharmacokinetics of tacrolimus in Asian paediatric liver transplant patients. Br J Clin Pharmacol 2000;50(6):531-41. DOI: 10.1046/j.1365-2125.2000.00288.x.
90. Textor SC, Wiesner R, Wilson DJ, et al. Systemic and renal hemodynamic differences between FK506 and cyclosporine in liver transplant recipients. Transplantation 1993;55(6):1332-9. (In eng). DOI:

10.1097/00007890-199306000-00023.
91. Lloberas N, Grinyó JM, Colom H, et al. A prospective controlled, randomized clinical trial of kidney transplant recipients developed personalized tacrolimus dosing using model-based Bayesian Prediction. Kidney Int 2023;104(4):840-850. (In eng). DOI: 10.1016/j.kint.2023.06.021.
92. Kausman JY, Patel B, Marks SD. Standard dosing of tacrolimus leads to overexposure in pediatric renal transplantation recipients. Pediatric transplantation 2008;12(3):329-335. DOI: https://doi.org/10.1111/j.1399-3046.2007.00821.x.
93. Metz D. Optimising Immunosuppressant dosing in kidney transplantation: better outcomes through quantitative pharmacology. Medicine, Dentistry and Health Sciences: University of Melbourne; 2020.
94. Meziyerh S, van Gelder T, Kers J, et al. Tacrolimus and Mycophenolic Acid Exposure Are Associated with Biopsy-Proven Acute Rejection: A Study to Provide Evidence for Longer-Term Target Ranges. Clin Pharmacol Ther 2023;114(1):192-200. DOI: https://doi.org/10.1002/cpt.2915.
95. Stumpf J, Budde K, Witzke O, et al. Fixed low dose versus concentration-controlled initial tacrolimus dosing with reduced target levels in the course after kidney transplantation: results from a prospective randomized controlled non-inferiority trial (Slow & Low study). EClinicalMedicine 2024;67:102381. (In eng). DOI: 10.1016/j.eclinm.2023.102381.
96. Anderson BJ, Holford NH. Understanding dosing: children are small adults, neonates are immature children. Arch Dis Child 2013;98(9):737-44. (In eng). DOI: 10.1136/archdischild-2013-303720.
97. Holford NHG, Anderson BJ. Allometric size: The scientific theory and extension to normal fat mass. Eur J Pharm Sci 2017;109s:S59-s64. (In eng). DOI: 10.1016/j.ejps.2017.05.056.
98. Kausman JY, Patel B, Marks SD. Standard dosing of tacrolimus leads to overexposure in pediatric renal transplantation recipients. Pediatric transplantation 2008;12(3):329-35. (In eng). DOI: 10.1111/j.1399-3046.2007.00821.x.
99. Janmahasatian S, Duffull SB, Ash S, Ward LC, Byrne NM, Green B. Quantification of lean bodyweight. Clin Pharmacokinet 2005;44(10):1051-65. (In eng). DOI: 10.2165/00003088-200544100-00004.
100. Anderson BJ, Holford NH. Mechanistic basis of using body size and maturation to predict clearance in humans. Drug Metab Pharmacokinet 2009;24(1):25-36. (In eng). DOI: 10.2133/dmpk.24.25.
101. Andrews LM, Hesselink DA, van Schaik RHN, et al. A population pharmacokinetic model to predict the individual starting dose of tacrolimus in adult renal transplant recipients. British Journal of Clinical Pharmacology 2019;85(3):601-615. DOI: https://doi.org/10.1111/bcp.13838.
102. Pallet N, Jannot AS, El Bahri M, et al. Kidney transplant recipients carrying the CYP3A4*22 allelic variant have reduced tacrolimus clearance and often reach supratherapeutic tacrolimus concentrations. Am J Transplant 2015;15(3):800-5. (In eng). DOI: 10.1111/ajt.13059.
103. Andreu F, Colom H, Elens L, et al. A New CYP3A5*3 and CYP3A4*22 Cluster Influencing Tacrolimus Target Concentrations: A Population Approach. Clinical Pharmacokinetics 2017;56(8):963-975. DOI: 10.1007/s40262-016-0491-3.
104. Andreu F, Colom H, Grinyo JM, Torras J, Cruzado JM, Lloberas N. Development of a population PK model of tacrolimus for adaptive dosage control in stable kidney transplant patients. Ther Drug Monit 2015;37(2):246-55. DOI: 10.1097/FTD.0000000000000134.
105. Elens L, Haufroid V. Genotype-based tacrolimus dosing guidelines: with or without CYP3A4*22? Pharmacogenomics 2017;18(16):1473-1480. (In eng). DOI: 10.2217/pgs-2017-0131.
106. Elens L, Capron A, van Schaik RH, et al. Impact of CYP3A4*22 allele on tacrolimus pharmacokinetics in early period after renal transplantation: toward updated genotype-based dosage guidelines. Ther Drug Monit 2013;35(5):608-16. (In eng). DOI: 10.1097/FTD.0b013e318296045b.
107. Elens L, Bouamar R, Hesselink DA, et al. A new functional CYP3A4 intron 6 polymorphism significantly affects tacrolimus pharmacokinetics in kidney transplant recipients. Clin Chem 2011;57(11):1574-83. (In eng). DOI: 10.1373/clinchem.2011.165613.
108. de Jonge H, de Loor H, Verbeke K, Vanrenterghem Y, Kuypers DR. In Vivo CYP3A4 Activity, CYP3A5 Genotype, and Hematocrit Predict Tacrolimus Dose Requirements and Clearance in Renal Transplant Patients. Clin Pharmacol Ther 2012;92(3):366-375. DOI: https://doi.org/10.1038/clpt.2012.109.
109. Storset E, Asberg A, Skauby M, et al. Improved Tacrolimus Target Concentration Achievement Using

Computerized Dosing in Renal Transplant Recipients--A Prospective, Randomized Study. Transplantation 2015;99(10):2158-66. DOI: 10.1097/TP.0000000000000708.

110. Staatz CE, Storset E, Bergmann TK, Hennig S, Holford N. Tacrolimus pharmacokinetics after kidney transplantation--Influence of changes in haematocrit and steroid dose. Br J Clin Pharmacol 2015;80(6):1475-6. DOI: 10.1111/bcp.12729.

111. Schijvens AM, van Hesteren FHS, Cornelissen EAM, et al. The potential impact of hematocrit correction on evaluation of tacrolimus target exposure in pediatric kidney transplant patients. Pediatric nephrology (Berlin, Germany) 2019;34(3):507-515. (In eng). DOI: 10.1007/s00467-018-4117-x.

112. Limsrichamrern S, Chanapul C, Mahawithitwong P, et al. Correlation of Hematocrit and Tacrolimus Level in Liver Transplant Recipients. Transplant Proc 2016;48(4):1176-8. (In eng). DOI: 10.1016/j.transproceed.2015.12.096.

113. Sikma MA, Hunault CC, Huitema ADR, De Lange DW, Van Maarseveen EM. Clinical Pharmacokinetics and Impact of Hematocrit on Monitoring and Dosing of Tacrolimus Early After Heart and Lung Transplantation. Clin Pharmacokinet 2020;59(4):403-408. (In eng). DOI: 10.1007/s40262-019-00846-1.

114. Laskow DA, Vincenti F, Neylan JF, Mendez R, Matas AJ. An open-label, concentration-ranging trial of FK506 in primary kidney transplantation: a report of the United States Multicenter FK506 Kidney Transplant Group. Transplantation 1996;62(7):900-5. (In eng). DOI: 10.1097/00007890-199610150-00005.

115. Kershner RP, Fitzsimmons WE. Relationship of FK506 whole blood concentrations and efficacy and toxicity after liver and kidney transplantation. Transplantation 1996;62(7):920-6. (In eng). DOI: 10.1097/00007890-199610150-00009.

116. Vincenti F, Laskow DA, Neylan JF, Mendez R, Matas AJ. One-year follow-up of an open-label trial of FK506 for primary kidney transplantation. A report of the U.S. Multicenter FK506 Kidney Transplant Group. Transplantation 1996;61(11):1576-81. (In eng). DOI: 10.1097/00007890-199606150-00005.

117. Webster AC, Woodroffe RC, Taylor RS, Chapman JR, Craig JC. Tacrolimus versus ciclosporin as primary immunosuppression for kidney transplant recipients: meta-analysis and meta-regression of randomised trial data. BMJ 2005;331(7520):810-810. (In eng). DOI: 10.1136/bmj.38569.471007.AE.

118. Bouamar R, Shuker N, Hesselink DA, et al. Tacrolimus predose concentrations do not predict the risk of acute rejection after renal transplantation: a pooled analysis from three randomized-controlled clinical trials(dagger). Am J Transplant 2013;13(5):1253-61. (In eng). DOI: 10.1111/ajt.12191.

119. Hu R, Barratt DT, Coller JK, Sallustio BC, Somogyi AA. Is There a Temporal Relationship Between Trough Whole Blood Tacrolimus Concentration and Acute Rejection in the First 14 Days After Kidney Transplantation? Ther Drug Monit 2019;41(4):528-532. (In eng). DOI: 10.1097/ftd.0000000000000656.

120. Ntobe-Bunkete B, Lemaitre F. Therapeutic drug monitoring in kidney and liver transplantation: current advances and future directions. Expert Rev Clin Pharmacol 2024;17(5-6):505-514. (In eng). DOI: 10.1080/17512433.2024.2354276.

121. Wallemacq P, Armstrong VW, Brunet M, et al. Opportunities to optimize tacrolimus therapy in solid organ transplantation: report of the European consensus conference. Ther Drug Monit 2009;31(2):139-52. (In eng). DOI: 10.1097/FTD.0b013e318198d092.

122. Lee MN, Butani L. Improved pharmacokinetic monitoring of tacrolimus exposure after pediatric renal transplantation. Pediatric transplantation 2007;11(4):388-93. (In eng). DOI: 10.1111/j.1399-3046.2006.00618.x.
