## Supplementary material for "Protocol for Bayesian combined multi-genotype and concentration informed tacrolimus dosing in paediatric solid organ transplantation (BRUNO-PIC)": Data Information and Databank Guidelines

### Data Information

The following types of data will be recorded within the BRUNO-PIC *REDCap database*.

- Study Number
- Patient Initials
- Year of birth
- Sex at Birth
- Age (at time of transplant)
- Email details for correspondence with participant/parent*
- Solid organ type - Heart/Kidney/Liver
- CYP3A4 and 3A5 genotype*
- Comorbidities
- Tacrolimus concentrations at set time points
- Tacrolimus dose at set time points
- NextDose tacrolimus dose calculations*
- Height, weight, Urea, Electrolytes & creatinine, liver function tests and Haematocrit at each timepoint if performed as part of routine clinical care.
- Medication reconciliation at each study time point, including Drug-Drug Interactions with Tacrolimus

*prospective intervention arm only

The following types of data will be recorded within the BRUNO-PIC Nextdose *website*.

- Patient identifier
- Transplant date
- Date of birth (or current age)
- Sex
- CYP3A4 and 3A5 genotype
- Tacrolimus doses and Time
- Observations incl: weight, concomitant steroid use and dose, Haematocrit. transplant date) are recorded in NextDose.

**Databank guidelines**

| | |
|---|---|
| **Name of the Bank** | ***BRUNO-PIC*** Database |
| **Custodian of the Bank** | Murdoch Children's Research Institute |
| **Purpose of the Data Storage** | The *REDCap* database will store all data related to study endpoints. |
| **Sample / data identifiability** | Upon enrolment in the study, participants will be allocated a study number and de-identified. Patients will be re-identifiable by a restricted group of researchers including Study PI, research assistants or academic pharmacists. |
| **Criteria for participation** | ▪ Age 1- 18 years.<br>▪ Heart OR Liver OR Kidney transplant (planned or on waiting list) excluding repeat liver transplant.<br>▪ Patient is amenable to blood draw from central line.<br>▪ Have no known allergy/hypersensitivity to tacrolimus and/or its formulation.<br>▪ Patient has a signed informed consent.<br>▪ Study enrolment limit has not been reached. |
| **Consent process for obtaining data** | Specific informed Consent will be gained as part of overall study, prior to enrolment. |
| **Sample and data input** | Blood samples will be drawn at enrolment for genotyping (0.5mL – 2mL). |
| **Location of the Bank** | The *REDCap* database is hosted at Murdoch Children's research Institute. |
| **Confidentiality / Security of data** | All patients will be de-identified for the purposes of the study. The study PI, academic pharmacists and research assistants who have been provided with a username/login can see identifiable data for patients. When data are retrieved from the BRUNO-PIC *REDCap database*, all data will be de-identified.<br><br>Backup of data will occur every day as per MCRI's SOP for data security. |
| **Destruction of Data** | Data can be destroyed upon request from the Study PI. |
