## Supplementary material for "Protocol for Bayesian combined multi-genotype and concentration informed tacrolimus dosing in paediatric solid organ transplantation (BRUNO-PIC)": PICF

| | | | |
|---|---|---|---|
| **Study Number:** | 2023/ETH02699 | | |
| **Short Name of Study:** | **BRUNO – PIC** | | |
| **Full Name of Study:** | *Genotype informed **B**ayesian dosing of tac**r**olim**u**s i**n** solid **o**rgan transplant – **P**harmacogenomic **I**mplementation in **C**hildren.* | | |
| **Principal Investigator:** | A/Prof Rachel Conyers and Dr David Metz | | |
| **Version Number:** | 3.0 | **Version Date:** | 08-October--2024 |

Thank you for taking the time to read this **Participant Information and Consent Form**. We are inviting you to take part in a study where we are trying to determine whether knowing your genetic your make-up along with other personal and health information can help to determine a more accurate tacrolimus after you have had a solid organ transplant. This study hopes to improve the dosing accuracy of tacrolimus giving you a more effective dose to prevent organ rejection and avoid potential tacrolimus related side-effects.

This form is 9 pages long. Please make sure you have all the pages.

**What is an Information and Consent Form?**

An Information and Consent Form tells you what is involved if you agree to participate in the study. It helps you decide whether or not you want to take part in the study. Please read it carefully.

Before you make a decision, you can ask us as many questions as you would like so that you are fully informed. You may also want to talk to your family, friends or other healthcare workers before making a decision.

**Taking part in the study is up to you**

You get to choose whether or not you take part in this study. If you decide you do not want to take part, this is ok. It will not affect your relationship with your treating departments at The Royal Children's Hospital or any treatment you receive. If you choose to take part in this study, you understand and agree that there will be no payment of any kind for participating in this study.

**Signing the form**

If you want to take part in this study, please sign the consent at the end of this form. By signing the consent form you are telling us that you:

- Understand what you have read in this form
- Have had a chance to ask questions and received satisfactory answers
- Consent to taking part in the study

We will give you a copy of this Information and Consent Form to keep.

### 1. What is the study about?

We are inviting you to take part in a study called **BRUNO-PIC** 'Genotype informed Bayesian dosing of tacrolimus in solid organ transplant – Pharmacogenomic Implementation in Children'. This study will look at some of your genes that are involved in breaking down medicines, in particular one called tacrolimus (immunosuppressant medicine).

Genes, which are the building blocks of our bodies, affect how we react to medicine. People have about 20,000 genes, and each gene has a specific job. Some genes make enzymes or proteins that break down medicine. If your genes are different from other people's genes, you might break down medicine faster or slower.

In this study our research team are looking to see if your genetic information can help to determine the right dose of the immunosuppressant medicine (tacrolimus) used during your solid-organ transplant. A solid organ transplant can include multiple organs in the body (heart, liver, kidney, lung or intestine) all of which can be transplanted if needed. In your transplant, tacrolimus will be the immunosuppressant most commonly used. It is a highly effective medicine used to help the transplant organ from being rejected by the person the organ is transferred in to.

As we continue to understand what role the genes in our body play in the breakdown of medicine like tacrolimus, we can predict using blood samples at different timepoints a more accurate dose of tacrolimus to reduce the risk of side effects and organ rejection after transplant. The genes we will be looking at in this study in include CYP3A4 and CYP3A5. These are known genes that are related with the enzymes we produce to breakdown tacrolimus in the body.

The current approach to adjusting tacrolimus doses is administer a dose of the medicine and then wait for a blood test result to help us estimate what the future dose should be. This is known as keeping the level of tacrolimus in the body within "therapeutic range". Using the suggested Bayesian approach in this study, will look at the level in the body, the genes that your body has and how they may affect how fast or slow you breakdown tacrolimus and combine this with other personal information we will collect to personalise the tacrolimus dose you receive as an individual. This is sometimes referred to as a personalised medicine.

This study aims to prove that using genotype-informed Bayesian dosing allows us to reach "therapeutic levels" of tacrolimus faster and maintain the tacrolimus levels within the therapeutic range compared to standard weight-based dosing methods. This may result in reducing the risk of rejection especially in the early stages post-transplant, leading to less complications from the transplant.

The primary aim of this study is to use your genotype to predict tacrolimus dosing and to measure the time it is within therapeutic range immediately post-transplant and for the first 2 months of tacrolimus use. We will compare these results to previous transplant patients in which their genotype was not used to predict the dosing of tacrolimus.

### 2. Who is running the study?

This study is being run by the Pharmacogenomics Team at Murdoch Children's Research Institute (MCRI) in collaboration with The Royal Children's Hospital (RCH). A/Prof Rachel Conyers and Dr David Metz are the principal investigators in this study. You might also hear them referred to as the 'PI. As Principal Investigator, they are in charge of this research study and the study team members. They have written the plan for this study – this plan is also called the protocol. As the Principal Investigator, they will be in charge of reviewing the information that we find and collect in this study, and in charge of telling participants and other people about our findings. The study team working with A/Prof Conyers and Dr Metz include Ms Dhrita Khatri (Academic Pharmacist Murdoch Children's Research Institute).

### 3. Why are we asking you to take part?

We are asking you to take part in this study because you are a patient of The Royal Childrens Hospital who is:

- Aged between 1 year to 18 years of age
- Planned for a solid-organ transplant (liver, heart or kidney)
- On a wait list for a solid-organ transplant (liver, heart or kidney)

### 4. What do you need to do this study?

If you agree to take part in this study, your involvement will last approximately 8 weeks.

**Screening**: After potential patients are identified from electronic medical records and multidisciplinary team meetings, the PI or a member from the PIs team will inform you about the study. You will be approached during appointments or on the ward. The study will be explained, questions will be answered, before a blood sample is taken.

**Table 1: What you need to do in this study**

<table>
<tr><th>Part of study</th><th>In-person or electronic?</th><th>How long will it take?</th><th>What does it involve?</th></tr>
<tr><td>Pharmacogenetic Testing</td><td>In-person</td><td>10 minutes</td><td>A 0.5mL – 2mL (~1/2 teaspoon) blood sample will be taken. We will take this sample at the same time as other blood tests are being performed as part of your standard care if possible.<br><br>We will use this sample to run the pharmacogenetic test which is explained below this table.</td></tr>
<tr><td>Tacrolimus levels (blood test)*</td><td>In-person</td><td>10 minutes</td><td><ul>
<li>A 1mL – 3mL (~1/2 teaspoon) blood sample will be taken in the morning prior to taking tacrolimus dose.</li>
<li>All tacrolimus concentrations collected are as part of standard of care, apart from the additional samples as shown in table below.</li>
<li>No venipuncture occurs beyond standard of care, with additional samples taken either via central line (DD4) or at times samples are being taken already as part of standard care.</li>
<li>Typically blood testing will occur daily during the first 2-4 weeks post-transplant and occur less often (i.e., 2-3 times per week) for the second month. The frequency and timing of tacrolimus blood test monitoring will depend on the type of organ transplanted, the transplant protocol, and individual patient factors.</li>
<li>Additional tacrolimus levels may be obtained as needed to optimise dosing and ensure therapeutic drug concentrations as per your treating medical team.</li>
<li>Additional samples for BRUNO-PIC participants:
<ul>
<li>Tacrolimus concentrations will be taken on days 3, 4 and weeks 3 and 8, which will be performed at times when blood is taken as part of clinical care.</li>
<li>These samples will be at the time of morning dose, then 1, 2 and 3h post dose.</li>
</ul></li>
</ul>
<table>
<tr><th>Days post-transplant</th><th>D3</th><th>D4</th><th>D21</th><th>D56</th></tr>
<tr><td></td><td>X</td><td>X</td><td>X</td><td>X</td></tr>
</table>
<br>*This is standard of care test and levels can be taken at the discretion of the treating medical team to ensure tacrolimus is within therapeutic range.</td></tr>
<tr><td>Access to Electronic Medical Record</td><td>Electronic</td><td>N/A</td><td>The study team will access your electronic medical records to review blood test results and to document tacrolimus results in your progress notes. The predicted dose recommendations using the Bayesian modelling “NextDose” program will be documented in the EMR.</td></tr>
</table>

### 5. Blood test for pharmacogenetic testing

The targeted pharmacogenomic panel will look at two genes, CYP3A5 and CYP3A4. The researchers have identified that these two genes are closely linked to how you metabolise tacrolimus. A report will be generated from this test which will summarise your metaboliser state. This means how you are predicted to metabolise (breakdown) the tacrolimus (i.e., fast or slow). The report will be placed in your electronic medical record (EMR). Once the results are in the medical record, they can be used by doctors, pharmacists, nurses and other clinicians to make decisions around your drug therapy.

The pharmacist in the study will use the results of the CYP3A5 and CYP3A4 gene result together with other measurable parameters (weight, height, age, sex, renal function, other concurrent medications, tacrolimus dose and levels). This information will be used to predict an appropriate dose of Tacrolimus for you using a Bayesian dosing calculator. This information will be discussed with your treating team and actioned.

### 6. Optional Future research consent

If you take part in this study, we will ask you to think about letting us contact you about future studies about Pharmacogenomics and individualised prescribing. If you say yes, we will contact you by mail, phone, and/or email. You can say no to this if you want to. If you say no, you can still take part in this BRUNO-PIC study.

### 7. Can you withdraw from the study?

Yes - you can stop taking part in the study at any time. You just need to tell a member of the study team (refer to page 8 for contact details) that you would no longer like to participate. You do not need to tell us the reason why. Once you leave the study, we will no longer use your information. If any information has been collected at study timepoints already up until the point of withdrawal this information may still be used unless you tell us otherwise.

At any time, you can ask us to destroy samples and health information that are stored with us. But we will not be able to destroy any samples and health information that have already been used or shared with researchers. Please only join this research study if you are happy with this approach. If you want to withdraw your samples and health information, please contact the study team.

Upon withdrawal, we will remove all of your medical and genetic information from our study. Information about your consent and withdrawal will be kept for at least 15 years after the end of the study or until you are aged 25 years (whichever is the later), prior to destruction.

### 8. What are the possible benefits for you and other people in the future?

We are doing this study for research purposes. Our aim is to progress our understanding of genotypes and using this information to help predict appropriate Tacrolimus doses. This may benefit you directly, as we may be able to reach the target drug levels of tacrolimus faster and remain within the therapeutic range for a longer time. We hope that this will decrease the chance of your body rejecting the transplanted organ and reduce the likelihood of getting toxic levels of Tacrolimus in your body. This research may also help us to improve the treatment of other children receiving solid organ transplants in the future.

**What are the possible risks, side effects, and inconveniences?**

- There are small risks with taking blood tests, mainly related to bleeding and infection at the site of needle prick. We will make sure that blood tests are done at a time when other routine bloods are taken where possible.
- Additional time will be required to take this extra blood test. You may feel some discomfort during the test. You may feel a sting when we put the needle in your arm. We can use a cream to numb the skin beforehand. You may get some bruising, swelling or bleeding where the needle enters the skin. You may also feel a little light-headed when your blood is taken.
- If you have a central venous access device, we will take bloods from there, if possible to avoid these risks.
- In this study, we are only searching for genes that are related to metabolising Tacrolimus and how this can be used to calculate the appropriate starting dose for you after a transplant.
- There is a small chance that you might become upset because you are taking part in this study. If this happens, you can take a break from the study tests. We can arrange for you to get free counselling or other suitable support. This will be provided by someone who is not part of the research team. You may also decide to withdraw from the study.

**9. What will happen to my blood sample?**

Your blood sample will be sent to researchers on this study within Murdoch Children's Research Institute and Victorian Clinical Genetics Service (VCGS). A small amount of the sample will be used to obtain your genetic results, the remaining sample will be stored in case we need to re-confirm your result. The sample is stored within the VCGS laboratories.

It is unlikely that the sample will be of any commercial value to Murdoch Children's Research Institute or Victorian Clinical Genetics Service. However, it is possible that there may be some commercial value in the future, although it is important to note that any commercial value is likely to be due to findings in a group of patients, rather than from samples from a single patient.

You will not be paid for taking part in the study, nor will you derive financial benefit from future discoveries.

No raw sequencing data will be provided to participants and only the genetic report generated for the purposes of the BRUNO-PIC study will be provided as part of this study.

**10. How will we keep your information confidential?**

We will collect and use personal and health information about you for research purposes. In this study, we will store your electronic information securely on an internal server, the REDCap database, stored at the Murdoch Children's Research Institute. Your consent form will be scanned and stored as a PDF file on the REDCap database. You will be given a study identifier number, so there will not be identifying information on the database. We will keep any paper copies of your information in a locked filling cabinet at the Royal Childrens Hospital.

These people may access your identifiable information:

- Research team members involved with this study, who will come from either The Royal Children's Hospital and Murdoch Children's Research Institute or the Victorian Clinical Genetics Service.
- Relevant Research Ethics and Governance Committee

We will not share your identifiable information with anyone else except as required by law.

**Sharing information**

To advance science, medicine and public health, we may share your de-identified data with any current and future funders, research studies, biobanks, medical journals or data research repositories. Some of these organisations may be located overseas. **Any data that we send overseas is not protected by Australian laws and regulations.** By signing this consent form, you are giving us permission to do this.

If we share your data, we will remove identifying details such as your name, date of birth and address and give the data a special code number.

We will also put security measures in place to protect your data if and when we transfer it to other people. We will make sure any files are completely de-identified and transferred via an encrypted file with access via password only.

Despite our best efforts, there is a small chance that you could be re-identified by someone outside of this research study. In the unlikely event that this happens, someone from the research team will contact you. If, at any point, you think that you may have been re-identified, please let us know.

**Storage of information**

Hardcopy and electronic signed consent forms as well as all research data will be kept for at least 15 years after the end of the study or until you are aged 25 years (whichever is the later), prior to destruction. Specimens in the Victorian Clinical Genetics Service will be kept for at least three years and genetic results will be kept indefinitely. The data will be securely stored at the Murdoch Children's Research Institute. Security measures include storing identifying information separately from the analysis data and keeping tight control access over who can access identifying information.

**11. How will you find out the study results?**

You have the right to access and correct the information we collect and store about you. This is in line with relevant Australian and/or Victorian privacy and other relevant laws. Please contact us if you would like to access this information. At the end of the research study, we may present the results at conferences. We may also publish the results in medical journals. We will do this in a way that protects your privacy.

At the end of the study, we will send you a final letter. This letter will explain what we found out in this study – in other words, our study results. The letter will not have any information specifically about you.

**12. Who should you contact for more information?**

If you would like more information about the study, please contact:

| | |
|---|---|
| **Name:** | A/Prof Rachel Conyers |
| **Contact telephone:** | 03 9936 6770 |
| **Email:** | |

In case of a medical emergency, you should call 000 or attend your nearest hospital's emergency department.

For other urgent matters related to this study, please contact:

| | |
|---|---|
| **Name:** | Dhrita Khatri |
| **Contact telephone:** | 03 9936 6087 |
| **Email:** | |

This study has been approved by the Sydney Children's Hospitals Network (SCHN) HREC (approval number2023/ETH02699). If you have any concerns or complaints about any aspect of the project or the way it is being conducted, you may contact the Executive Officer of the SCHN HREC on (02) 7825 1253 or.

You can contact the Director of Research Operations at The Royal Children's Hospital if you:

- have any concerns or complaints about the study
- are worried about your rights as a research participant
- would like to speak to someone independent of the study.

**Consent Form – Participant**

The Royal Children's Hospital Melbourne

| | |
|---|---|
| **Study Number:** | 2023/ETH02699 |
| **Short Name of Study:** | BRUNO-PIC |
| **Version Number:** | 3.0 **Version Date:** 02-October-2024 |

- I have read this information statement and I understand its contents.
- I understand what I have to do in this study.
- I understand the risks I could face because of my involvement in this study.
- I voluntarily consent to take part in this research study.
- I have had an opportunity to ask questions about the study and I am satisfied with the answers I have received.
- I understand that this study has been approved by the Sydney Children's Hospitals Network Human Research Ethics Committee. I understand that the study is required to be carried out in line with the National Statement on Ethical Conduct in Human Research (2007).
- I understand I will receive a copy of this Information Statement and Consent Form.

**Optional consent**

| | | |
|---|---|---|
| **a. Optional consent: [contact about future studies]**<br>I consent to be contacted about future research studies related to Pharmacogenomics and individualised prescribing. | ☐ I consent | ☐ I **do not** consent |

| | | |
|---|---|---|
| Participant Name | Participant Signature | Date |
| Name of Witness to Participant Signature | Witness Signature | Date |

**Declaration by researcher:** I have explained the study to the participant who has signed above. I believe that they understand the purpose, extent and possible risks of their involvement in this study.

| | | |
|---|---|---|
| Research Team Member Name | Research Team Member Signature | Date |

**Note:** All parties signing the consent form must date their own signature.

| | | | |
|---|---|---|---|
| **Study Number:** | 2023/ETH02699 | | |
| **Short Name of Study:** | **BRUNO – PIC** | | |
| **Full Name of Study:** | *Genotype informed **B**ayesian dosing of tac**r**olim**u**s i**n** solid **o**rgan transplant – **P**harmacogenomic **I**mplementation in **C**hildren.* | | |
| **Principal Investigator:** | A/Prof Rachel Conyers and Dr David Metz | | |
| **Version Number:** | 3.0 | **Version Date:** | 08-October-2024 |

Thank you for taking the time to read this **Parent/Guardian Information and Consent Form**. We are inviting your child to take part in a study where we are trying to determine whether knowing your child's genetic make-up along with other personal and health information can help to determine a more accurate tacrolimus after your child has had a solid organ transplant. This study hopes to improve the dosing accuracy of tacrolimus giving your child a more effective dose to prevent organ rejection and avoid potential tacrolimus related side-effects.

This form is 9 pages long. Please make sure you have all the pages.

#### What is an Information and Consent Form?

An Information and Consent Form tells you what is involved if you agree to participate in the study. It helps you decide whether or not you want to take part in the study. Please read it carefully.

Before you make a decision, you can ask us as many questions as you would like so that you are fully informed. You may also want to talk to your family, friends or other healthcare workers before making a decision.

#### Taking part in the study is up to you

You get to choose whether or not your child takes part in this study. If you decide you do not want your child to take part, this is ok. It will not affect their relationship with their treating departments at The Royal Children's Hospital or any treatment they receive. If you choose for your child to take part in this study, you understand and agree that there will be no payment of any kind for participating in this study.

#### Signing the form

If you want your child to take part in this study, please sign the consent at the end of this form. By signing the consent form you are telling us that you:

- Understand what you have read in this form
- Have had a chance to ask questions and received satisfactory answers
- Consent to your child taking part in the study

We will give you a copy of this Information and Consent Form to keep.

### 1. What is the study about?

We are inviting your child to take part in a study called **BRUNO-PIC** 'Genotype informed Bayesian dosing of tacrolimus in solid organ transplant – Pharmacogenomic Implementation in Children'. This study will look at some of your child's genes that are involved in breaking down medicines, in particular one called tacrolimus (immunosuppressant medicine).

Genes, which are the building blocks of our bodies, affect how we react to medicine. People have about 20,000 genes, and each gene has a specific job. Some genes make enzymes or proteins that break down medicine. If your genes are different from other people's genes, you might break down medicine faster or slower.

In this study our research team are looking to see if your child's genetic information can help to determine the right dose of the immunosuppressant medicine (tacrolimus) used during your child's solid-organ transplant. A solid organ transplant can include multiple organs in the body (heart, liver, kidney, lung or intestine) all of which can be transplanted if needed. In your child's transplant, tacrolimus will be the immunosuppressant most commonly used. It is a highly effective medicine used to help the transplant organ from being rejected by the person the organ is transferred in to.

As we continue to understand what role the genes in our body play in the breakdown of medicine like tacrolimus, we can predict using blood samples at different timepoints a more accurate dose of tacrolimus to reduce the risk of side effects and organ rejection after transplant. The genes we will be looking at in this study in include CYP3A4 and CYP3A5. These are known genes that are related with the enzymes we produce to breakdown tacrolimus in the body.

The current approach to adjusting tacrolimus doses is administer a dose of the medicine and then wait for a blood test result to help us estimate what the future dose should be. This is known as keeping the level of tacrolimus in the body within "therapeutic range". Using the suggested Bayesian approach in this study, will look at the level in the body, the genes that your child's body has and how they may affect how fast or slow your child breakdowns tacrolimus and combine this with other personal information we will collect to personalise the tacrolimus dose your child receives as an individual. This is sometimes referred to as a personalised medicine.

This study aims to prove that using genotype-informed Bayesian dosing allows us to reach "therapeutic levels" of tacrolimus faster and maintain the tacrolimus levels within the therapeutic range compared to standard weight-based dosing methods. This may result in reducing the risk of rejection especially in the early stages post-transplant, leading to less complications from the transplant.

The primary aim of this study is to use your child's genotype to predict tacrolimus dosing and to measure the time it is within therapeutic range immediately post-transplant and for the first 2 months of tacrolimus use. We will compare these results to previous transplant patients in which their genotype was not used to predict the dosing of tacrolimus.

### 2. Who is running the study?

This study is being run by the Pharmacogenomics Team at Murdoch Children's Research Institute (MCRI) in collaboration with The Royal Children's Hospital (RCH). A/Prof Rachel Conyers and Dr David Metz are the principal investigators in this study. You might also hear them referred to as the 'PI. As Principal Investigator, they are in charge of this research study and the study team members. They have written the plan for this study – this plan is also called the protocol. As the Principal Investigator, they will be in charge of reviewing the information that we find and collect in this study, and in charge of telling participants and other people about our findings. The study team working with A/Prof Conyers and Dr Metz include Ms Dhrita Khatri (Academic Pharmacist Murdoch Children's Research Institute).

### 3. Why are we asking you to take part?

We are asking your child to take part in this study because they are a patient of The Royal Childrens Hospital who is:

- Aged between 1 year to 18 years of age
- Planned for a solid-organ transplant (liver, heart or kidney)
- On a wait list for a solid-organ transplant (liver, heart or kidney)

### 4. What do you need to do this study?

If you agree for your child to take part in this study, your child's involvement will last approximately 8 weeks.

**Screening**: After potential patients are identified from electronic medical records and multidisciplinary team meetings, the PI or a member from the PIs team will inform you about the study. You and your child will be approached during appointments or on the ward. The study will be explained, questions will be answered, before a blood sample is taken.

**Table 1: What you need to do in this study**

<table>
<tr><th>Part of study</th><th>In-person or electronic?</th><th>How long will it take?</th><th>What does it involve?</th></tr>
<tr><td>Pharmacogenetic Testing</td><td>In-person</td><td>10 minutes</td><td>A 0.5mL – 2mL (~1/2 teaspoon) blood sample will be taken. We will take this sample at the same time as other blood tests are being performed as part of your child’s standard care if possible.<br><br>We will use this sample to run the pharmacogenetic test which is explained below this table.</td></tr>
<tr><td>Tacrolimus levels (blood test)*</td><td>In-person</td><td>10 minutes</td><td><ul>
<li>A 1mL – 3mL (~1/2 teaspoon) blood sample will be taken in the morning prior to taking tacrolimus dose at a maximum of 9 time points throughout the study.</li>
<li>All tacrolimus concentrations collected are as part of standard of care, apart from the additional samples as shown in table below.</li>
<li>No venipuncture occurs beyond standard of care, with additional samples taken either via central line (DD4) or at times samples are being taken already as part of standard care</li>
<li>Typically blood testing will occur daily during the first 2-4 weeks post-transplant and occur less often (i.e., 2-3 times per week) for the second month. The frequency and timing of tacrolimus blood test monitoring will depend on the type of organ transplanted, the transplant protocol, and individual patient factors.</li>
<li>Additional tacrolimus levels may be obtained as needed to optimise dosing and ensure therapeutic drug concentrations as per your child’s treating medical team.</li>
<li>Additional samples for BRUNO-PIC participants:<ul>
<li>Tacrolimus concentrations will be taken on days 3, 4 and weeks 3 and 8, which will be performed at times when blood is taken as part of clinical care.</li>
<li>These samples will be at the time of morning dose, then 1, 2 and 3h post dose.</li>
</ul></li>
</ul>
<table>
<tr><th>Days post-transplant</th><th>D3</th><th>D4</th><th>D21</th><th>D56</th></tr>
<tr><td></td><td>X</td><td>X</td><td>X</td><td>X</td></tr>
</table>
<br>*This is standard of care test and levels can be taken at the discretion of the treating medical team to ensure tacrolimus is within therapeutic range.</td></tr>
<tr><td>Access to Electronic Medical Record</td><td>Electronic</td><td>N/A</td><td>The study team will access your child’s electronic medical records to review blood test results and to document tacrolimus results in your child’s progress notes. The predicted dose recommendations using the Bayesian modelling “NextDose” program will be documented in the EMR.</td></tr>
</table>

### 5. Blood test for pharmacogenetic testing

The targeted pharmacogenomic panel will look at two genes, CYP3A5 and CYP3A4. The researchers have identified that these two genes are closely linked to how you metabolise tacrolimus. A report will be generated from this test which will summarise your child's metaboliser state. This means how your child is predicted to metabolise (breakdown) the tacrolimus (i.e., fast or slow). The report will be placed in your child's electronic medical record (EMR). Once the results are in the medical record, they can be used by doctors, pharmacists, nurses and other clinicians to make decisions around your child's drug therapy.

The pharmacist in the study will use the results of the CYP3A5 and CYP3A4 gene result together with other measurable parameters (weight, height, age, sex, renal function, other concurrent medications, tacrolimus dose and levels). This information will be used to predict an appropriate dose of Tacrolimus for your child using a Bayesian dosing calculator. This information will be discussed with your child's treating team and actioned.

### 6. Optional Future research consent

If you take part in this study, we will ask you to think about letting us contact you and your child about future studies about Pharmacogenomics and individualised prescribing. If you say yes, we will contact you by mail, phone, and/or email. You can say no to this if you want to. If you say no, your child can still take part in this BRUNO-PIC study.

### 7. Can you withdraw from the study?

Yes - you can stop your child taking part in the study at any time. You just need to tell a member of the study team (refer to page 8 for contact details) that you would no longer like your child to participate. You do not need to tell us the reason why. Once you leave the study, we will no longer use your child's information. If any information has been collected at study timepoints already up until the point of withdrawal this information may still be used unless you tell us otherwise.

At any time, you can ask us to destroy samples and health information that are stored with us. But we will not be able to destroy any samples and health information that have already been used or shared with researchers. Please only join this research study if you are happy with this approach. If you want to withdraw your child's samples and health information, please contact the study team.

Upon withdrawal, we will remove all of your child's medical and genetic information from our study. Information about your consent and withdrawal will be kept for at least 15 years after the end of the study or until your child is aged 25 years (whichever is the later), prior to destruction.

### 8. What are the possible benefits for you and other people in the future?

We are doing this study for research purposes. Our aim is to progress our understanding of genotypes and using this information to help predict appropriate Tacrolimus doses. This may benefit your child directly, as we may be able to reach the target drug levels of tacrolimus faster and remain within the therapeutic range for a longer time. We hope that this will decrease the chance of your child's body rejecting the transplanted organ and reduce the likelihood of getting toxic levels of Tacrolimus in your child's body. This research may also help us to improve the treatment of other children receiving solid organ transplants in the future.

**9. What are the possible risks, side effects, and inconveniences?**

- There are small risks with taking blood tests, mainly related to bleeding and infection at the site of needle prick. We will make sure that blood tests are done at a time when other routine bloods are taken where possible.
- Additional time will be required to take this extra blood test. You may feel some discomfort during the test. You may feel a sting when we put the needle in your child's arm. We can use a cream to numb the skin beforehand. You may get some bruising, swelling or bleeding where the needle enters the skin. You may also feel a little light-headed when your child's blood is taken.
- If you have a central venous access device, we will take bloods from there, if possible to avoid these risks.
- In this study, we are only searching for genes that are related to metabolising Tacrolimus and how this can be used to calculate the appropriate starting dose for you after a transplant.
- There is a small chance that your child might become upset because you are taking part in this study. If this happens, you can take a break from the study tests. We can arrange for you to get free counselling or other suitable support. This will be provided by someone who is not part of the research team. You may also decide to withdraw from the study.

**10. What will happen to my blood sample?**

Your child's blood sample will be sent to researchers on this study within Murdoch Children's Research Institute and Victorian Clinical Genetics Service (VCGS). A small amount of the sample will be used to obtain your child's genetic results, the remaining sample will be stored in case we need to re-confirm your child's result. The sample is stored within the VCGS laboratories.

It is unlikely that the sample will be of any commercial value to Murdoch Children's Research Institute or Victorian Clinical Genetics Service. However, it is possible that there may be some commercial value in the future, although it is important to note that any commercial value is likely to be due to findings in a group of patients, rather than from samples from a single patient.

You and your child will not be paid for taking part in the study, nor will you or your child derive financial benefit from future discoveries.

No raw sequencing data will be provided to participants and only the genetic report generated for the purposes of the BRUNO-PIC study will be provided as part of this study.

**11. How will we keep your child's information confidential?**

We will collect and use personal and health information about your child for research purposes. In this study, we will store your child's electronic information securely on an internal server, the REDCap database, stored at the Murdoch Children's Research Institute. Your child's consent form will be scanned and stored as a PDF file on the REDCap database. Your child will be given a study identifier number, so there will not be identifying information on the database. We will keep any paper copies of your child's information in a locked filling cabinet at the Royal Childrens Hospital.

These people may access your child's identifiable information:

- Research team members involved with this study, who will come from either The Royal Children's Hospital and Murdoch Children's Research Institute or the Victorian Clinical Genetics Service.
- Relevant Research Ethics and Governance Committee

We will not share your child's identifiable information with anyone else except as required by law.

**Sharing information**

To advance science, medicine and public health, we may share your child's de-identified data with any current and future funders, research studies, biobanks, medical journals or data research repositories. Some of these organisations may be located overseas. **Any data that we send overseas is not protected by Australian laws and regulations.** By signing this consent form, you are giving us permission to do this.

If we share your child's data, we will remove identifying details such as your child's name, date of birth and address and give the data a special code number.

We will also put security measures in place to protect your child's data if and when we transfer it to other people. We will make sure any files are completely de-identified and transferred via an encrypted file with access via password only.

Despite our best efforts, there is a small chance that your child could be re-identified by someone outside of this research study. In the unlikely event that this happens, someone from the research team will contact you. If, at any point, you think that your child may have been re-identified, please let us know.

**Storage of information**

Hardcopy and electronic signed consent forms as well as all research data will be kept for at least 15 years after the end of the study or until your child is aged 25 years (whichever is the later), prior to destruction. Specimens in the Victorian Clinical Genetics Service will be kept for at least three years  and genetic results will be kept indefinitely. The data will be securely stored at the Murdoch Children's Research Institute. Security measures include storing identifying information separately from the analysis data and keeping tight control access over who can access identifying information.

**12. How will you find out the study results?**

You have the right to access and correct the information we collect and store about your child. This is in line with relevant Australian and/or Victorian privacy and other relevant laws. Please contact us if you would like to access this information. At the end of the research study, we may present the results at conferences. We may also publish the results in medical journals. We will do this in a way that protects your child's privacy.

At the end of the study, we will send you a final letter. This letter will explain what we found out in this study – in other words, our study results. The letter will not have any information specifically about your child.

**13. Who should you contact for more information?**

If you would like more information about the study, please contact:

| | |
|---|---|
| **Name:** | A/Prof Rachel Conyers |
| **Contact telephone:** | 03 9936 6770 |
| **Email:** | |

In case of a medical emergency, you should call 000 or attend your nearest hospital's emergency department.

For other urgent matters related to this study, please contact:

| | |
|---|---|
| **Name:** | Dhrita Khatri |
| **Contact telephone:** | 03 9936 6087 |
| **Email:** | |

This study has been approved by the Sydney Children's Hospitals Network (SCHN) HREC (approval number: 2023/ETH02699). If you have any concerns or complaints about any aspect of the project or the way it is being conducted, you may contact the Executive Officer of the SCHN HREC on (02) 7825 1253 or.

You can contact the Director of Research Operations at The Royal Children's Hospital if you:

- have any concerns or complaints about the study
- are worried about your child's rights as a research participant
- would like to speak to someone independent of the study.

**Consent Form – Parent/Guardian**

| | | | |
|---|---|---|---|
| **Study Number:** | 2023/ETH02699 | | |
| **Short Name of Study:** | BRUNO-PIC | | |
| **Version Number:** | 3.0 | **Version Date:** | 02-October-2024 |

- I have read this information statement, and I understand its contents.
- I understand what I have to do in this study.
- I understand the risks I could face because of my involvement in this study.
- I voluntarily consent to take part in this research study.
- I have had an opportunity to ask questions about the study and I am satisfied with the answers I have received.
- I understand that this study has been approved by the Sydney Children's Hospitals Network Human Research Ethics Committee. I understand that the study is required to be carried out in line with the National Statement on Ethical Conduct in Human Research (2007).
- I understand I will receive a copy of this Information Statement and Consent Form.

**Optional consent**

| | | |
|---|---|---|
| **a. Optional consent: [contact about future studies]**<br>I consent to be contacted about future research studies related to Pharmacogenomics and individualised prescribing. | ☐ I consent | ☐ I **do not** consent |

____________________________
Participant Name

| | | |
|---|---|---|
| Parent/Guardian Name | Parent/Guardian Signature | Date |
| Name of Witness to Participant Signature | Witness Signature | Date |

**Declaration by researcher:** I have explained the study to the participant who has signed above. I believe that they understand the purpose, extent and possible risks of their involvement in this study.

| | | |
|---|---|---|
| Research Team Member Name | Research Team Member Signature | Date |

**Note:** All parties signing the consent form must date their own signature.
